# A large-scale multi-centre study characterising atrophy heterogeneity in Alzheimer’s disease

**DOI:** 10.1101/2024.08.27.24312499

**Authors:** Vikram Venkatraghavan, Damiano Archetti, Pierrick Bourgeat, Chenyang Jiang, Mara ten Kate, Anna C. van Loenhoud, Rik Ossenkoppele, Charlotte E. Teunissen, Elsmarieke van de Giessen, Yolande A.L. Pijnenburg, Giovanni B. Frisoni, Béla Weiss, Zoltán Vidnyánszky, Tibor Auer, Stanley Durrleman, Alberto Redolfi, Simon M. Laws, Paul Maruff, Australian Imaging Biomarkers and Lifestyle Study, Alzheimer’s Disease Neuroimaging Initiative, E-DADS Consortium, Neil P. Oxtoby, Andre Altmann, Daniel C. Alexander, Wiesje M. van der Flier, Frederik Barkhof, Betty M. Tijms

**Affiliations:** Alzheimer Centre Amsterdam, Neurology, Vrije Universiteit, Amsterdam UMC, location VUmc, Amsterdam, the Netherlands; Amsterdam Neuroscience, Neurodegeneration, Amsterdam, the Netherlands; Laboratory of Neuroinformatics, IRCCS Istituto Centro San Giovanni di Dio Fatebenefratelli, Brescia, Italy; The Australian e-Health Research Centre, CSIRO Health and Biosecurity, Brisbane, Queensland, Australia; Department of Radiology and Nuclear Medicine, Amsterdam Neuroscience, Amsterdam University Medical Center, Location VUmc, Amsterdam, the Netherlands; Clinical Memory Research Unit, Lund University, Sweden; Neurochemistry Laboratory, Department of Laboratory Medicine, Vrije Universiteit Amsterdam, Amsterdam UMC location VUmc, Amsterdam, the Netherlands; Amsterdam Neuroscience, Brain Imaging, Amsterdam, the Netherlands; Laboratory of Neuroimaging of Aging (LANVIE), University of Geneva, Geneva, Switzerland; Geneva Memory Center, Department of Rehabilitation and Geriatrics, Geneva University Hospitals, Geneva, Switzerland; Brain Imaging Centre, HUN-REN Research Centre for Natural Sciences, Budapest, Hungary; Machine Perception Research Laboratory, HUN-REN Institute for Computer Science and Control, Budapest, Hungary; School of Psychology, University of Surrey, Guildford, United Kingdom; Sorbonne Université, Institut du Cerveau - Paris Brain Institute – ICM, CNRS, Inria, Inserm, AP-HP, Hôpital Pitié-Salpêtrière, Paris, France; Centre for Precision Health, Edith Cowan University, Joondalup, Western Australia, Australia; Cogstate Ltd., Melbourne, Victoria, Australia; Centre for Medical Image Computing, Department of Medical Physics and Biomedical Engineering and Department of Computer Science, University College London, UK; Department of Epidemiology and Data Science, Vrije Universiteit, Amsterdam UMC, location VUmc, the Netherlands; Queen Square Institute of Neurology, University College London, UK

**Keywords:** Alzheimer’s disease, heterogeneity, subtypes, data-driven, MRI

## Abstract

**Background:** Previous studies reported on the existence of atrophy-based Alzheimer’s disease (AD) subtypes that associate with distinct clinical symptoms. However, the consistency of AD atrophy subtypes across approaches remains uncertain. This large-scale study aims to assess subtype concordance in individuals using two methods of data-driven subtyping.

**Methods:** We included *n* = 10,011 patients across the clinical spectrum from 10 AD cohorts across Europe, regional volumes using Freesurfer v7.1.1. To characterise atrophy heterogeneity in the AD continuum, we introduced a hybrid two-step approach called Snowphlake (Staging NeurOdegeneration With PHenotype informed progression timeLine of biomarKErs) to identify subtypes and sequence of atrophy-events within each subtype. We compared our results with SuStaIn (Subtype and Stage Inference) which jointly estimates both, and was trained and validated similarly. The training dataset included Aβ+ participants (*n* = 1,195), and a control group of Aβ-cognitively unimpaired participants (*n* = 1,692). We validated model staging within each subtype, in a held-out clinical-validation dataset (*n* = 6,362) comprising patients across the clinical spectrum irrespective of Aβ biomarker status and an independent external dataset (*n* = 762). Furthermore, we validated the clinical significance of the detected subtypes, in a subset of Aβ+ validation datasets with *n* = 1,796 in the held-out sample and *n* = 159 in the external dataset. Lastly, we performed concordance analysis to assess the consistency between the methods.

**Results:** In the AD dementia (AD-D) training data, Snowphlake identified four subtypes: diffuse cortical atrophy (21.1%, *age* 67.5 ± 9.3), parieto-temporal atrophy (19.8%, *age* 60.9 ± 7.9), frontal atrophy (24.8%, *age* 67.6 ± 8.8) and subcortical atrophy (25.1%, 68.3 ± 8.2). The subtypes assigned in Aβ+ validation datasets were associated with alterations in specific cognitive domains (Cohen’s *f:* [0.15 - 0.33]), while staging correlated with Mini-Mental State Examination (MMSE) scores (R: [-0.51 *to* - 0.28]) in the validation datasets. SuStaIn also identified four subtypes: typical (55.7%, *age* 66.7 ± 7.8), limbic-predominant (24.2%, *age* 72.2 ± 6.6), hippocampal-sparing (14.6%, *age* 62.8 ± 6.9), and subcortical (0.8%, *age* 68.2 ± 7.6). The subtypes assigned in Aβ+ validation datasets using SuStaIn were also associated with alterations in specific cognitive domains (Cohen’s *f*: [0.17 - 0.34]), while staging correlated with MMSE scores in the validation datasets (R: [-0.54 *to* - 0.26]). However, we observed low concordance between Snowphlake and SuStaIn, with 39.7% of AD-D patients consistently grouped in concordant subtypes by both the methods.

**Conclusion:** In this multi-cohort study, both Snowphlake and SuStaIn identified four subtypes that were associated with different symptom profiles and atrophy-severity measures that were associated with global cognition. The low concordance between Snowphlake and SuStaIn suggests that heterogeneity may rather be a spectrum than discretised by subtypes.

## Introduction

Alzheimer’s disease (AD) is the leading cause of dementia.^1^ It is characterised by progressive loss of brain volume (atrophy) and cognitive decline. Across individuals with AD, there is substantial variability in severity and pattern of brain atrophy,^2-5^ as well as in the symptoms that AD patients manifest.^6,7^ Understanding the variability in brain atrophy between patients, and how they explain differences in cognitive symptoms, could improve tailored patient care management.

One approach to study heterogeneity in atrophy patterns is by data-driven analysis of structural magnetic resonance imaging (MRI) that quantify *in-vivo* atrophy patterns in AD patients. Previous studies taking this approach, using different techniques, identified either three subtypes^8,9,10^ or four subtypes^4,11^ of AD. The most frequently identified subtypes include a medial temporal lobe (MTL) atrophy subtype and hippocampal-sparing subtype.^2,4,8,9,11^ Other subtypes that have been identified include subcortical atrophy subtype^2,9^, parieto-occipital atrophy subtype^4^, cortical atrophy subtype^2,4,9^, and minimal atrophy subtype.^4,11^ Although these findings suggest that atrophy-based subtypes may represent robust biological entities, there remains inconsistency in the specific subtypes found, number of subtypes found, and in their associations with clinical symptoms. Possibly, this may be explained by difference in methodology used for subtyping, but so far remains unclear to what extent different subtyping methodologies converge on identifying the same subtypes when performed in the same patient population.

Apart from distinct patterns of atrophy, studies have identified another dimension that contributes to atrophy heterogeneity i.e. severity of atrophy (also referred to as atrophy stage).^2,3^ Consequently, it remains a challenge to reliably identify data-driven subtypes that reflect meaningful phenotypic differences independent of disease severity, which might further explain the inconsistencies in atrophy subtypes observed across studies. To overcome this challenge, data-driven disease progression models (DPMs),^12^ such as SuStaIn^2^ and Disease Course Mapping^13^, have been developed to identify subtypes and severity jointly within a single framework. However, these methods remain computationally expensive and thus use a limited number of volumetric (or thickness) markers. Other studies have used regular machine-learning (ML) approaches for subtyping by selecting patients within the same clinical stage of AD^4,9^. While the regular ML approaches are computationally efficient as compared to DPMs and thus scalable to large cohorts and markers with greater spatial resolution, regular ML methods do not account for atrophy severity. To address this drawback, in this work, we combined a well-validated ML approach for AD subtyping using non-negative matrix factorization (NMF)^4,14^ with a scalable disease progression model called discriminative event-based model (DEBM)^15,16^ for estimating severity. The resulting hybrid-method was termed Snowphlake (Staging NeurOdegeneration With PHenotype informed progression timeLine of biomarKErs) and this was used to study AD heterogeneity and compare our results with those obtained using SuStaIn.

In this large-scale multi-centre study including *n* = 10,011 participants from 10 cohorts across Europe, United States, and Australia, we first characterised atrophy heterogeneity in the AD continuum using Snowphlake and compared our results with SuStaIn, trained and validated similarly. Second, we studied how the data-driven estimates of atrophy heterogeneity for each method were related to the cognitive symptoms that patients experience. Finally, we examined the concordance between the subtype assigned by Snowphlake and SuStaIn.

## Methods

### Study participants

We selected participants with a clinical diagnosis of AD dementia (AD-D), mild cognitive impairment (MCI), subjective cognitive decline (SCD), or were cognitively normal (CN) when they had a 3D T1w MRI scan available, from 10 cohorts across Europe, United States of America, and Australia. The included cohorts were: Amsterdam Dementia Cohort (ADC)^17^, Alzheimer’s Disease Neuroimaging Initiative (ADNI)^18^, Australian Imaging Biomarker & Lifestyle Flagship Study of Ageing (AIBL)^19^, National Alzheimer’s Coordinating Center (NACC)^20^, Open Access Series of Imaging Studies (OASIS)^21^, Alzheimer’s Repository Without Borders (ARWiBo)^22^, European DTI Study on Dementia (EDSD)^23^, Italian Alzheimer’s Disease Neuroimaging Initiative (I-ADNI)^24^, European Alzheimer’s Disease Neuroimaging Initiative (also known as PharmaCOG)^25^, and the Geneva memory-centre cohort (GMC)^26^. The characteristics of each cohort are summarized in Supplementary Table 1. ADNI data used in the preparation of this article were obtained from the database adni.loni.usc.edu. Further details about ADNI are mentioned in the Supplementary methods section S1.1. The institutional review boards of all participating institutes approved the protocol for data collection and its subsequent use in retrospective analyses. The clinical diagnosis of participants in each cohort was performed by the different study teams according to international criteria and have been described in detail in each of those cohorts. In the present study we grouped the CN and SCD participants together as cognitively unimpaired (CU).

**Table 1:** Participant Demographics. Values indicated in this table are calculated after automated quality control.

| Cohort | Age [years] | Sex (F/M) | CN and SCD | MCI | AD-D |
| --- | --- | --- | --- | --- | --- |
|  |  |  | *Aβ* Status:<br>– / + / unknown | *Aβ* Status:<br>– / + / unknown | *Aβ* Status:<br>– / + / unknown |
| ADC | 63.9 ± 9.2 | 1,675 / 1,952 | 687 / 184 / 456 | 256 / 328 / 199 | 79 / 1053 / 385 |
| ADNI | 72.1 ± 7.0 | 875 / 909 | 378 / 184 / 113 | 254 / 397 / 177 | 21 / 192 / 68 |
| AIBL | 72.7 ± 6.5 | 298 / 224 | 268 / 91 / 28 | 27 / 49 / 7 | 6 / 43 / 3 |
| ARWiBo | 55.1 ± 16.0 | 482 / 293 | 1 / 0 / 593 | 0 / 14 / 89 | 4 / 10 / 64 |
| EDSD | 70.4 ± 7.3 | 191 / 174 | 0 / 0 / 136 | 24 / 45 / 43 | 0 / 1 / 116 |
| I-ADNI | 72.1 ± 8.0 | 105 / 64 | 0 / 0 / 7 | 0 / 0 / 35 | 0 / 0 / 127 |
| NACC | 71.2 ± 10.2 | 882 / 688 | 358 / 0 / 0 | 39 / 126 / 463 | 18 / 191 / 375 |
| OASIS | 71.9 ± 10.8 | 197 / 105 | 0 / 0 / 185 | 0 / 0 / 90 | 0 / 0 / 27 |
| PharmaCog | 69.0 ± 7.4 | 78 / 57 | 0 / 0 / 0 | 52 / 83 / 0 | 0 / 0 / 0 |
| GMC | 71.5 ± 10.5 | 443 / 319 | 39 / 16 / 151 | 54 / 108 / 257 | 3 / 35 / 99 |
| **Total** | 67.6 ± 10.9 | 5,226 / 4,785 | 1,731 / 475 / 1,669 | 706 / 1,150 / 1,360 | 131 / 1,525 / 1,264 |

Abbreviations: CN: cognitively normal; SCD: subjective cognitive decline; MCI: mild cognitive impairment; AD-D: Clinical diagnosis of AD Dementia.

### Study data, MRI processing and harmonization

Across cohorts, baseline 3D T1w MRI scans were acquired with 44 different MRI scanners, with varied image acquisition protocols. Supplementary Table 2 gives an overview of the scanners included in this study. Cortical reconstruction and volumetric segmentation were performed with a Docker container of Freesurfer v7.1.1 in 3 different centres (ADC and NACC in Amsterdam, ADNI and AIBL in Brisbane, and the rest in Brescia) to extract volumes of 68 cortical regions as per the Desikan-Killiany atlas and 14 subcortical brain regions. Automated quality control for Freesurfer segmentations utilized the Euler number,^27,28^ with outlier thresholds determined independently for each scanner. These thresholds were based on the interquartile range (IQR) specific to each scanner, where outliers were identified as 1.5×IQR below the first quartile.^27,28^ Furthermore, subjects with total intracranial volume (TIV) greater than the threshold of 1.5×IQR above the third quartile computed independently for males and females, were identified as outliers. These outliers were excluded from further analysis in this study. The number of participants excluded based on these two criteria were *n* = 1,198 (10.7%), leaving a total number of scans of *n*= 10,011 participants included for subsequent analyses.

**Table 2:**
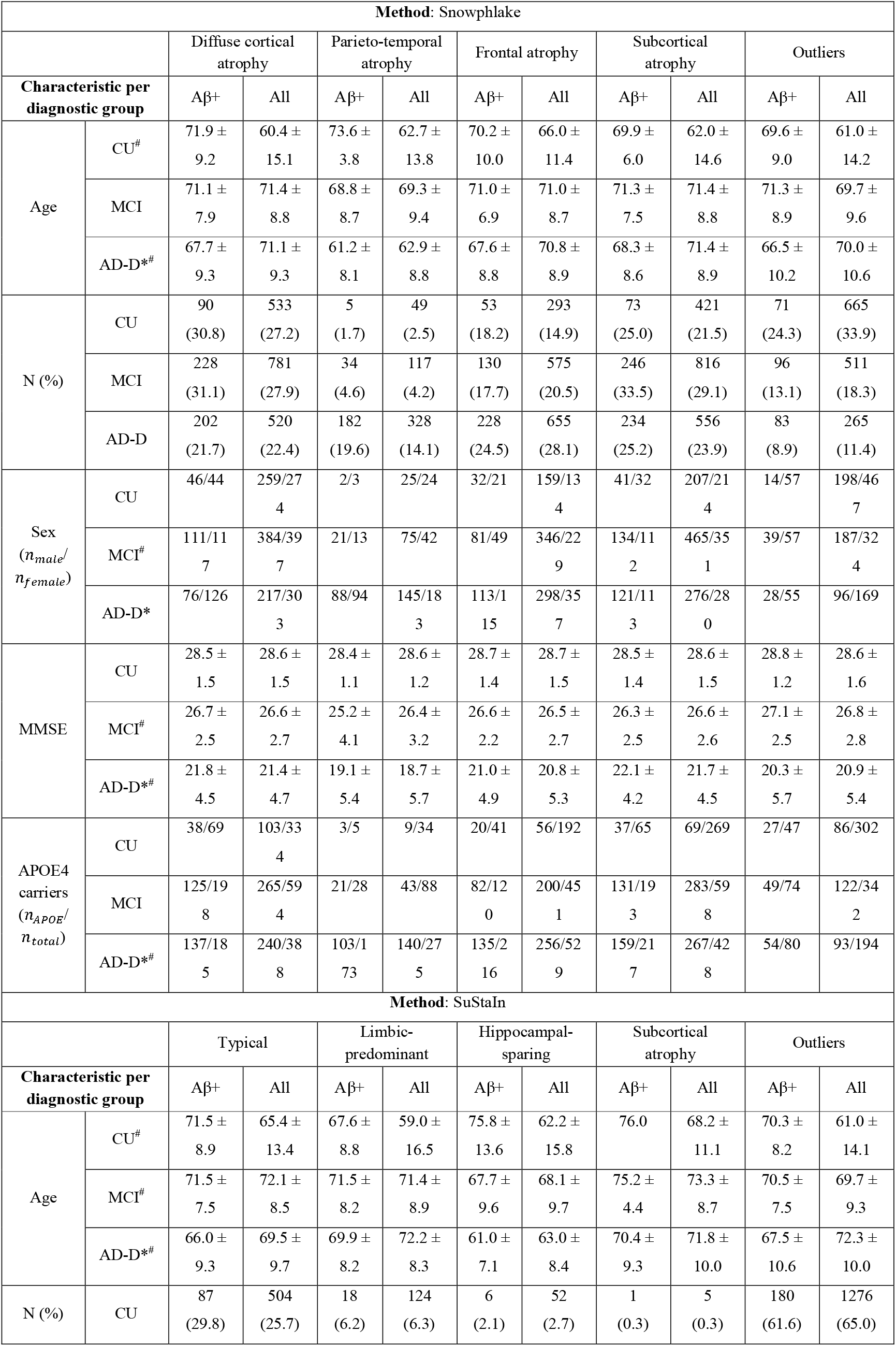
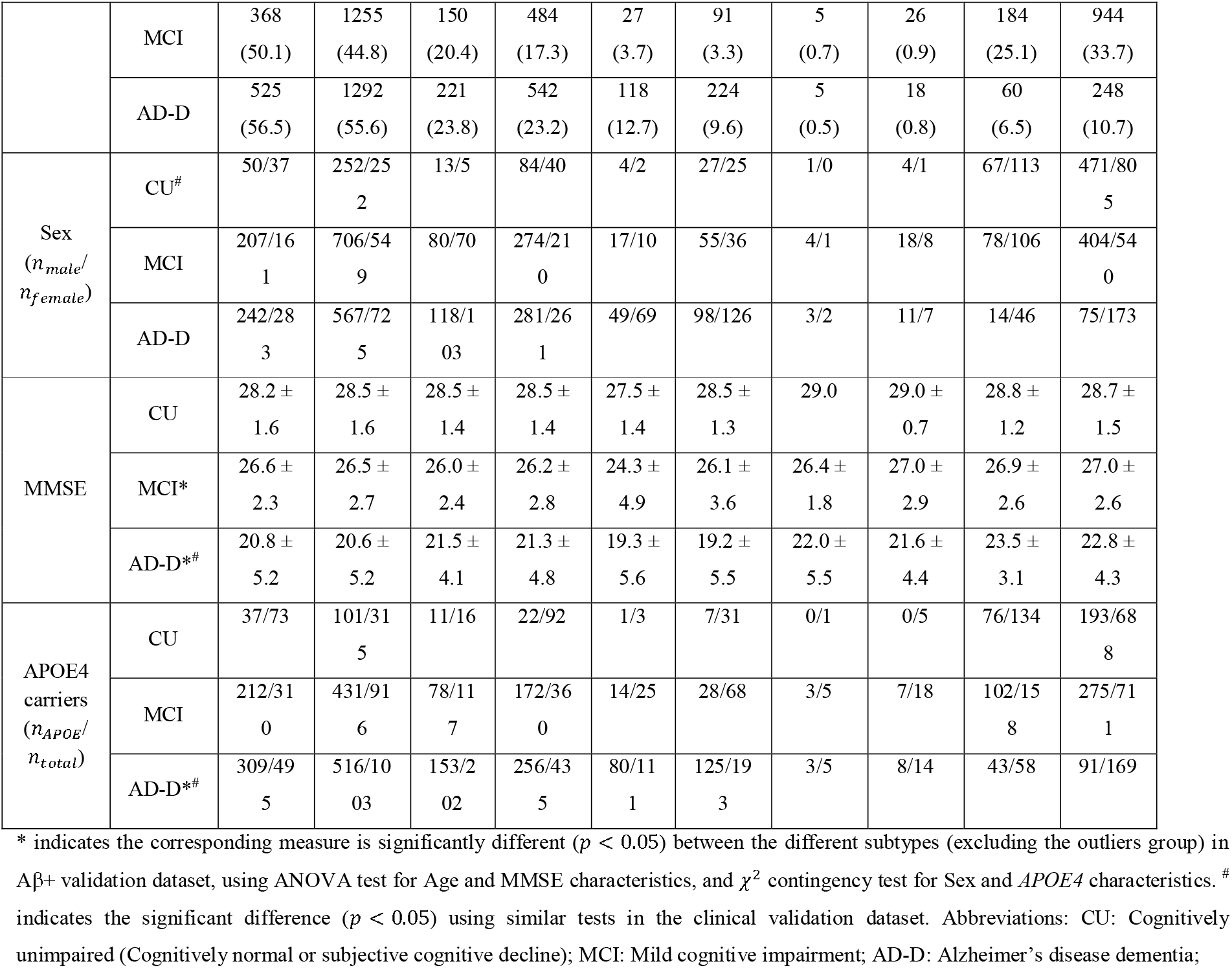
Characteristics of atrophy-based subtypes assigned by the trained Snowphlake and SuStaIn models, pooled across the validation datasets. (held-out validation dataset and external dataset).

We harmonized cortical and subcortical volumes by removing scanner related batch effects while preserving the effects of age, sex, and clinical stage. In our analysis, we used ComBat harmonization^29^ with empirical Bayes optimization to remove batch-related effects, with the largest single-scanner data from the ADNI cohort (Siemens TrioTim 3T scanner, *n* = 257) used as a reference batch. Finally, because SuStaIn is a computationally intensive algorithm and prior subtyping studies using SuStaIn have used between 12 to 21 input features^2,30^, we reduced the number of cortical areas by constructing 24 composite regions, comprising 17 composite cortical ROIs (details of the mapping to derive these composite ROI volume from Freesurfer cortical parcellation are tabulated in Supplementary Table 3) and 7 subcortical regions (namely: Thalamus, Caudate, Putamen, Pallidum, Hippocampus, Amygdala, and Accumbens-area). We corrected for the effects of total intracranial volume and normal aging by regressing out the effects that were estimated in the Aβ-CU participants (see the next section for details on determining amyloid status). The harmonized volumes were combined using the sum of left and right counterparts. These volumes were converted to w-scores (covariate-adjusted z-score) based on the mean and standard deviation of Aβ-CU participants in the study.

### Amyloid Status

Where information about amyloid markers was available, individuals were labelled as having a normal or abnormal amyloid biomarker (Aβ-/ Aβ+ for normal/abnormal respectively) based on either cerebrospinal fluid (CSF, available in ADC, ADNI, ARWiBo, EDSD, PharmaCog, and in NACC after 2015), positron emission tomography (PET) images, or pathological examination (NACC). CSF testing and PET imaging performed during the baseline visit (within a timeframe of 90 days of MRI) were considered for this purpose. Positivity in PET images was determined by either visual readouts by radiologists (available in ADC and GMC), centiloid values (available in ADNI, AIBL, cut-off = 30),^31^ or a combination of the two (in NACC after 2015). The cut-off points for Aβ positivity based on CSF were defined for each cohort independently based on Aβ_1–42_ concentrations. The details of cut-off point selection and assays used are in Supplementary Section S1.2. Details of the Aβ PET processing pipeline and the tracers used are in Supplementary Section S1.3. In ADC, ADNI, and NACC, participants were considered Aβ+ if either one of CSF or PET were positive. In pre-2015 NACC cohort, due to the absence of either of these biomarkers, autopsy-confirmed AD-related neuro-pathologic change (ADNC) based on ABC summary score^32^ (comprising Aβ plaque score, modified Braak stage, modified CERAD score) was used to define Aβ positivity in patients, when available. These scores were categorized as either non-AD, or graded as low, intermediate, or high ADNC in the NACC cohort. In this study, MCI and AD-D participants with low to high ADNC were termed Aβ positive. Participants for whom amyloid status was unavailable were excluded from training the models, and their inclusion in the validation experiments is detailed in the study design.

### Cognitive-data preparation

Neuropsychological test batteries assessing the cognitive domains of episodic memory, attention and executive function, language, and visuospatial function were used to compute composite scores for these domains. Cognitive tests performed during the baseline visit (within a timeframe of 90 days of MRI) were considered for this purpose. Aβ-CU participants’ data were used as a reference group for computing these composite scores. The methodological details of computing the cognitive domain scores in each of our cohorts are included in Supplementary material Section S1.4. We computed the domain scores in the cohorts of ADC, ADNI, AIBL, NACC, and GMC in our analysis. Cognitive test data in the remaining cohorts were not available to us. In the GMC cohort, the language domain score was not computed as the cognitive test battery in that cohort did not include any associated tests for assessing language. Since the different cohorts had different neuropsychological tests to assess the patients, we computed the domain scores independently in the different cohorts, with the Aβ-CU participants in that cohort serving as a reference group to compute z-scores for individual tests. Subsequently, for each domain, multiple test scores belonging to a specific domain were averaged to compute the domain score.

### Study design

We divided our combined cohorts into three different datasets: training dataset, held out clinical-validation dataset, and an independent external dataset. A subset of the clinical-validation dataset and the external dataset with Aβ+ participants was further selected for a few experiments (Aβ+ validation dataset). Figure 1 gives a graphical overview of the study design described here.

**Figure 1:**
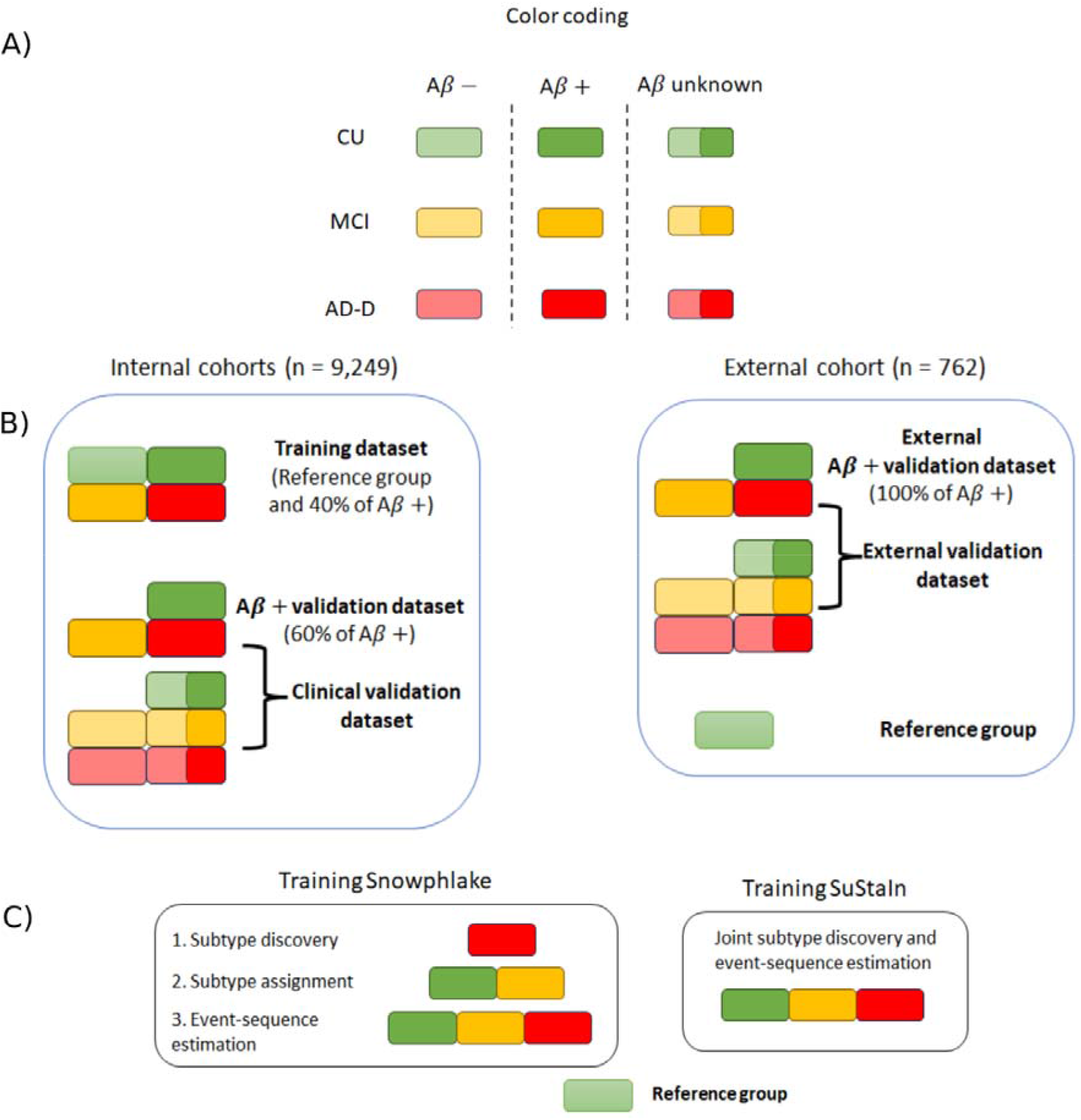
Graphical overview of this study. **A)** Shows the color coding used in the graphical over to denote participants in different clinical stages of the disease as well as their Aβ status. **B)** Overview of the data partitioning into the training dataset, Aβ+ validation dataset, clinical-validation dataset, and external validation datasets, including the inclusion criteria for participants in each dataset. **C)** Overview of the steps involved in training the Snowphlake and SuStaIn models. The reference group shown here is used in both the methods for creating a reference distribution and for w-scoring the imaging biomarkers. Abbreviations: CU: Cognitively unimpaired consisting of both cognitively normal (CN) individuals and subjective cognitive decliners (SCD); MCI: mild cognitively impaired; AD-D: individuals with clinical diagnosis of AD Dementia; + denotes Aβ positivity; - denotes Aβ negativity

The training dataset comprised 40% of the combined Aβ+ participants randomly selected from six cohorts (ADC, ADNI, AIBL, NACC, ARWiBO, EDSD). With the aim of creating atrophy-based subtyping models that are equally generalizable to AD patients across all ages, we ensured the training set had a uniform age distribution. Hence the participants were selected in the training dataset based on weighted random sampling without replacement, with weights inversely proportional to the age distribution in each clinical stage. Moreover, we also included Aβ-CU participants in all the cohorts except GMC to serve as a reference group for training the models.

The held-out clinical-validation dataset consisted of all the participants not included in the training dataset or the reference group from ADC, ADNI, AIBL, NACC, ARWiBO, EDSD, I-ADNI, OASIS, and PharmaCOG. The GMC cohort was chosen as the independent external-validation dataset. The difference between the held-out clinical-validation dataset and the independent external-validation dataset is that for the training dataset all the Aβ+ participants from the GMC cohort were excluded.

The Aβ+ validation datasets comprised the remaining 60% Aβ+ participants not included in training from the aforementioned six cohorts (ADC, ADNI, AIBL, NACC, ARWiBO, EDSD) and 100% Aβ+ participants in the external dataset.

### Characterising atrophy heterogeneity

We used two data-driven approaches for estimating atrophy subtypes and severity: Snowphlake and SuStaIn. Snowphlake is a hybrid method we introduce using non-negative matrix factorization (NMF)^4,14^ for subtyping followed by DEBM^15,16^ for estimating sequence of atrophy-events *within* each subtype. Although each component of this approach has independently been validated before, this is the first study to jointly use them for the purpose of subtype and severity estimation. To ensure easy reproducibility of this approach, we built a python software toolbox: https://github.com/snowphlake-dpm/snowphlake. SuStaIn is a disease progression modelling technique developed previously^2^, with an existing python software package.^33^ The key conceptual difference is that Snowphlake is a two-step subtype-then-stage approach, while SuStaIn estimates both subtype and stage jointly.

#### Snowphlake

The subtyping model was trained on Aβ+ AD-D participants, using the non-smooth variant of non-negative matrix factorization (ns-NMF)^4,34^ with KL-divergence as a distance metric. Ns-NMF is a stochastic dual-clustering approach that is designed to estimate sparse clusters in the data. With different random initializations resulting in slightly different subtypes, ns-NMF was run *n*_*run*_ times on the training data, where *n*_*run*_ = 25 X *n*_*AD*-*D*_. Here, *n*_*AD*-*D*_ was the number of Aβ+ AD-D participants in the training data. The run with the least residual of subtyping (*res*_*k*_) was chosen as the optimal solution, where *k* is the number of subtypes. For choosing the optimal number of subtypes (*n*_*opt*_), a random permutation of the training data was subsequently subtyped. *k*_*opt*_is chosen such that Δ*residual*= *res*_*k*-l_ -*res*_*k*_ for the training data is higher than that in the random permutations. On subtyping, each participant is assigned a weight for each subtype. These subtype weights were further used to detect outliers within each subtype based on minimum covariance determinant algorithm^35^ with Mahalanobis distance metric.

Next, based on the identified subtypes, we assigned the subtypes of Aβ+ *MCI*and Aβ+ *CU* participants in the training dataset. We then determined the sequences of atrophy-events for each subtype using co-initialized discriminative event-based model (DEBM).^15,16^ Briefly, Gaussian mixture modelling (GMM) was used to estimate the probabilities for each region to be abnormal for each participant, with Aβ-CU group considered as a reference group for GMM. These probabilistic abnormality values were used to infer a sequence for each Aβ+ participant in the training data. These individual estimates were aggregated using generalized Mallows model^15^ to estimate the sequence of atrophy-events for each subtype. Further details about training DEBM are in Supplementary Section S1.5.

#### SuStaIn

We trained SuStaIn on the same training data as used in Snowphlake, with the pySuStaIn toolbox^33^. We used the cross-validation information criteria (CVIC) for selecting optimum number of subtypes, with *w* = -1 and *w*= -2 chosen as event thresholds. The methodological details of the SuStaIn approach has been described in detail in *Young et al*^*2*^. For the sake of completeness, the method has been briefly described in Supplementary Section S1.6.

For both Snowphlake and SuStaIn, the trained models were used to assign atrophy-based subtype and stage to participant data in the different validation datasets.

### Statistical analysis to characterise subtypes and evaluate concordance of assigned subtypes

The subtype and staging measures assigned in the Aβ+ validation dataset, clinical-validation dataset, and external-validation dataset by both methods, were used further for investigating if these measures were associated with symptom profile and severity respectively.

#### Experiment 1: Validating the estimated staging

To evaluate the staging system of Snowphlake and SuStaIn, the trained models were used to assign the subtypes and stages of all participants in the Aβ+ validation dataset, clinical-validation dataset, and external-validation dataset. The assigned stage within each subtype was used to compute Pearson’s correlation with Mini-Mental Status Examination (MMSE), as a proxy for disease severity.

#### Experiment 2: Comparison of subtypes on cognitive symptoms

In the absence of ground-truth in data-driven subtyping, we used the association of the identified subtypes with the patients’ cognitive-symptom profile, to determine their validity. We performed analysis of variance (ANOVA) tests in MCI and AD-D patients in Aβ+ validation dataset and Aβ+ subset of the external-validation dataset to determine if subtypes differed in terms of deficits in specific cognitive domains, after correction for confounding effects of age, sex, and level of education. These statistical tests were performed for both subtyping methods in the validation datasets, independently in each of ADC, ADNI, NACC, AIBL, and GMC. Lastly, we performed random-effect meta-analysis pooling the results of independent cohorts and accounted for multiple testing using false discovery rate (FDR) correction.

#### Experiment 3: Concordance between Snowphlake and SuStaIn

The motivation to investigate concordance between the methods was to go beyond group-level definitions of subtypes to individuals assigned to these subtypes. High concordance between the two methods would indicate individual patients in different subtypes have distinct atrophy pattern much like their group-level definitions, while low concordance would indicate individual atrophy patterns vary substantially even within each subtype. To quantify the concordance between the two methods, we constructed contingency matrices of participant subtypes by Snowphlake and SuStaIn for Aβ+ CU, MCI, and AD-D in the training and in the validation dataset. Concordant subtype-pairs are defined as the Snowphlake subtype most frequently co-occurring with SuStaIn subtypes identified in Aβ+ AD-D patients.

We performed additional analyses for identifying atrophy pattern determinants for participants to not be grouped into concordant subtype-pairs. To this end, we created average w-score volume maps for all AD-D participants in the concordant subtype-pairs and their less frequently co-occurring counterparts. We performed a t-test between these average w-score maps, to investigate if there are significant differences in atrophy patterns that influence concordance between the two methods. The null hypothesis in this test is that there is no difference in average w-scores of participants within a specific SuStaIn subtype, grouped in different subtypes of Snowphlake. Lastly, we estimated the sequence of atrophy-events in the concordant subtype-pairs using DEBM, the methodological equivalent of Snowphlake with 1-subtype and w-score EBM, the methodological equivalent of SuStaIn with 1-subtype.

## Results

The demographics of participants and their amyloid status are summarized in Table 1. Overall, our combined dataset (from 10 cohorts) consisted of *n* = 3,150 Aβ+ participants (*n*_*ADD*_ = 1,525 ; *n*_*MCI*_= 1,150 ; *n*_*CU*_ = 475), *n* = 2,568 Aβ-participants (*n*_*ADD*_ = 131; *n*_*MCI*_= 706; *n*_*CU*_ = 1,731), and *n* = 4,293 participants with unknown Aβ status (*n*_*ADD*_ = 1,264; *n*_*MCI*_= 1,360; *n*_*CU*_ = 1,669). This combined dataset was divided into a training dataset, held-out validation dataset, and an external-validation dataset. The training dataset consisted of *n* = 1,195 Aβ+ participants (*n*_*ADD*_ = 596; *n*_*MCI*_= 416; *n*_*CU*_ = 183) and *n* = 1,692 Aβ-CU reference group participants. The held-out validation dataset consisted of *n* = 6,362 participants across the clinical spectrum (*n*_*ADD*_ = 2,187; *n*_*MCI*_= 2,381; *n*_*CU*_ = 1,794) and the external dataset consisted of *n* = 723 patients (*n*_*ADD*_ = 137; *n*_*MCI*_= 419; *n*_*CU*_ = 167) and *n*= 39 Aβ-CU reference group participants. A subset of participants in the validation datasets with Aβ+ status (Aβ+ validation dataset) consisted of *n* = 1,796 participants in the internal cohorts (*n*_*ADD*_ = 894; *n*_*MCI*_= 626; *n*_*CU*_ = 276) and *n* = 159 participants in the external cohort.

The age of the *n* = 1,525 Aβ+ AD-D patients included in our study was 66.8 ± 8.7 years (see Supplementary Figure 1), with ADC predominantly being a young-onset AD cohort, while the rest being predominantly late-onset AD cohorts. Supplementary Figure 1 also shows the age distribution in the different clinical stages within the Aβ+ patient population and in our selected training dataset. 52.2% (5226/10,011) of the included participants were women, while 47.6% (569/1195) of the Aβ+ patients included in the training dataset were women. Furthermore, all the imaging markers used in this study except Pallidum volume were different between the Aβ+ AD-D patients and Aβ-CU reference group with *p* > 0.05 for Pallidum and *p* < 10^−5^ for all other markers, after correcting for multiple testing with FDR.

### Subtypes identified with Snowphlake and SuStaIn

Snowphlake and SuStaIn each identified four subtypes. Supplementary Figure 2 shows the criteria used for selecting the optimum number of subtypes for each modelling technique (Δ*residual*for Snowphlake, CVIC for SuStaIn). For SuStaIn, the CVIC value for the 5-subtype solution was marginally better than the 4-subtype solution. However, only 3/1,195 Aβ+ patients in the training data belonged to 5^th^ subtype. We hence chose the 4-subtype solution for our further analysis.

The atrophy subtypes identified by Snowphlake, along with the prevalence of each subtype and age distribution among AD-D Aβ+ patients in the training and Aβ+ validation datasets were: Diffuse cortical atrophy subtype (*Training*: 21.6% (*n* = 129/596), *age* = 66.5 ± 7.8; Aβ + *Validation*: 21.1% (*n* = 189/894), *age* = 67.5 ± 9.4), Parieto-temporal atrophy subtype (*Training*: 19.2% (*n* = 115/596), *age* = 63.1 ± 6.9; Aβ + *ValiDation*: 19.7%(*n* = 177/896), *age* = 60.9 ± 7.9), Frontal atrophy subtype (*Training*: 25.5% (*n* = 152/596), *age* = 68.3 ± 7.9; Aβ + *ValiDation*: 24.8% (*n* = 222/894), *age* = 67.6 ± 8.9), and Subcortical atrophy subtype (*Training*: 24.8% (*n* = 148/596), *age* = 70.0 ± 7.2; Aβ + *ValiDation*: 25.2% (*n* = 225/894), *age* = 68.3 ± 8.3), with prominent temporal lobe atrophy in each of the identified subtypes. Apart from these subtypes, an additional outlier group not assigned to any subtype was detected (Training: 8.7%; Aβ + *ValiDation*: 9.1%). Figure 2 depicts the sequence of atrophy-events estimated for each subtype by Snowphlake. Supplementary Figure 3 shows the uncertainty in these estimates.

**Figure 2:**
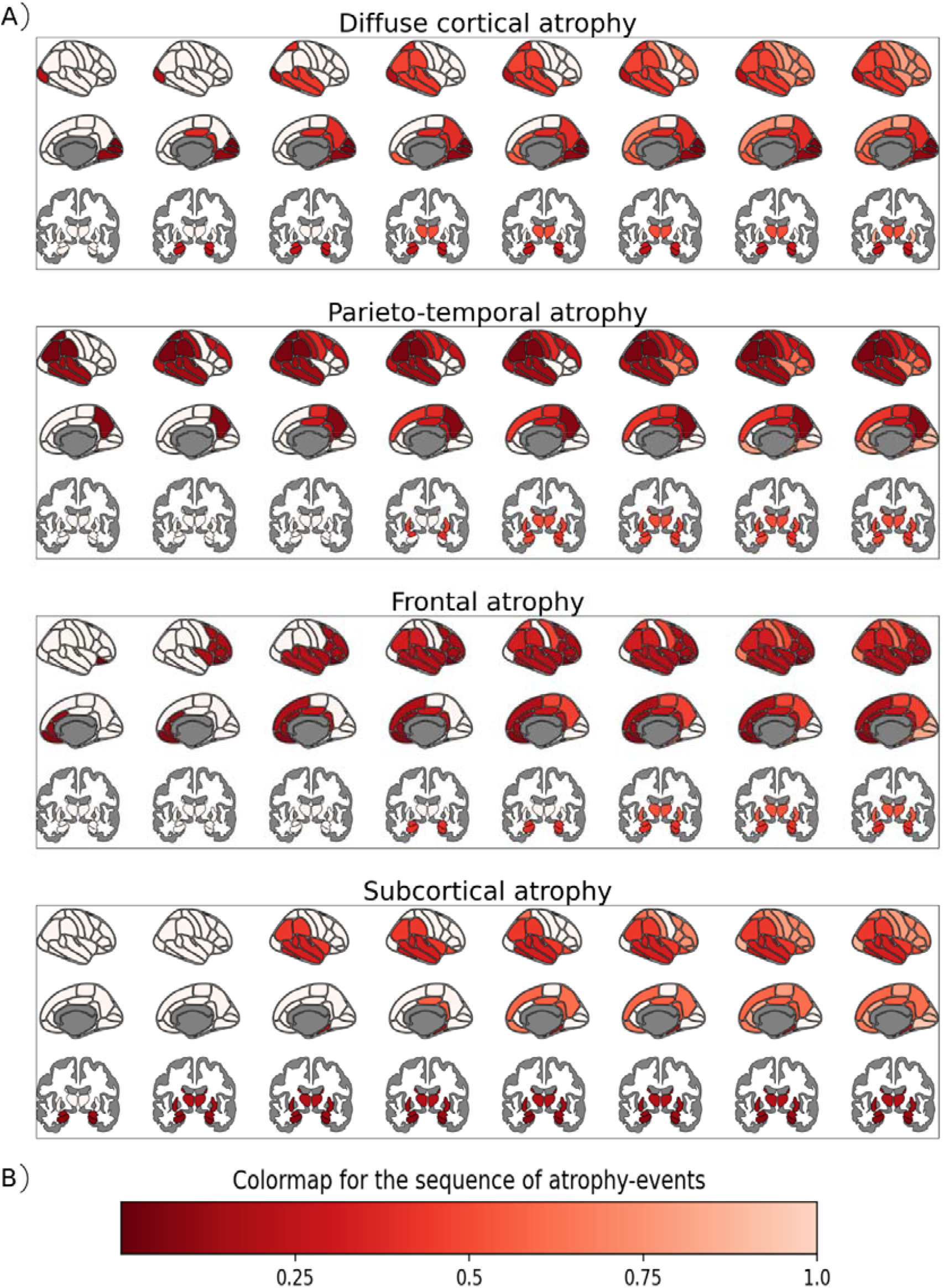
Snowphlake modelling in the Aβ+ participants in the multi-cohort harmonized training dataset. **A)** These plots depict the subtypes and sequence of atrophy-events for each subtype estimated. Within each subtype, the x-axis corresponds to the stage of the disease. Each column shows the brain in its lateral, medial, and subcortical views, with the regions that is expected to be abnormal at this stage for the subtype in shades of red and unaffected regions in white. **B)** The scale for the colour map goes from 0 to 1, the normalized staging scale for Snowphlake, where 0 represents a region becoming abnormal at the earliest stages of the disease and 1 represents late stage.

The prevalence, age and MMSE distribution, and the percentage of *APOE4* carriers in each of these atrophy subtypes across the different clinical stages in the pooled validation datasets (held-out validation and external validation pooled together) are summarized in Table 2 and these results in each cohort independently are reported in Supplementary Table 4. Age of onset of AD-D differed significantly (*p* < 0.05) between the four identified subtypes in the pooled validation datasets, as well as in each of the cohorts independently, except EDSD (*p* = 0.11), with Parieto-temporal atrophy subtype consisting of the youngest AD-D patients (61.2 ± 8.1) and subcortical atrophy subtype the oldest (68.3 ± 8.6). In ADNI, AIBL, ARWiBo, I-ADNI and OASIS cohorts, MMSE of the AD-D patients in different subtypes were not significantly different (*p* > 0.05), indicating the identified subtypes and severity were disentangled. In ADC, EDSD and NACC cohorts, MMSE of AD-D patients was significantly different (*p* < 0.05) between subtypes, with the Parieto-temporal atrophy subtype having the lowest MMSE among the four subtypes. Percentage of *APOE4* carriers was significantly different (*p* < 0.05) in the AD-D dementia patients in the pooled validation datasets. The percentage of outliers across all Aβ+ validation datasets decreased with clinical stage (CU: 25.0%, MCI: 12.4%, AD-D: 9.1%) indicating that characteristic atrophy patterns emerge as the disease progresses.

Supplementary Figure 4 depicts the atrophy subtypes and sequence of atrophy-events estimated by SuStaIn and Supplementary Figure 5 shows the posterior probability distribution of these sequences using Markov chain Monte Carlo (MCMC) sampling, interpreted as the uncertainty in this estimation. The identified subtypes were Typical subtype (with early hippocampus and temporal lobe atrophy), Limbic predominant subtype, Hippocampal sparing subtype, and Subcortical subtype. The prevalence of these subtypes and age distribution among AD-D participants in the training and Aβ+ validation dataset were: Typical (*Training*: 55.7% (*n* = 332/596), *age* = 66.7 ± 7.8; Aβ + *Validation*: 56.0% (*n* = 501/894), *age* = 65.8 ± 9.3), Limbic predominant (*Training*: 24.1% (*n* = 144/ 596), *age* = 72.2 ± 6.6; Aβ + *Validation*: 24.0%(215/894), *age* = 69.8 ± 8.2), Hippocampal sparing (*Training*: 14.5% (*n* = 87/596), *age* = 62.8 ± 6.9; Aβ + *Validation*: 12.9% (*n* = 115/894) *age* = 60.9 ± 7.0), Subcortical atrophy (*Training*: 0.8% (*n* = 5/596), *age* = 68.2 ± 7.6; Aβ + *Validation*: 0.5% (*n* = 5/894), *age* = 70.4 ± 9.3). Apart from these subtypes, an outlier group (defined as AD-D patients in stage 0) was detected (Training: 4.7%; Aβ + *Validation*: 6.5%) The prevalence, age and MMSE distribution, and the percentage of *APOE4* carriers in each of these subtypes across the different diagnostic categories in the pooled validation datasets have been summarized in Table 2 and these results in each cohort independently are reported in Supplementary Table 5. Age of onset of AD dementia and *APOE4* carriership percentage differed significantly (*p* < 0.05) between the four subtypes identified by SuStaIn with Hippocampal sparing subtype consisting of the youngest AD-D patients (61.0 ± 7.1) and Limbic-predominant and Subcortical atrophy subtypes the oldest (69.9 ± 8.2 and 70.4 ± 9.3 respectively).

#### Experiment 1: Atrophy-based model stage correlates with MMSE

Figure 3 depicts the correlation between the atrophy-based patient stage assigned by Snowphlake for the clinical-validation dataset and external dataset, with MMSE, a clinical screening tool for measuring disease severity of the patient. The atrophy-based stage showed significant correlation with MMSE within all four subtypes, with higher atrophy stage related to worse MMSE scores (*R* = -0.51 *to* -0.28), with *p* < 0.0001 in the clinical-validation dataset and *p* < 0.05 in the external-validation dataset. The distribution of atrophy-based stages for the different diagnostic groups (of CU, MCI, AD-D) were different (*p* < 0.0001) and are also shown in Figure 3. Supplementary Figure 6 depicts a similar plot for these correlations for the Aβ+ validation dataset and the Aβ+ subset of the external dataset. Supplementary Figure 7 shows the correlation between the atrophy-based patient stage assigned by SuStaIn for the clinical-validation dataset and external dataset, with MMSE. The atrophy-based stage assigned by SuStaIn showed significant correlation with MMSE within all subtypes except in the subcortical atrophy subtype (*R* = -0.54 *to* - 0.26) with *p*< 0.0001 in the clinical-validation dataset and *p*< 0.01 in the external-validation dataset and *p* > 0.05 for the subcortical atrophy subtype in both validation datasets.

**Figure 3:**
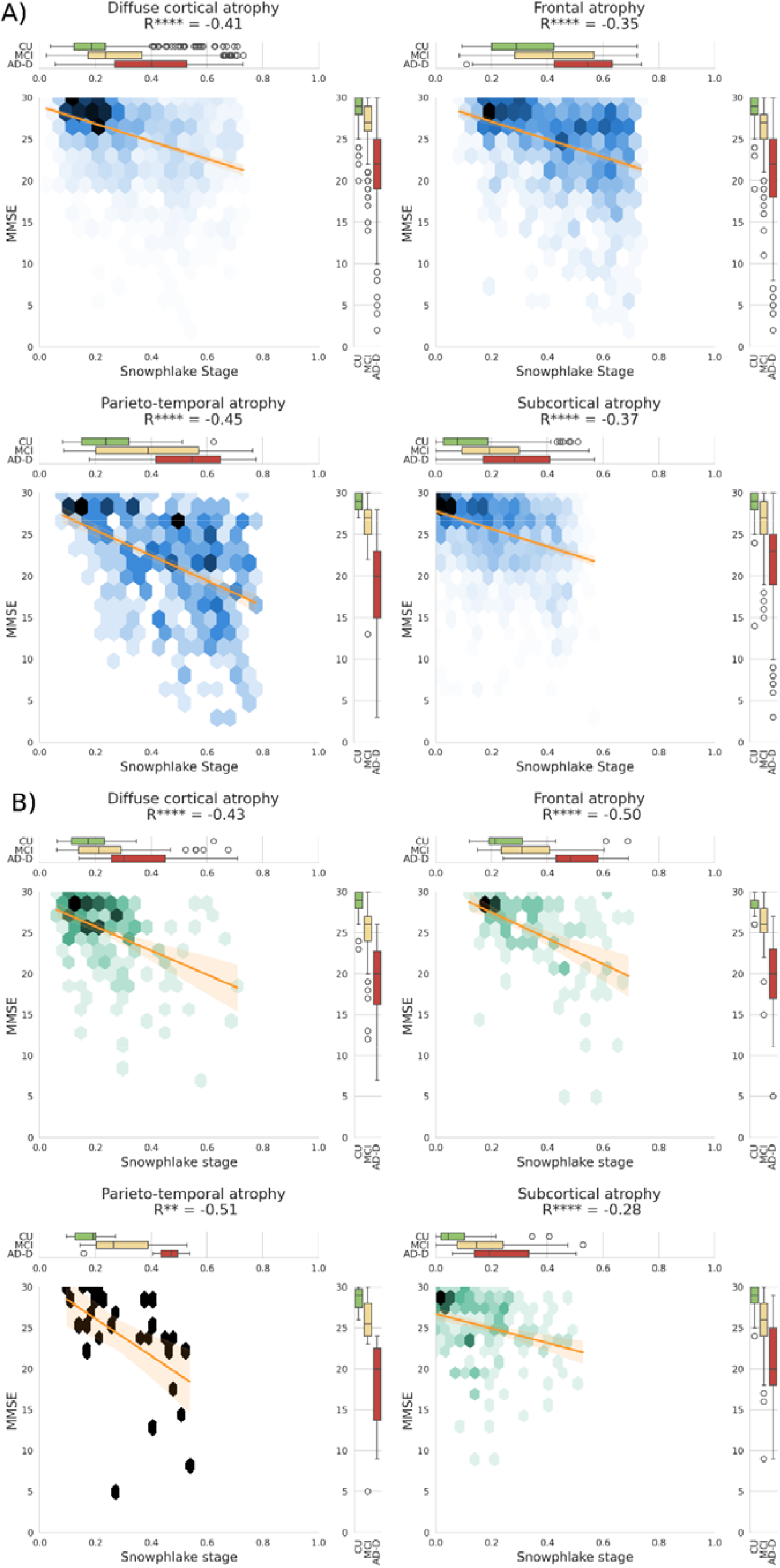
Experiment 1: Correlation of the estimated stage (measuring atrophy severity) using Snowphlake with MMSE in A) clinical-validation cohort B) external-validation cohort. Figures A) and B) both consists of 4 hex-plots, one for each subtype assigned by the trained Snowphlake model. The colour of a bin in the hex-plot denotes the relative proportion of the participants. The boxplot on top of each hex-plot shows the distribution of estimated Snowphlake stage for the participants in the different clinical groups. The boxplot at the right of each hex-plot shows the distribution of MMSE in the different clinical groups. The line overlaying on each hex-plot shows the regression line between MMSE and Snowphlake’s stage. The text on top of each hex-plot shows the correlation coefficient (R) between estimated stage and MMSE. The asterisk (*) next to R denotes the significance level. * corresponds to *p* < 0.05; ** corresponds to *p* < 0.01; *** corresponds to *p* < 0.001; **** corresponds to *p* < 0.0001. Abbreviations: CU: Cognitively unimpaired (Cognitively normal or subjective cognitive decline); MCI: Mild cognitive impairment; AD-D: Alzheimer’s disease dementia;

#### Experiment 2: Cognitive domain characteristics of the subtypes

Figure 4 shows the effect sizes (Cohen’s f-statistic) of cognitive domain score differences between subtypes identified by Snowphlake and SuStaIn, for the diagnostic groups of MCI and AD-D. These subtype differences are computed for participants in the Aβ+ validation cohorts of ADC, ADNI, NACC, AIBL, and GMC for the cognitive domains of memory, executive function and attention, language, and visuospatial function. For Snowphlake, the mean effect sizes for the four cognitive domains were between *f* = 0.15 *to* 0.33 in the AD-D group, and were between *f* = 0.15 *to* 0.24 in the MCI group. For SuStaIn, the mean effect sizes for the four cognitive domains were between *f* = 0.17 *to* 0.34 in the AD-D group and were between *f* = 0.08 *to* 0.20 in the MCI group. There were no significant differences between the effect sizes of Snowphlake and SuStaIn for AD-D patients, while Snowphlake was significantly better at detecting differences in the language domain for MCI patients (FDR-corrected *p* = 0.016) than SuStaIn’s subtypes. There was significant heterogeneity (based on Cochran’s Q statistic) observed between cohorts for both the methods, for both the diagnostic groups. Supplementary Figure 8, depicts the subtype differences in the distribution of cognitive domain scores independently in each cohort for Snowphlake subtypes, which further highlights the differences in associations across different cohorts. Supplementary Figure 9 shows similar associations between the subtypes and cognitive domain scores in different cohorts for the subtypes identified by SuStaIn.

**Figure 4:**
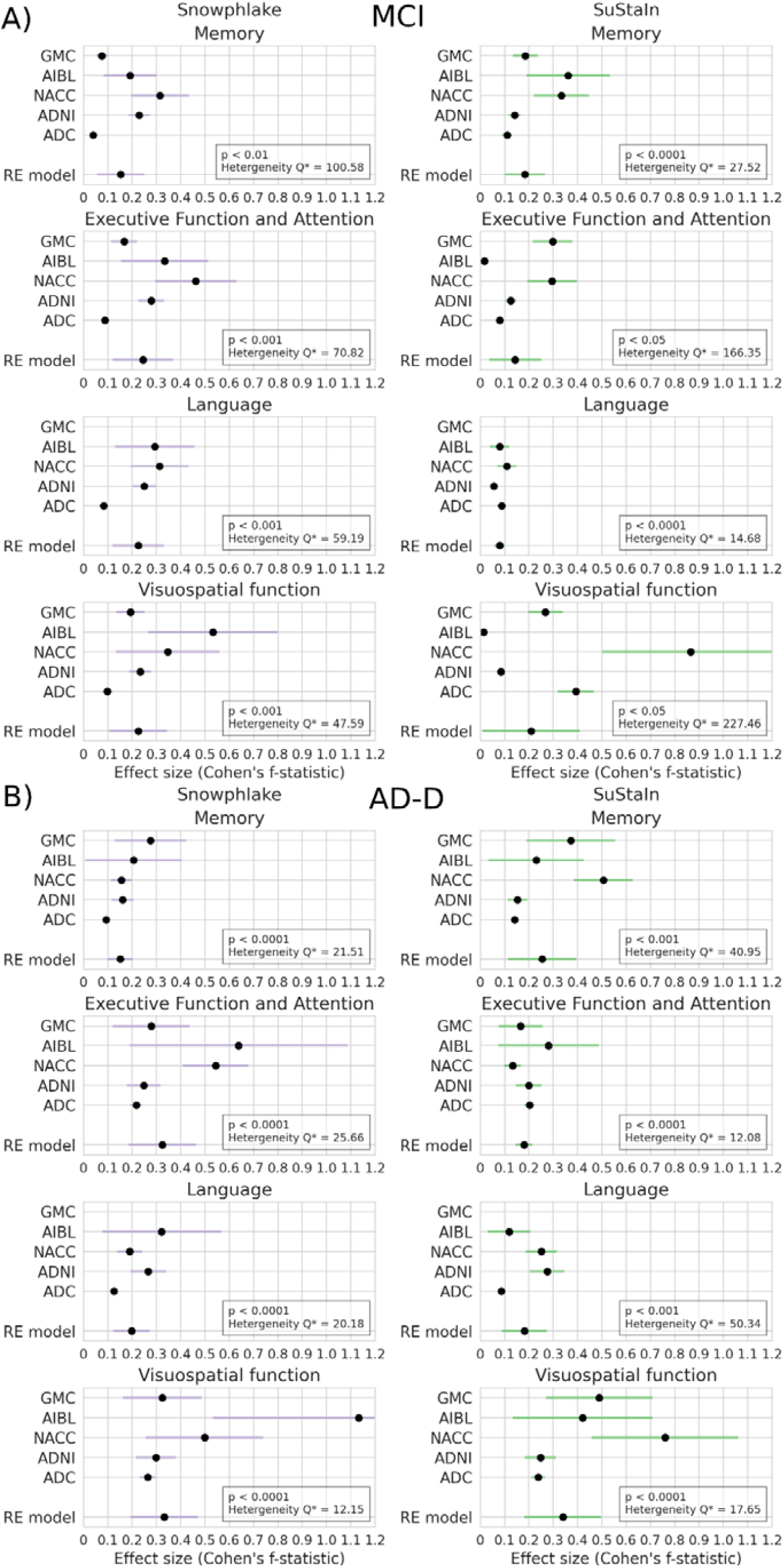
Experiment 2: Cognitive domain differences between subtypes assigned in the Aβ+ validation datasets. Cognitive domain differences are shown for subtypes assigned by Snowphlake (left) and SuStaIn (right) in **A)** MCI patients and **B)** AD-D patients. Each sub-plot shows the effect size (Cohen’s f-statistic) and its confidence internal for a cognitive domain in 5 different cohorts within the Aβ+ validation datasets. The combined effect-size of the random effect (RE) model obtained via meta-analysis across the different cohorts, and the corresponding confidence internal is shown within each subplot as well. The p-value corresponding to the RE model and the Cochran’s Q statistic measuring heterogeneity across cohorts is shown at the bottom right of each sub-plot. The Q* indicates that the shown Cochran’s Q statistic is significant (< 0.0001).

#### Experiment 3: Concordant subtype-pairs

When comparing how participants were clustered, we observed a low concordance between Snowphlake and SuStaIn. Figure 5 shows the contingency matrices between Snowphlake and SuStaIn subtype assignments in different clinical stages of Aβ+ participants, in the training and validation datasets.

**Figure 5:**
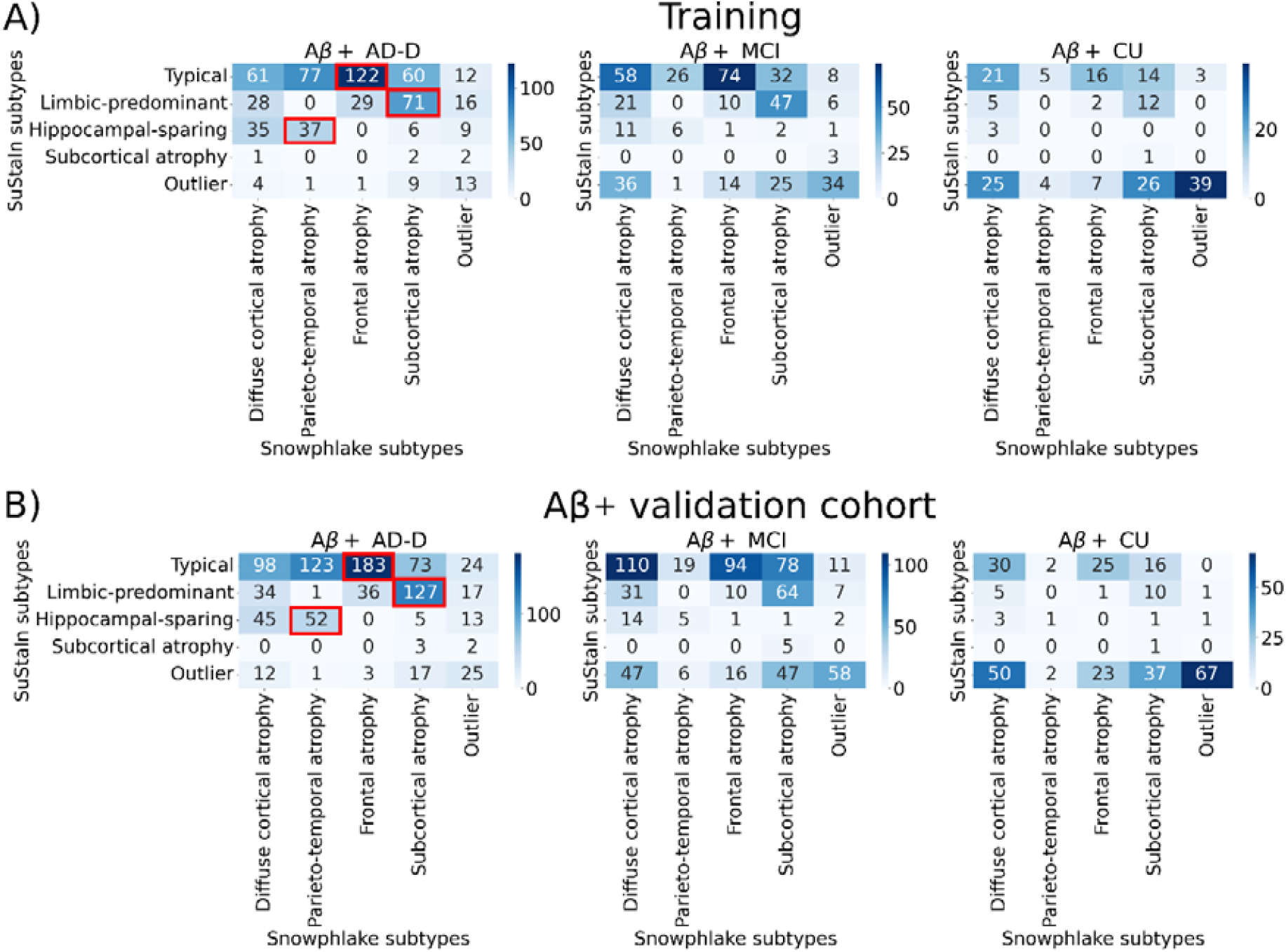
Experiment 3: Concordance of Snowphlake and SuStaIn subtypes. **A)** shows the contingency matrix of estimated atrophy-based subtypes using Snowphlake and SuStaIn for participants in the training dataset, in different clinical stages of the disease. **B)** shows a similar contingency matrices for participants in the Aβ+ validation dataset, in different clinical stages of the disease. The squares marked in red in the contingency matrix for AD-D patients correspond to the frequently co-occurring subtypes between SuStaIn and Snowphlake, also referred to as concordant subtypes. Abbreviations: CU: Cognitively unimpaired (Cognitively normal or subjective cognitive decline); MCI: Mild cognitive impairment; AD-D: Alzheimer’s disease dementia;

Of the *n* = 501 AD-D individuals assigned to the typical subtype (with prominent hippocampal and temporal lobe atrophy) of SuStaIn in the Aβ+ validation dataset, *n* = 183 (36.5%) were also assigned to the frontal atrophy subtype (with prominent frontal and temporal lobe atrophy) of Snowphlake, which is referred to as concordant subtype-pair #1. Of the *n* = 215 AD-D individuals assigned to the limbic-predominant subtype (with prominent thalamus, hippocampus, and amygdala atrophy) of SuStaIn in the Aβ+ validation dataset, *n* = 127 (59.1%) were also assigned to the subcortical-atrophy subtype of Snowphlake, which is referred to as concordant subtype-pair #2. Of the *n* = 115 AD-D individuals assigned to the hippocampal-sparing subtype of SuStaIn in the Aβ+ validation dataset, *n* = 52 (45.2%) were also assigned to the parieto-temporal atrophy subtype of Snowphlake, which is referred to as concordant subtype-pair #3. The Subcortical atrophy subtype of SuStaIn was too small to be compared. The concordant subtype-pairs accounted only for 38.6% (*n* = 230/596) of Aβ+ AD-D participants in the training dataset and 40.5% (*n* = 362/894) in the Aβ+ validation dataset. Cohort-wise contingency matrix shown in Supplementary Figure 10 further added to the evidence that low concordance was consistent across cohorts.

Average w-score maps of the concordant subtype-pairs #1 and #2 and their less frequently co-occurring counterparts showed significant differences (*p*_*FDR*_ < 0.05) within the Typical and Limbic-predominant subtypes of SuStaIn in 14 and 18 of the 24 regions respectively, adding further evidence for the spectrum of differences in atrophy within SuStaIn subtypes. Similar analysis for the concordant subtype-pairs #3 showed significant differences (*p*_*FDR*_ < 0.05) within the Hippocampal-sparing atrophy subtype of SuStaIn in 5 of the 24 regions. Supplementary Figure 11 visualizes these regional differences.

Lastly, progression modelling in the three concordant subtype-pairs using DEBM and w-score EBM showed that the estimated atrophy-event sequences using the two methods were largely similar. The normalized Kendall’s Tau (KT) metric measuring the dissimilarity between the sequences estimated by SuStaIn and Snowphlake were: *KT* = 0.11 for concordant subtype-pair #1, *KT* = 0.14 for concordant subtype-pair #2, and *KT* = 0.12 for concordant subtype-pair #3. These values are within the expected error ranges of each model,^15^ indicating that the estimated sequences in concordant subtype-pairs using the two methods agree with each other. The sequences of atrophy-events estimated using DEBM and z-score EBM are shown in Figure 6.

**Figure 6:**
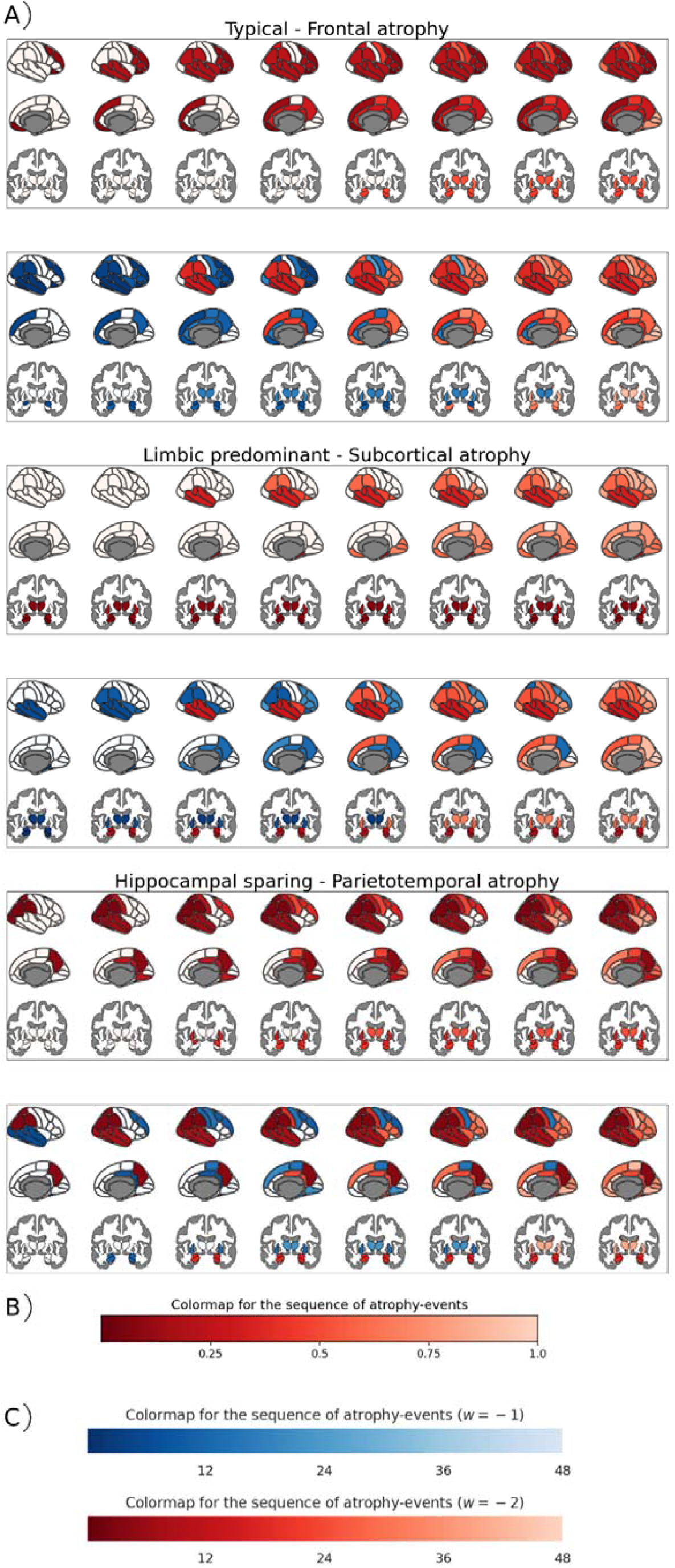
Experiment 3: Snowphlake and SuStaIn modelling of the Aβ+ participants in the three identified concordant subtypes. **A)** For each concordant subtype, the top row depicts the sequence of atrophy-events obtained using DEBM, the methodological equivalent of Snowphlake with 1-subtype. The bottom row depicts the sequence of atrophy-events obtained using w-score EBM the methodological equivalent of SuStaIn with 1-subtype. Within each subtype, the x-axis corresponds to the stage of the disease. Each column shows the brain in its lateral, medial, and subcortical views, with the regions that is expected to be abnormal at this stage. **B)** shows the scale of the colour map used for DEBM plots goes from 0 to 1, where 0 represents a region becoming abnormal at the earliest stages of the disease and 1 represents late stage. **C)** shows the scale of the colour map used for w-score EBM plots, in which regions that are expected to be mildly affected (*w* = -1) are shown in shades of blue, and severely affected (*w* = -2) in shades of red, and unaffected regions in white. The scale for the color map goes from 1 to 48, where 1 represents a region getting affected at the earliest stages of the disease and 48 represents late stage.

## Discussion

In this large-scale multi-cohort study of atrophy-heterogeneity in AD, we used a novel methodology, Snowphlake, that couples a previously-validated ML approach for disease subtyping (NMF)^4,14^ with data-driven disease progression modelling (DEBM), to estimate sequences of atrophy-events in four atrophy-based subtypes of AD. We compared our results with those obtained using SuStaIn and used the trained models to assign subtypes and atrophy stage in patient populations not included in training them. The assigned subtypes in validation datasets were associated with distinct cognitive profiles and the atrophy stage with the subtypes correlated with global cognition level of patients. We have made the trained models of both SuStaIn and Snowphlake openly available at https://snowphlake-dpm.github.io, along with the associated code. The source code for Snowphlake has also been made available at: https://github.com/snowphlake-dpm/snowphlake, while the source code for SuStaIn was previously made available by Aksman et al.^33^ A thorough comparison of Snowphlake’s subtype assignments with that of SuStaIn’s provided evidence for a spectrum of differences in atrophy among AD patients, rather than discretised by distinct subtypes.

### The identified atrophy-based subtypes were consistent with literature

Snowphlake identified a parieto-temporal atrophy subtype where the AD-D patients were consistently the youngest and had worse visuospatial function, attention and executive function consistent with prior studies on young-onset AD patients.^4,6,36^ This subtype also had a significantly lower percentage of *APOE*4 carriers in the ADC cohort, also observed in a previous study^36^, as well as in the and the ARWiBo cohort. Still, *APOE*4 carriership did not differ significantly in other cohorts in our study, which may be because those cohorts predominantly consisted of late-onset AD patients. The subcortical atrophy subtype (also referred to as “mild atrophy” in literature) patients had the least affected cognition across all domains when compared to the other subtypes.^4,9,11^ The diffuse cortical atrophy subtype (or cortical atrophy subtype) and frontal atrophy subtype have also been identified in previous studies^10,37,38^. Moreover, the subtypes identified by SuStaIn in this study (typical, hippocampal-sparing, limbic-predominant) were aligned with the neuropathological subtypes of AD reported in literature^3,39^ and largely aligned with the previous studies of atrophy-subtypes using SuStaIn.^2,10,40^

### Comparing Snowphlake and SuStaIn subtypes

A novel approach in our study was that we compared two data-driven AD subtyping techniques directly on the same patient population, while the comparisons in the previous studies so far have been based on the identified atrophy characteristics or patient characteristics.^2,4,9^ The subtypes identified by the two methods in our analysis also showed some similarities in patient characteristics, for example, parieto-temporal atrophy subtype of Snowphlake and hippocampal-sparing subtype of SuStaIn both consisted of significantly younger-onset AD-D patients. Nevertheless, our direct comparison showed low concordance between the subtype assignments of the two methods, highlighting the limitations of indirect comparisons based on read-outs.

While comparing average w-score maps of patients within a specific SuStaIn subtype, but assigned to different Snowphlake subtype, we saw significant differences in atrophy profiles, providing further evidence that atrophy patterns might vary substantially between individuals within a data-driven subtype. The three concordant subtype-pairs that accounted for approximately 40% of individuals with AD-D were the typical subtype with temporal and frontal lobe atrophy, the limbic predominant subtype with severe subcortical atrophy, and the hippocampal sparing subtype with parieto-temporal atrophy. The sequence of atrophy-events estimated by the two methods in these concordant subtype-pairs agreed with each other, showing that in spite of the methodological differences, similar inferences could be made in these concordantly subtyped individuals. Although these concordant subtype-pairs are in line with previous literature^3,5^, future work on synthetic data simulating a spectrum of atrophy differences would be crucial for understanding more about concordant subtype-pairs. However, the notion that not all patients were clustered similarly, suggests that group estimates of atrophy subtypes may be driven by a particular subset of patients, and may not capture heterogeneity of all patients. Future studies should further investigate more continuous measures of subtyping that may be able to better capture such nuance and heterogeneity.

The differences in estimated subtypes by the two methods arise from the differences in the objective-functions being optimized by the methods. While SuStaIn optimizes a non-linear objective-function to jointly estimate subtypes and atrophy-stage, Snowphlake uses linear objective-function in NMF to identify subtypes. Each of them have been shown before to identify true subtypes in the presence of distinct subtypes.^2,41^ The low concordance between the atrophy-subtype assignments of the two methods can hence be seen as evidence of a spectrum of atrophy differences between individuals with AD. This spectrum could either consist of distinct prototypical subtypes coupled with a lot of variations in a large number of AD patients, or it could be a continuum of atrophy-variations with the Snowphlake and SuStaIn identifying different variations depending on the objective-function used for their optimization. While the non-linear objective-function of SuStaIn identifies non-uniform distribution of the identified subtypes, Snowphlake’s linear objective-function identifies four subtypes that were roughly uniformly distributed. In the absence of ground-truth in data-driven subtyping, the ability of the identified subtypes to associate with distinct cognitive profiles determines their validity.

### Differences in cognitive domain profiles

The subtypes identified by Snowphlake and SuStaIn each showed significant differences in cognitive domain scores in both Aβ+ MCI and AD-D patients. While the effect sizes were comparable for Snowphlake and SuStaIn for AD-D patients, Snowphlake showed marginally stronger effect sizes for MCI patients, potentially indicating that Snowphlake’s subtypes are more sensitive at associating with different symptom profiles at the prodromal stage of the disease. While some of the differences between subtypes (by either method) assigned were consistent across the multiple cohorts in our study, we also observed significant heterogeneity in associations across cohorts. These differences could potentially indicate genuine cohort-wise differences in how atrophy causes symptoms or could be due to using different cognitive tests to compute cognitive domain scores in different cohorts. Future work on studying these associations could focus on working with harmonized cognitive data across multiple cohorts.^42,43^ Notwithstanding these inconsistencies, the significant differences in cognitive domain profiles between subtypes indicate that data-driven subtyping models have the potential to identify personalized end-points in future interventions to boost statistical power.^44,45^

### Methodological considerations and limitations

A potential limitation of our approach is that while our algorithms allow estimation of sequences of atrophy events, these remain inferences based on cross-sectional data. While there have been prior studies that validated these inferences on longitudinal datasets^46,47^, future studies could focus on a similar large-scale validation on multi-cohort longitudinal datasets to confirm if these subtypes remain consistent in preclinical and prodromal AD patients as the disease develops. One of the strengths of our study is that we have made the trained models and source code openly available and validated the subtype assignments in external datasets. Future studies can hence use these trained models to identify proteomic profiles, genetic and lifestyle factors driving these subtypes in large external cohorts. Another important feature of this study is that our combined multi-cohort data had many patients with young-onset AD-D. This could potentially be a strength of our study since young-onset AD-D patients have less comorbidity or it could be a limitation with the identified subtypes being an over-representation of young-onset AD-D patients. Lastly, by decoupling atrophy-based subtyping from disease progression modelling in the Snowphlake framework, we pave the way for the inclusion of high-dimensional imaging features (such as voxel-based measures) in data-driven subtyping and staging analysis.

## Conclusion

In conclusion, in this large-scale multi-centre study, we identified four atrophy-based subtypes using Snowphlake and SuStaIn. Subtype assignments in independent validation datasets were associated with different cognitive symptoms, and estimated atrophy-severity measures were associated with global cognition. The low concordance of subtypes between the two methods indicates that atrophy differences between individuals may be a spectrum rather than strictly delineated subtypes. Based on our findings, future research should prioritize developing novel approaches to capture and analyse this spectrum of heterogeneity in atrophy patterns to help us further understand the biological-basis for the observed variability in atrophy patterns between individuals.

## Data Availability

The ADNI data used is this study were obtained from the ADNI database (adni.loni.usc.edu). The ADC data used in this study are available from the corresponding author, upon reasonable request. The AIBL imaging data used in this study were obtained from the AIBL LONI database (https://ida.loni.usc.edu/login.jsp?project=AIBL), while cognitive and genetic data can be requested from the AIBL management team, upon reasonable request by submitting an Expression of Interest (EOI) form available on the AIBL website (https://aibl.org.au/collaboration/). The NACC data used in this study were obtained from https://naccdata.org/. The OASIS data used in this study were obtained from https://sites.wustl.edu/oasisbrains/website. The data of the other cohorts used in this study can be requested from the neuGRID (https://www.neugrid2.eu/) and GAAIN (https://www.gaain.org) platforms after registration.

## Acknowledgments

The authors would like to thank all the research participants and their families for donating their data for scientific research.

Data collection and sharing for ADNI was funded by the Alzheimer’s Disease Neuroimaging Initiative (ADNI) (National Institutes of Health Grant U01 AG024904) and DOD ADNI (Department of Defense award number W81XWH-12-2-0012). ADNI is funded by the National Institute on Aging, the National Institute of Biomedical Imaging and Bioengineering, and through generous contributions from the following: AbbVie, Alzheimer’s Association; Alzheimer’s Drug Discovery Foundation; Araclon Biotech; BioClinica, Inc.; Biogen; Bristol-Myers Squibb Company; CereSpir, Inc.; Cogstate; Eisai Inc.; Elan Pharmaceuticals, Inc.; Eli Lilly and Company; EuroImmun; F. Hoffmann-La Roche Ltd and its affiliated company Genentech, Inc.; Fujirebio; GE Healthcare; IXICO Ltd.; Janssen Alzheimer Immunotherapy Research & Development, LLC.; Johnson & Johnson Pharmaceutical Research & Development LLC.; Lumosity; Lundbeck; Merck & Co., Inc.; Meso Scale Diagnostics, LLC.; NeuroRx Research; Neurotrack Technologies; Novartis Pharmaceuticals Corporation; Pfizer Inc.; Piramal Imaging; Servier; Takeda Pharmaceutical Company; and Transition Therapeutics. The Canadian Institutes of Health Research is providing funds to support ADNI clinical sites in Canada. Private sector contributions are facilitated by the Foundation for the National Institutes of Health (www.fnih.org). The grantee organization is the Northern California Institute for Research and Education, and the study is coordinated by the Alzheimer’s Therapeutic Research Institute at the University of Southern California. ADNI data are disseminated by the Laboratory for Neuro Imaging at the University of Southern California.

The AIBL study (https://aibl.org.au) is a consortium between Austin Health, CSIRO, Edith Cowan University, the Florey Institute (The University of Melbourne), and the National Ageing Research Institute. The study has received partial financial support from the Alzheimer’s Association (US), the Alzheimer’s Drug Discovery Foundation, an Anonymous foundation, the Science and Industry Endowment Fund, the Dementia Collaborative Research Centres, the Victorian Government’s Operational Infrastructure Support program, the Australian Alzheimer’s Research Foundation, the National Health and Medical Research Council (NHMRC), and The Yulgilbar Foundation. Numerous commercial interactions have supported data collection and analyses. In-kind support has also been provided by Sir Charles Gairdner Hospital, Cogstate Ltd, Hollywood Private Hospital, The University of Melbourne, and St Vincent’s Hospital. The AIBL team wishes to thank all clinicians who referred patients with AD and/or MCI to the study. We also thank all those who took part as subjects in the study for their commitment and dedication to helping advance research into the early detection and causation of AD. We thank all the investigators within the AIBL who contributed to the design and implementation of the resource and/or provided data but did not actively participate in the development, analysis, interpretation or writing of this current study.

The NACC database is funded by NIA/NIH Grant U24 AG072122. NACC data are contributed by the NIA-funded ADRCs: P30 AG062429 (PI James Brewer, MD, PhD), P30 AG066468 (PI Oscar Lopez, MD), P30 AG062421 (PI Bradley Hyman, MD, PhD), P30 AG066509 (PI Thomas Grabowski, MD), P30 AG066514 (PI Mary Sano, PhD), P30 AG066530 (PI Helena Chui, MD), P30 AG066507 (PI Marilyn Albert, PhD), P30 AG066444 (PI John Morris, MD), P30 AG066518 (PI Jeffrey Kaye, MD), P30 AG066512 (PI Thomas Wisniewski, MD), P30 AG066462 (PI Scott Small, MD), P30 AG072979 (PI David Wolk, MD), P30 AG072972 (PI Charles DeCarli, MD), P30 AG072976 (PI Andrew Saykin, PsyD), P30 AG072975 (PI David Bennett, MD), P30 AG072978 (PI Neil Kowall, MD), P30 AG072977 (PI Robert Vassar, PhD), P30 AG066519 (PI Frank LaFerla, PhD), P30 AG062677 (PI Ronald Petersen, MD, PhD), P30 AG079280 (PI Eric Reiman, MD), P30 AG062422 (PI Gil Rabinovici, MD), P30 AG066511 (PI Allan Levey, MD, PhD), P30 AG072946 (PI Linda Van Eldik, PhD), P30 AG062715 (PI Sanjay Asthana, MD, FRCP), P30 AG072973 (PI Russell Swerdlow, MD), P30 AG066506 (PI Todd Golde, MD, PhD), P30 AG066508 (PI Stephen Strittmatter, MD, PhD), P30 AG066515 (PI Victor Henderson, MD, MS), P30 AG072947 (PI Suzanne Craft, PhD), P30 AG072931 (PI Henry Paulson, MD, PhD), P30 AG066546 (PI Sudha Seshadri, MD), P20 AG068024 (PI Erik Roberson, MD, PhD), P20 AG068053 (PI Justin Miller, PhD), P20 AG068077 (PI Gary Rosenberg, MD), P20 AG068082 (PI Angela Jefferson, PhD), P30 AG072958 (PI Heather Whitson, MD), P30 AG072959 (PI James Leverenz, MD).

## Supplementary material

Supplementary material is available online.

