## Supplementary material for "A large-scale multi-centre study characterising atrophy heterogeneity in Alzheimer’s disease"

### S1. Supplementary methods

#### S1.1 ADNI

The ADNI was launched in 2003 as a public-private partnership, led by Principal Investigator Michael W. Weiner, MD. The primary goal of ADNI has been to test whether serial magnetic resonance imaging (MRI), positron emission tomography (PET), other biological markers, and clinical and neuropsychological assessment can be combined to measure the progression of mild cognitive impairment (MCI) and early Alzheimer's disease (AD).

#### S1.2 Selecting thresholds for CSF Aβ

In ADC, $A\beta_{1-42}$ concentrations in CSF were statistically corrected for time-dependant drifts in values. Gaussian mixture modeling (GMM) was used to fit a bimodal Gaussian distribution in the resulting data, to identify cut-off points for identifying Aβ positivity.[1] This resulted in a cut-off point of 813 for Innotest assay. Apart from this, a cut-off point of 1089 was used for CSF $A\beta_{1-42}$ concentrations derived from Elecsys assay.[2]

In ADNI, $A\beta_{1-42}$ concentrations obtained from INNO BIA AlzBIO3 xMAP multiplex immunoassay[3] were used to fit a GMM with a bimodal Gaussian distribution. A cut-off point of 1029 was identified based on this analysis. The cohorts of ARWiBo, EDSD, and PharmaCog used ELISA assay, where the $A\beta_{1-42}$ concentrations were rescaled to match the distribution of ADNI subjects.[4] The aforementioned cut-off point of ADNI was then used in these cohorts as well.

In the NACC cohort, the CSF assay methods were not standardized across the AD research centers, potentially resulting in different cutoffs for each center. The thresholded CSF status was provided in the uniform data set made available for this study.

#### S1.3 Aβ positivity in PET scans

In ADC[5] and GMC[6] cohorts, Aβ status in PET images were decided based on radiological readouts. In ADNI and AIBL cohorts, centiloid values were computed for PET images with multiple tracers using capAIBL pipeline[7]. PET images in ADNI used either florbetapir (AV45), Pittsburgh Compound-B (PiB), or florbetaben (FBB) tracer, while those in AIBL used either AV45, PIB, or flutemetamol (FLUTE) tracer. Centiloid-based positivity threshold was chosen as 30 as reported in *Salvado et al.*[8]

In the NACC cohort, the PET tracers and evaluation methods were not standardized across the AD research centers. The thresholded PET Aβ status was provided in the uniform data set made available for this study.

#### S1.4 Cognitive domain score computation

**Memory**: In ADC, the domain memory included the immediate and delayed recall of the Dutch version of the Rey Auditory Verbal Learning Task (RAVLT),[9] part B of the Visual Association Test (VAT),[10] and delayed recall of the Rey Complex Figure Test (RCFT).[11] For AIBL and ADNI, pre-computed domain scores were used and have been described in detail elsewhere.[12,13] In the NACC cohort, memory domain scores were computed independently for pre-2015 participants and post-2015 participants, since the uniform data set (UDS) and the corresponding cognitive test battery were updated from version 2 (v2) to version 3 (v3), in 2015.[14] The memory domain in v2 included immediate and delayed story recall, whereas in v3 it included immediate and delayed craft story (CS) recall.[15] In the GMC cohort, the domain included immediate and delayed story recollection test, three-objects three-places test,[16] and doors test.[17]

**Attention and Executive function**: In ADC, this domain included the Trail Making Test (TMT) part A and B,[18] the forward and backward condition of digit span,[19] and the Stroop test,[20] the letter fluency test[21], and letter digit substitution test (LDST).[22] For ADNI, pre-computed domain scores were used and have been described in detail elsewhere.[23] In the AIBL cohort, this domain included digit symbol substitution test,[19] the forward and backward condition of digit span,[19] controlled oral word association test,[24] and Category switching test.[25] In the NACC cohort, the domain score for v2 included the forward and backward condition of digit span, WAIS-R digit symbol test,[19] and TMT part A and B. The domain score in v3 included the forward and backward condition of digit span, letter fluency test, and TMT part A and B. In the GMC cohort, the domain score included WAIS-R digit symbol test, TMT part A and B, letter fluency test, the forward and backward condition of digit span, and Victoria stroop test was used.[26]

**Visuospatial function**: In ADC, this domain included Rey–Osterrieth Complex Figure test[27] and two subtests of the visual object and space perception battery: number location, and fragmented letters.[28] For AIBL and ADNI, pre-computed domain scores were used and have been described in detail elsewhere.[29,30] In the NACC cohort, the domain score was not computed for v2 due to the absence of relevant tests in the UDS, whereas for v3 it was computed based on total score for copy of Benson figure test.[31] In the GMC cohort, the domain score was based on clock drawing test.[32]

**Language**: In ADC, the domain language included the category fluency test (animal naming)[33] and the naming condition of the VAT. For AIBL and ADNI, pre-computed domain scores were used and have been described in detail elsewhere.[29,30] In the NACC cohort, the domain score for both v2 and v3 consisted of category fluency test (naming animals and vegetables). Additionally, v2 used total score in Boston naming test,[34] while v3 used total score in Multilingual naming test.[35] The domain score was not computed in the GMC cohort due to the absence of relevant tests.

#### S1.5 Training the discriminative event-based model (DEBM)

First, we performed Gaussian mixture modelling (GMM) with the Aβ− CU group as reference, to derive cut-off points for volumes in each region to be termed "abnormal" in a data-driven way. This was done in a semi-coupled manner for the different subtypes as described in Venkatraghavan *et al. as* co-initialized GMM.[36] Secondly, using the results of GMM, we estimated posterior probabilities for each region to be abnormal for each Aβ+ participant within each subtype. This in turn was used to approximate the atrophy progression pattern for each Aβ+ participant ($t_{ij}$, where $i$ refers to one of the $k_{opt}$ subtypes, $j$ refers to each individual within the subtype). Lastly, these individual estimates were combined to robustly estimate the atrophy progression pattern in each subtype ($T_i$). To estimate the uncertainty of this inferred progression pattern, we performed 100 iterations of bootstrap resampling while keeping the subtype embeddings fixed. This ensured that the definitions of the data-driven subtypes remained consisted across the 100 random bootstrap resampling iterations.

#### S1.6 SuStaIn

We used w-score volumetric data of Aβ+ participants across all clinical stages to train the SuStaIn model. The sequence of atrophy-events were described using piecewise linear w-score model, with $w = -1$ and $w = -2$ chosen as event thresholds, computed by considering Aβ− CU participants as reference. The optimum number of subtypes, potentially up to a maximum of 5 subtypes, were estimated using a cross-validation information criteria (CVIC)[37] using 3-fold cross-validation strategy. To get a probabilistic estimate of the likely sequence of atrophy-events in each of the estimated subtypes, Markov chain Monte Carlo (MCMC) sampling technique was used to sample and evaluate $1 \times 10^6$ potential candidates of sequences. These sequences were used to estimate the atrophy severity of individuals, as measured by the maximum likelihood position along the sequence of events for each subtype, independently for each individual. This approach was used to predict the subtypes and severity of all the participants in the internal as well as in the external cohorts.

### S2. Supplementary Figures and Tables

**Supplementary Table 1:** Cohort characteristics.

| Cohort | Data collection period included in this study | Setting | Inclusion criteria | Participant amyloid status derived from |
|---|---|---|---|---|
| ADC | 1998 - 2020 | Tertiary memory clinic | Referred to the memory clinic and diagnosed with AD-D, MCI, or SCD in clinic. | CSF, PET visual scores |
| ADNI | 2004 - 2021 | Research | Participants in the substudies of ADNI 1, ADNI Go, ADNI 2, and ADNI 3 were included. The inclusion criteria for these studies and the differences between these substudies are detailed by R. Peterson et al.,[38] M.W. Weiner et al.,[39,40] | CSF, PET centiloid scores |
| AIBL | 2007 - 2014 | Research | Participant data that was made publicly available on LONI website ida.loni.usc.edu were included. The inclusion criteria for AIBL have been detailed by C. Fowler et al.[12] | PET centiloid scores* |
| ARWiBo | 2001 – 2013 | Memory clinic | Referred to the memory clinic, diagnosed with AD-D, MCI, SCD or CN, without other psychiatric or neurological condition. | CSF |
| EDSD | 2010 – 2016 | Research | Participants were enrolled in 11 centers across Europe. The inclusion criteria for EDSD were detailed by Brueggen et al.[41] | CSF |
| I-ADNI | 2009 - 2011 | Research | Participants were enrolled in nine Italain memory clinics. The inclusion criteria for I-ADNI has been detailed by Cavedo et al.[42] | NA |
| NACC | 2005 – 2022 | Multiple memory clinics | Each center enrolls its participants according to its own protocol — e.g., clinician referral, self-referral by participants or family members, active recruitment in community organizations. | CSF, PET†, Neuropathology |
| OASIS | NA | Research | Participants in the substudies OASIS-1 and OASIS-2 ere included. The Inclusion Criteria for the substudies were detailed in Marcus et al.[43] | NA |
| PharmaCog | 2011 - 2015 | Research | Participants with MCI were longitudinally follow-ed up across 13 European research centers. | CSF |
| GMC | 2007 - 2021 | Memory clinic | Referred to the memory clinic, diagnosed with AD-D, MCI, SCD or CN, without other psychiatric or neurological condition. | PET visual scores |

* CSF biomarkers are available in the AIBL study, but were not used in our work.
† PET quantification varied across the different AD research centers that are part of NACC.
Abbreviations: SCD: Subjective cognitive decline; MCI: Mild cognitive impairment; AD-D: Alzheimer's disease dementia; NA: Not available; CSF: cerebrospinal fluid; PET: positron emission tomography.

**Supplementary Table 2:** Scanners considered in this study and the sex distribution of the participants scanned in them.

| Manufacturer | Scanner Model | Magnetic Field (T) | Number of scans (Female %) |
|---|---|---|---|
| Canon | Titan | 3.0 | 571 (43.6%) |
| GE | Discovery MR750 | 3.0 | 673 (49.9%) |
| | Discovery MR750w | 3.0 | 23 (34.8%) |
| | Genesis Signa | 1.5 | 514 (61.9%) |
| | Signa Excite | 1.5 | 362 (47.2%) |
| | Signa PET/MR | 3.0 | 33 (48.4%) |
| | Signa HDx | 1.5 | 30 (40.0%) |
| | Signa HDx | 3.0 | 96 (42.7%) |
| | Signa HDxt | 1.5 | 473 (45.5%) |
| | Signa HDxt | 3.0 | 981 (46.5%) |
| | Signa Premier | 3.0 | 10 (50.0%) |
| Philips | Achieva | 1.5 | 11 (36.3%) |
| | Achieva | 3.0 | 242 (61.6%) |
| | Achieva dStream | 3.0 | 25 (60.0%) |
| | Eclipse | 1.5 | 43 (69.8%) |
| | Gemini | 3.0 | 9 (66.7%) |
| | Gyroscan NT | 1.0 | 173 (63.6%) |
| | Ingenia | 1.5 | 18 (55.6%) |
| | Ingenia | 3.0 | 64 (39.0%) |
| | Ingenuity | 3.0 | 647 (46.8%) |
| | Intera | 1.0 | 409 (61.6%) |
| | Intera | 1.5 | 59 (28.8%) |
| | Intera | 3.0 | 50 (54.0%) |
| | Intera Achieva | 1.5 | 5 (20.0%) |
| | Intera Gyroscan | 1.5 | 27 (40.7%) |
| Siemens | Aera | 1.5 | 15 (53.3%) |
| | Allegra | 3.0 | 84 (59.5%) |
| | Avanto | 1.5 | 294 (50.0%) |
| | Biograph | 3.0 | 5 (0.0%) |
| | Espree | 1.5 | 7 (42.9%) |
| | Magnetom Expert | 1.0 | 765 (48.8%) |
| | Magnetom Impact | 1.0 | 9 (77.8%) |
| | Magnetom Vida | 3.0 | 19 (31.6%) |
| | Magnetom Vision | 1.5 | 26 (76.9%) |
| | Prisma | 3.0 | 143 (52.4%) |
| | Prima fit | 3.0 | 169 (56.8%) |
| | Skyra | 3.0 | 847 (59.4%) |
| | Sonata | 1.5 | 403 (46.4%) |
| | Sonata Vision | 1.5 | 5 (60.0%) |
| | Symphony | 1.5 | 148 (59.5%) |
| | Trio | 3.0 | 67 (65.7%) |
| | Trio Tim | 3.0 | 837 (53.5%) |
| | Verio | 3.0 | 318 (57.5%) |
| | Vision | 1.5 | 302 (65.2%) |

**Supplementary Table 3:** Mapping to derive the composite cortical ROIs from Freesurfer cortical parcellation

| Composite ROI | Freesurfer cortical parcellation |
|---|---|
| Temporal lobe | bankssts, transversetemporal, superiortemporal, temporalpole, entorhinal, middletemporal, inferiortemporal, fusiform |
| Superior frontal gyrus | superiorfrontal, frontalpole |
| Middle frontal gyrus | caudalmiddlefrontal, rostralmiddlefrontal |
| Inferior frontal gyrus | parsopercularis, parsorbitalis, parstriangularis |
| Gyrus rectus | medialorbitofrontal |
| Orbitofrontal gyri | lateralorbitofrontal |
| Precentral gyrus | precentral, paracentral |
| Postcentral gyrus | postcentral |
| Superior parietal gyrus | superiorparietal, precuneus |
| Inferolateral remainder of parietal lobe | inferiorparietal, supramarginal |
| Lateral remainder of occipital lobe | lateraloccipital |
| Cuneus | cuneus, pericalcarine |
| Lingual gyrus | lingual |
| Insula | insula |
| Gyrus cinguli anterior part | caudalanteriorcingulate, rostralanteriorcingulate |
| Gyrus cinguli posterior part | posteriorcingulate, isthmuscingulate |
| Parahippocampal and ambient gyri | parahippocampal |

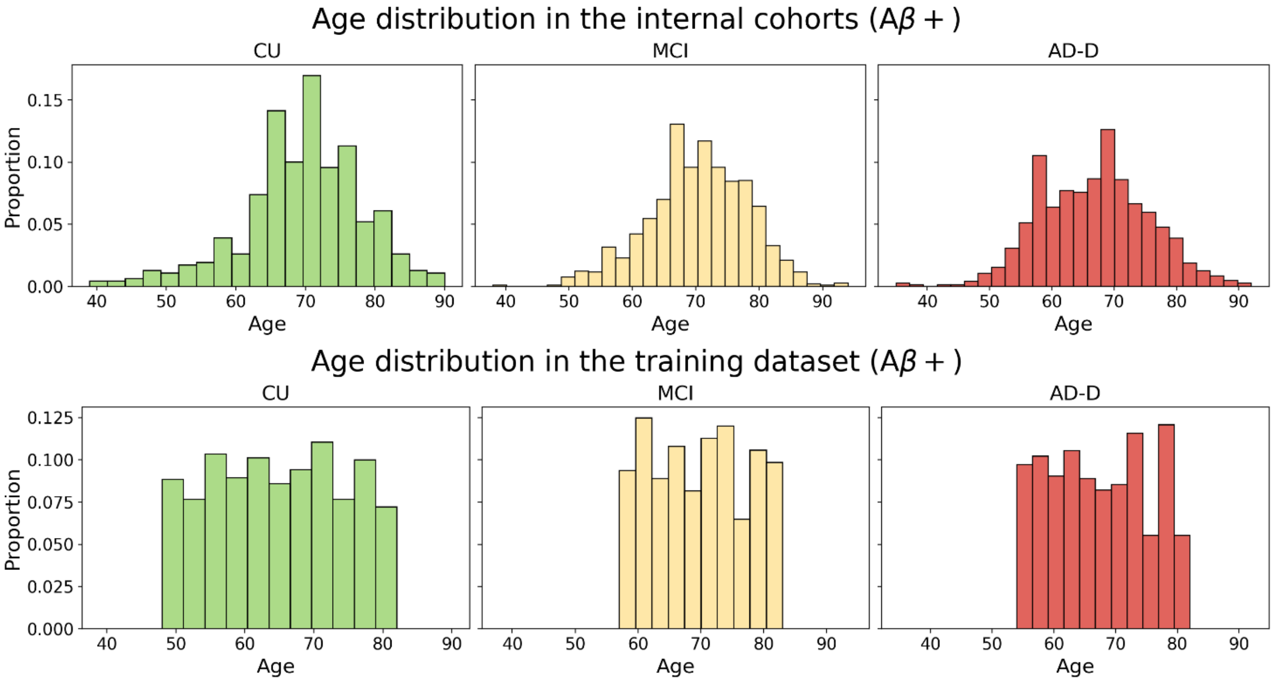


**Supplementary Figure 1:** Normalized histograms depicting the age distribution of Aβ+ participants. The top row shows the age distribution of the Aβ+ participants in all the cohorts except the GMC cohort (external). The bottom row shows the age distribution of the participants in the training dataset, selected based on weighted random sampling without replacement. Abbreviations: CU = Cognitively unimpaired; MCI = Mild cognitively impaired; AD-D = Alzheimer's disease dementia.

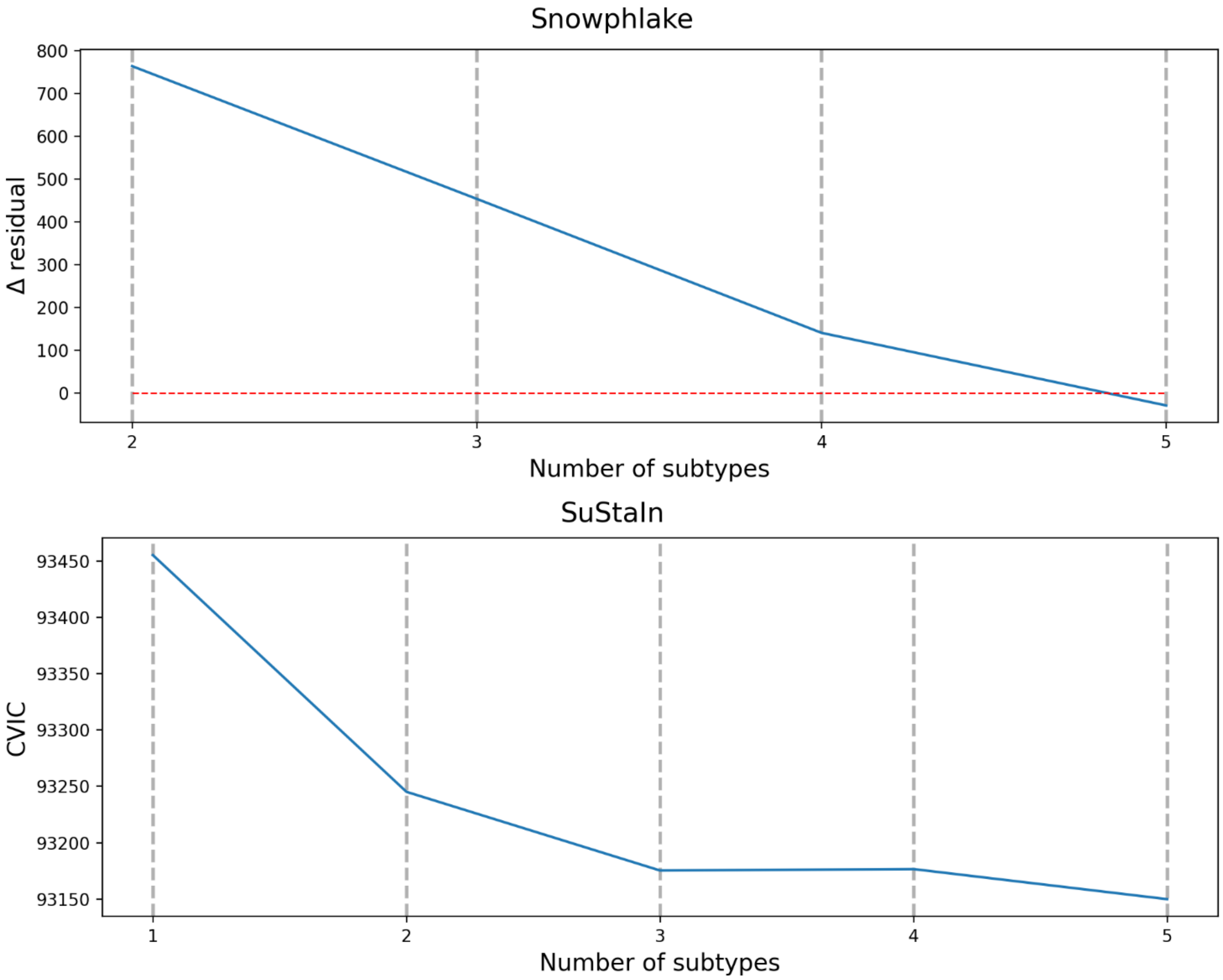


**Supplementary Figure 2**: Criteria for selecting the optimum number of subtypes for Snowphlake (top) and SuStaIn (bottom). For Snowphlake, the optimum number of subtypes chosen should ideally by higher than the red-dotted line.

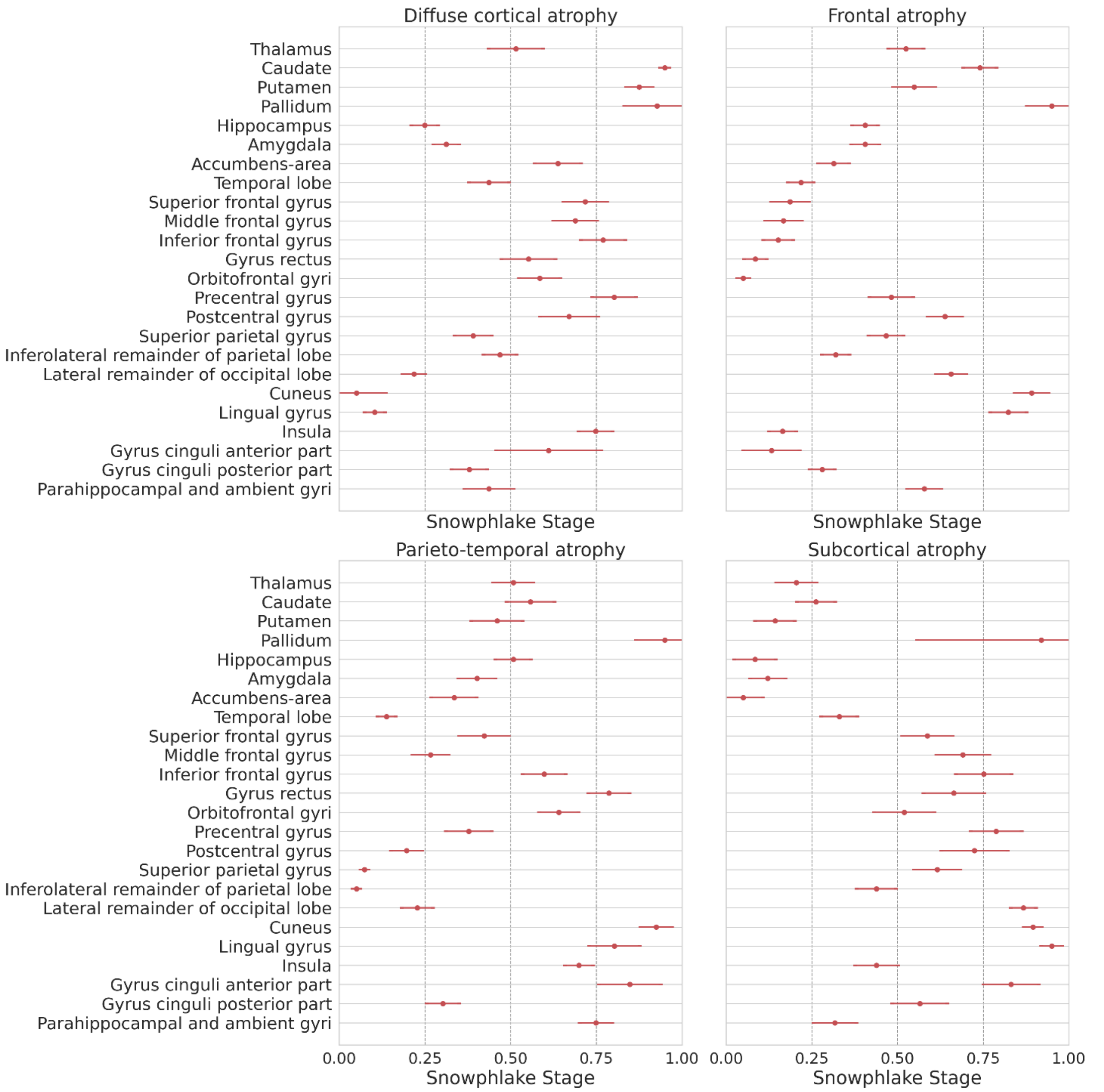


**Supplementary Figure 3:** Snowphlake event-centre variance diagram, depicting uncertainty in the atrophy-events estimated in Aβ+ participants in the training dataset. The event-centres of the impacted brain regions are indicated as points on a normalized staging-scale between 0 and 1, along with their standard errors represented by horizontal bars. Standard errors were estimated from a set of 100 independent bootstrap re-sampling of the training dataset.

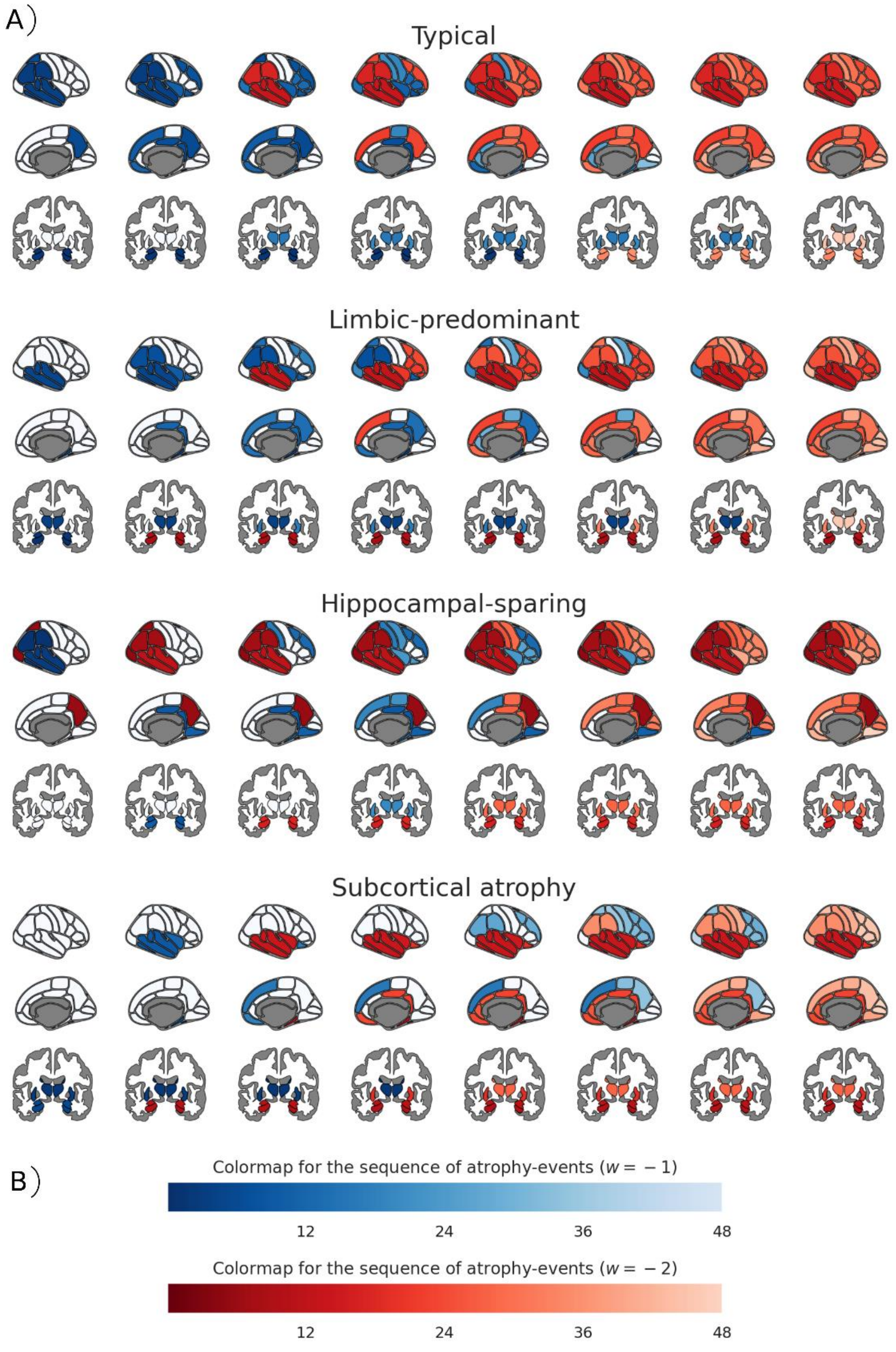


**Supplementary Figure 4:** SuStaIn modelling in the Aβ+ participants in the multi-cohort training dataset. These plots depicts the subtypes and sequence of atrophy-events for each subtype estimated. Within each subtype, the x-axis corresponds to the stage of the disease. Each column shows the brain in its lateral, medial, and subcortical views, with the regions that is expected to be mildly affected ($w = -1$) at this stage for the subtype in shades of blue, severely affected ($w = -2$) in shades of red, and unaffected regions in white. The scale for the color map goes from 1 to 48, the staging scale for SuStaIn, where 1 represents a region getting affected at the earliest stages of the disease and 48 represents late stage.

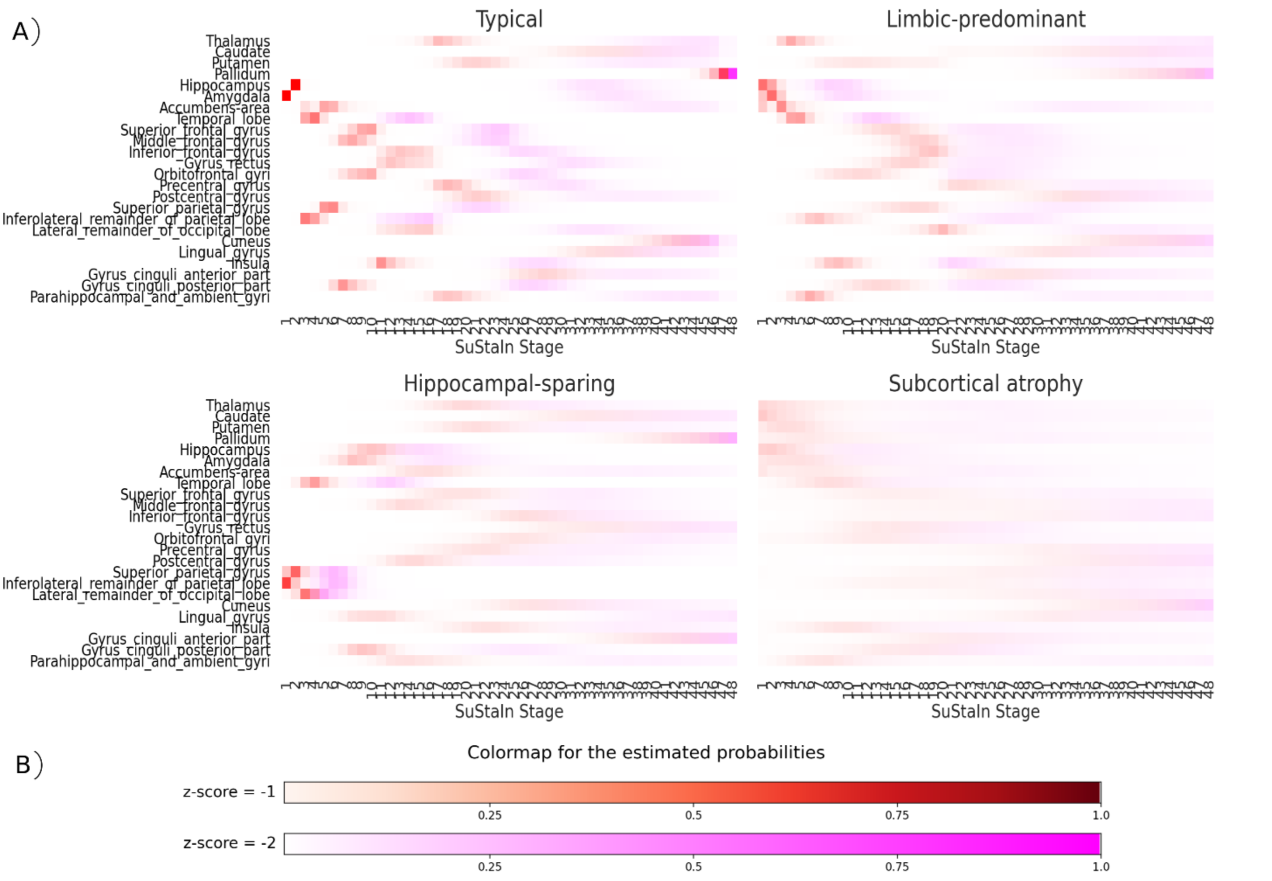


**Supplementary Figure 5**: SuStaIn Positional Variance Diagrams (PVDs) estimated using MCMC simulations. Each row corresponds to a region and each coloured entry in a PVD marks the probability that the biomarker has surpassed a w-score threshold. The colour scales used for $w = -1$ and $w = -2$ are shown in (B). While a higher w-score reflects a more severe abnormality of the biomarker, a low positional variance, which is marked in a clearer colour, is associated with a higher degree of certainty.

**Supplementary Table 4**: Characteristics of atrophy-based subtypes assigned by the trained Snowphlake model, in the internal and external validation cohorts. * indicates the corresponding measure is significantly different ($p < 0.05$) between the different subtypes (excluding the outliers group) in Aβ+ validation cohort, using ANOVA test for Age and MMSE characteristics, and $\chi^2$ contingency test for *APOE4* characteristics. # indicates the significant difference ($p < 0.05$) using similar tests in the clinical validation cohort. Abbreviations: CU: Cognitively unimpaired (Cognitively normal or subjective cognitive decline); MCI: Mild cognitive impairment; AD-D: Alzheimer's disease dementia; NA: Not applicable.

| | | Diffuse cortical atrophy | | Parieto-temporal atrophy | | Frontal atrophy | | Subcortical atrophy | | Outliers | |
|---|---|---|---|---|---|---|---|---|---|---|---|
| | | Aβ+ | All | Aβ+ | All | Aβ+ | All | Aβ+ | All | Aβ+ | All |
| **Characteristic per diagnostic group** | | **Validation Cohort: ADC** $n = 937$ (Aβ+); $n = 2{,}312$ | | | | | | | | | |
| Age | CU# | 63.3±9.6 | 59.9±10.7 | 73.0±1.4 | 65.0±11.1 | 63.2±9.6 | 63.6±9.9 | 67.6±7.2 | 62.6±10.7 | 61.5±10.8 | 59.3±10.5 |
| | MCI*# | 68.2±6.1 | 67.8±7.3 | 60.9±8.1 | 63.0±7.5 | 67.5±8.0 | 67.4±8.5 | 66.3±8.1 | 66.0±8.4 | 66.1±10.2 | 65.3±7.9 |
| | AD-D*# | 64.4±7.2 | 66.3±7.9 | 60.2±7.4 | 60.9±7.6 | 66.1±7.8 | 68.6±8.3 | 65.7±7.1 | 67.3±7.6 | 62.6±8.9 | 66.6±9.8 |
| N (%) | CU | 27 (27.0%) | 176 (31.7%) | 2 (2.0%) | 15 (2.7%) | 21 (21.0%) | 109 (19.6%) | 32 (32.0%) | 141 (25.4%) | 18 (18.0%) | 115 (20.7%) |
| | MCI | 73 (35.8%) | 180 (27.3%) | 8 (3.9%) | 22 (3.3%) | 41 (20.1%) | 158 (24.0%) | 68 (33.3%) | 213 (32.3%) | 14 (6.9%) | 86 (13.1%) |
| | AD-D | 122 (19.3%) | 215 (19.6%) | 141 (22.3%) | 195 (17.8%) | 170 (26.9%) | 342 (31.2%) | 151 (23.9%) | 246 (22.4%) | 49 (7.7%) | 99 (9.0%) |
| Sex ($n_{male}$/ $n_{female}$) | CU | 11/16 | 102/74 | 0/2 | 9/6 | 13/8 | 66/43 | 21/11 | 86/55 | 3/15 | 27/88 |
| | MCI# | 32/41 | 105/75 | 6/2 | 18/4 | 27/14 | 115/43 | 36/32 | 137/76 | 6/8 | 40/46 |
| | AD-D*# | 37/85 | 83/132 | 67/74 | 92/103 | 78/92 | 166/176 | 77/74 | 126/120 | 11/38 | 35/64 |
| MMSE | CU | 27.8±1.5 | 28.2±1.7 | 27.5±0.7 | 27.9±1.0 | 28.6±1.3 | 28.1±1.8 | 28.0±1.5 | 28.4±1.4 | 28.2±1.4 | 28.2±2.0 |
| | MCI | 26.7±2.5 | 26.6±2.5 | 24.6±1.8 | 25.8±2.3 | 26.5±2.0 | 26.3±2.5 | 26.2±2.2 | 26.3±2.4 | 27.0±2.0 | 26.9±1.9 |
| | AD-D*# | 21.4±4.8 | 21.0±5.1 | 18.7±5.3 | 18.5±5.5 | 20.8±4.9 | 20.4±5.3 | 21.8±4.4 | 21.6±4.6 | 19.8±6.0 | 20.1±5.6 |
| APOE4 carriers ($n_{APOE}$/ $n_{total}$) | CU | 20/27 | 54/176 | 1/2 | 4/15 | 10/21 | 30/109 | 20/32 | 32/141 | 10/18 | 27/115 |
| | MCI | 48/73 | 74/180 | 5/8 | 11/22 | 30/41 | 68/158 | 47/68 | 94/213 | 10/14 | 21/86 |
| | AD-D*# | 93/122 | 129/215 | 78/141 | 88/195 | 110/170 | 165/342 | 108/151 | 151/246 | 30/49 | 44/99 |
| **Characteristic per diagnostic group** | | **Validation Cohort: ADNI** $n = 465$ (Aβ+); $n = 1{,}098$ | | | | | | | | | |
| | | Diffuse cortical atrophy | | Parieto-temporal atrophy | | Frontal atrophy | | Subcortical atrophy | | Outliers | |
| Age | CU | 74.8±6.0 | 74.8±5.8 | 74.0±5.3 | 74.4±4.1 | 74.8±6.5 | 75.3±5.7 | 71.8±4.1 | 73.5±4.9 | 71.2±5.8 | 72.1±5.3 |
| | MCI | 71.2±7.1 | 71.9±7.4 | 71.8±6.9 | 70.5±7.4 | 73.0±5.4 | 71.8±7.1 | 73.3±5.8 | 73.3±6.8 | 70.9±7.2 | 69.6±7.8 |
| | AD-D*# | 76.3±8.5 | 76.0±7.7 | 67.6±9.6 | 68.6±8.1 | 71.2±8.8 | 73.2±7.3 | 72.2±6.8 | 74.0±7.3 | 75.2±8.7 | 73.6±8.4 |
| N (%) | CU | 40 (34.8%) | 74 (32.5%) | 3 (2.6%) | 7 (3.1%) | 18 (15.7%) | 45 (19.7%) | 26 (22.6%) | 52 (22.8%) | 28 (24.3%) | 50 (21.9%) |
| | MCI | 73 (30.4%) | 207 (30.8%) | 13 (5.4%) | 40 (6.0%) | 50 (20.8%) | 144 (21.5%) | 69 (28.8%) | 178 (26.5%) | 35 (14.6%) | 102 (15.2%) |
| | AD-D | 35 (31.8%) | 69 (34.7%) | 14 (12.7%) | 21 (10.6%) | 12 (10.9%) | 31 (15.6%) | 36 (32.7%) | 53 (26.6%) | 13 (11.8%) | 25 (12.6%) |
| Sex ($n_{male}$/ $n_{female}$) | CU | 22/18 | 41/33 | 2/1 | 3/4 | 11/7 | 30/15 | 12/14 | 24/28 | 5/23 | 14/36 |
| | MCI# | 41/32 | 102/105 | 7/6 | 24/16 | 33/17 | 98/46 | 46/23 | 117/61 | 17/18 | 49/53 |
| | AD-D | 17/18 | 32/37 | 6/8 | 10/11 | 7/5 | 19/12 | 17/19 | 28/25 | 6/7 | 10/15 |
| MMSE | CU | 29.2±1.0 | 29.0±1.1 | 29.0±1.0 | 29.1±0.7 | 29.0±1.2 | 29.2±1.0 | 29.2±1.0 | 29.2±0.9 | 29.3±1.1 | 29.3±1.0 |
| | MCI | 27.5±2.2 | 27.7±1.9 | 26.9±1.8 | 27.4±1.8 | 27.4±2.0 | 27.5±1.8 | 27.4±1.7 | 27.7±1.8 | 27.9±2.0 | 28.0±1.8 |
| | AD-D | 23.2±2.1 | 23.0±2.1 | 23.0±2.4 | 23.3±2.4 | 24.0±2.0 | 23.3±2.1 | 23.5±2.5 | 23.5±2.3 | 22.2±2.7 | 22.7±2.5 |
| APOE4 carriers ($n_{APOE}$/ $n_{total}$) | CU | 17/40 | 27/73 | 2/3 | 4/7 | 9/18 | 16/44 | 13/26 | 16/51 | 17/28 | 23/48 |
| | MCI | 45/69 | 99/201 | 11/13 | 21/40 | 31/48 | 66/138 | 42/68 | 79/174 | 23/33 | 40/94 |
| | AD-D | 27/35 | 48/68 | 11/14 | 17/21 | 10/12 | 19/31 | 32/36 | 41/52 | 11/13 | 17/25 |
| | | **Validation Cohort: AIBL** $n = 111$ (Aβ+); $n = 182$ | | | | | | | | | |

| Characteristic per diagnostic group | | Diffuse cortical atrophy | | Parieto-temporal atrophy | | Frontal atrophy | | Subcortical atrophy | | Outliers | |
|---|---|---|---|---|---|---|---|---|---|---|---|
| Age | CU | 77.1±6.3 | 76.2±5.9 | NA | NA | 77.6±7.3 | 77.0±7.3 | 72.1±6.4 | 73.4±6.3 | 73.7±6.9 | 72.7±6.7 |
| | MCI | 76.8±8.1 | 74.1±7.9 | 75.0±6.2 | 73.5±5.9 | 74.5±4.9 | 69.7±6.8 | 75.6±6.2 | 76.0±5.8 | 74.2±4.4 | 76.5±8.5 |
| | AD-D*# | 80.7±7.5 | 79.8±5.4 | 60.3±3.2 | 60.3±3.2 | 67.0±7.7 | 71.8±9.8 | 75.0±6.8 | 75.8±6.1 | 73.0 | 73.0±7.3 |
| N (%) | CU | 21 (34.4%) | 29 (32.6%) | 0 (0.0%) | 0 (0.0%) | 10 (16.4%) | 14 (15.7%) | 7 (11.5%) | 11 (12.4%) | 23 (37.7%) | 35 (39.3%) |
| | MCI | 9 (27.3%) | 18 (26.9%) | 3 (9.1%) | 4 (6.0%) | 2 (6.1%) | 9 (13.4%) | 14 (42.4%) | 25 (37.3%) | 5 (15.2%) | 11 (16.4%) |
| | AD-D | 3 (17.6%) | 5 (19.2%) | 3 (17.6%) | 3 (11.5%) | 4 (23.5%) | 6 (23.1%) | 6 (35.3%) | 8 (30.8%) | 1 (5.9%) | 4 (15.4%) |
| Sex ($n_{male}$/ $n_{female}$) | CU | 12/9 | 14/15 | NA | NA | 6/4 | 8/6 | 5/2 | 9/2 | 6/17 | 6/29 |
| | MCI | 4/5 | 9/9 | 1/2 | 2/2 | 1/1 | 5/4 | 7/7 | 14/11 | 1/4 | 4/7 |
| | AD-D | 0/3 | 1/4 | 1/2 | 1/2 | 2/2 | 2/4 | 4/2 | 5/3 | 0/1 | 2/2 |
| MMSE | CU | 28.2±1.7 | 28.3±1.5 | NA | NA | 28.6±2.0 | 28.6±1.7 | 27.7±1.6 | 27.8±1.4 | 28.6±1.0 | 28.6±1.1 |
| | MCI | 26.3±1.9 | 27.4±2.0 | 26.7±1.5 | 27.0±1.4 | 25.0±1.4 | 27.3±1.6 | 25.5±2.6 | 26.4±2.3 | 27.6±1.9 | 27.6±1.9 |
| | AD-D* | 18.7±1.5 | 19.0±2.1 | 18.7±3.1 | 18.7±3.1 | 22.8±1.7 | 19.7±6.9 | 23.2±3.0 | 22.4±3.1 | 22.0 | 23.8±3.1 |
| APOE4 carriers ($n_{APOE}$/ $n_{total}$) | CU | 9/21 | 9/29 | NA | NA | 7/10 | 9/14 | 4/7 | 5/11 | 9/23 | 10/35 |
| | MCI | 7/9 | 8/17 | 2/3 | 2/4 | 1/2 | 3/9 | 11/13 | 15/24 | 2/5 | 2/11 |
| | AD-D | 2/2 | 4/4 | 1/3 | 1/3 | 3/4 | 4/6 | 3/5 | 5/7 | 1/1 | 2/4 |
| Characteristic per diagnostic group | | **Validation Cohort: ARWiBo** $n = 17$ (Aβ+); $n = 767$ | | | | | | | | | |
| | | Diffuse cortical atrophy | | Parieto-temporal atrophy | | Frontal atrophy | | Subcortical atrophy | | Outliers | |
| Age | CU# | NA | 43.3±10.9 | NA | 54.5±15.8 | NA | 56.0±12.9 | NA | 49.3±16.2 | NA | 53.5±15.0 |
| | MCI# | 72.0 | 65.8±8.6 | 70.0 | 68.0±4.4 | 69.8±10.9 | 73.5±8.9 | 72.0±1.4 | 73.4±7.0 | 77.0±0.0 | 68.2±8.7 |
| | AD-D# | 61.5±8.3 | 65.9±10.8 | NA | 59.1±7.8 | 62.5±0.7 | 72.9±6.7 | NA | 71.8±8.0 | 67.0 | 72.3±6.9 |
| N (%) | CU | 0 (NA) | 135 (22.8%) | 0 (NA) | 18 (3.0%) | 0 (NA) | 42 (7.1%) | 0 (NA) | 115 (19.4%) | 0 (NA) | 283 (47.7%) |
| | MCI | 1 (10.0%) | 16 (16.2%) | 1 (10.0%) | 3 (3.0%) | 4 (40.0%) | 19 (19.2%) | 2 (20.0%) | 17 (17.2%) | 2 (20.0%) | 44 (44.4%) |
| | AD-D | 4 (57.1%) | 10 (13.3%) | 0 (0.0%) | 7 (9.3%) | 2 (28.6%) | 30 (40.0%) | 0 (0.0%) | 14 (18.7%) | 1 (14.3%) | 14 (18.7%) |
| Sex ($n_{male}$/ $n_{female}$) | CU | NA | 53/82 | NA | 9/9 | NA | 21/21 | NA | 36/79 | NA | 111/172 |
| | MCI | 0/1 | 4/12 | 1/0 | 1/2 | 1/3 | 6/13 | 1/1 | 10/7 | 1/1 | 15/29 |
| | AD-D | 0/4 | 2/8 | NA | 5/2 | 2/0 | 9/21 | NA | 7/7 | 0/1 | 2/12 |
| MMSE | CU | NA | 28.9±1.4 | NA | 28.9±1.2 | NA | 28.8±1.5 | NA | 28.6±1.9 | NA | 28.6±1.7 |
| | MCI | 29.0 | 26.3±2.3 | 26.0 | 26.0±2.0 | 26.0±2.9 | 26.4±3.0 | 23.0±9.9 | 24.1±3.4 | 22.5±9.2 | 25.6±3.9 |
| | AD-D | 23.5±8.0 | 23.8±5.5 | NA | 20.3±4.4 | 26.5±0.7 | 22.7±6.0 | NA | 24.3±4.1 | 27.0 | 23.4±5.1 |
| APOE4 carriers ($n_{APOE}$/ $n_{total}$) | CU | NA | 12/63 | NA | 0/9 | NA | 3/20 | NA | 13/52 | NA | 27/114 |
| | MCI | 1/1 | 6/12 | 1/1 | 1/2 | 0/2 | 2/10 | 0/1 | 4/8 | 0/1 | 9/23 |
| | AD-D# | 2/3 | 4/5 | NA | 0/6 | 1/2 | 8/18 | NA | 1/4 | 1/1 | 4/6 |
| Characteristic per diagnostic group | | **Validation Cohort: EDSD** $n = 28$ (Aβ+); $n = 347$ | | | | | | | | | |
| | | Diffuse cortical atrophy | | Parieto-temporal atrophy | | Frontal atrophy | | Subcortical atrophy | | Outliers | |
| Age | CU | NA | 68.4±5.2 | NA | 67.5±10.6 | NA | 69.3±5.5 | NA | 69.3±6.8 | NA | 67.7±6.1 |
| | MCI | 69.6±7.2 | 68.5±7.3 | 66.0 | 74.3±6.7 | 68.3±2.6 | 71.7±6.7 | 66.7±9.2 | 71.4±7.2 | 77.3±6.0 | 70.9±8.4 |
| | AD-D | NA | 73.1±6.3 | NA | 67.2±9.9 | NA | 73.5±8.8 | 73.0 | 72.4±7.6 | NA | 75.9±8.5 |
| N (%) | CU | 0 (NA) | 39 (28.7%) | 0 (NA) | 2 (1.5%) | 0 (NA) | 24 (17.6%) | 0 (NA) | 27 (19.9%) | 0 (NA) | 44 (32.4%) |
| | MCI | 10 (37.0%) | 25 (26.6%) | 1 (3.7%) | 6 (6.4%) | 6 (22.2%) | 18 (19.1%) | 7 (25.9%) | 22 (23.4%) | 3 (11.1%) | 23 (24.5%) |
| | AD-D | 0 (0.0%) | 21 (17.9%) | 0 (0.0%) | 12 (10.3%) | 0 (0.0%) | 31 (26.5%) | 1 (100.0%) | 41 (35.0%) | 0 (0.0%) | 12 (10.3%) |
| | CU | NA | 15/24 | NA | 1/1 | NA | 12/12 | NA | 18/9 | NA | 19/25 |

| | | | | | | | | | | | |
|---|---|---|---|---|---|---|---|---|---|---|---|
| Sex ($n_{male}$/ $n_{female}$) | MCI | 7/3 | 15/10 | 1/0 | 5/1 | 4/2 | 8/10 | 5/2 | 12/10 | 1/2 | 12/11 |
| | AD-D | NA | 13/8 | NA | 3/9 | NA | 11/20 | 1/0 | 18/23 | NA | 4/8 |
| MMSE | CU | NA | 28.6±1.3 | NA | 29.5±0.7 | NA | 28.8±1.0 | NA | 28.9±1.1 | NA | 28.7±1.1 |
| | MCI | 26.6±2.1 | 27.0±1.6 | 25.0 | 27.2±1.3 | 25.0±2.5 | 26.5±2.1 | 26.3±3.0 | 25.8±2.3 | 25.3±0.6 | 26.1±2.1 |
| | AD-D[#] | NA | 21.8±4.2 | NA | 13.0±6.1 | NA | 19.5±5.0 | 29.0 | 22.6±2.9 | NA | 20.6±5.6 |
| APOE4 carriers ($n_{APOE}$/ $n_{total}$) | CU | 0/0 | 5/16 | 0/0 | 1/1 | 0/0 | 4/10 | 0/0 | 3/11 | 0/0 | 5/18 |
| | MCI | 4/7 | 6/10 | 0/1 | 1/3 | 3/4 | 3/8 | 6/7 | 7/11 | 3/3 | 9/16 |
| | AD-D | 0/0 | 3/7 | 0/0 | 3/3 | 0/0 | 7/12 | 0/0 | 8/13 | 0/0 | 1/5 |
| **Characteristic per diagnostic group** | | **Validation Cohort: I-ADNI $n = 0$ (Aβ+); $n = 169$** | | | | | | | | | |
| | | Diffuse cortical atrophy | | Parieto-temporal atrophy | | Frontal atrophy | | Subcortical atrophy | | Outliers | |
| Age | CU | NA | NA | NA | NA | NA | 59.5±6.4 | NA | 70.7±8.0 | NA | 79.5±9.2 |
| | MCI | NA | 68.0±7.6 | NA | 70.5±6.4 | NA | 75.4±6.2 | NA | 75.0±1.9 | NA | 69.4±4.3 |
| | AD-D[#] | NA | 74.3±7.3 | NA | 66.3±7.9 | NA | 72.5±8.0 | NA | 78.4±5.3 | NA | 71.1±7.5 |
| N (%) | CU | 0 (NA) | 0 (0.0%) | 0 (NA) | 0 (0.0%) | 0 (NA) | 2 (28.6%) | 0 (NA) | 3 (42.9%) | 0 (NA) | 2 (28.6%) |
| | MCI | 0 (NA) | 16 (45.7%) | 0 (NA) | 2 (5.7%) | 0 (NA) | 7 (20.0%) | 0 (NA) | 5 (14.3%) | 0 (NA) | 5 (14.3%) |
| | AD-D | 0 (NA) | 40 (31.5%) | 0 (NA) | 24 (18.9%) | 0 (NA) | 29 (22.8%) | 0 (NA) | 18 (14.2%) | 0 (NA) | 16 (12.6%) |
| Sex ($n_{male}$/ $n_{female}$) | CU | NA | NA | NA | NA | NA | 1/1 | NA | 3/0 | NA | 0/2 |
| | MCI | NA | 10/6 | NA | 1/1 | NA | 5/2 | NA | 1/4 | NA | 1/4 |
| | AD-D | NA | 17/23 | NA | 6/18 | NA | 10/19 | NA | 6/12 | NA | 3/13 |
| MMSE | CU | NA | NA | NA | NA | NA | 28.5±2.1 | NA | 29.3±0.6 | NA | 28.5±2.1 |
| | MCI | NA | 27.1±1.7 | NA | 28.5±2.1 | NA | 26.3±1.9 | NA | 24.0±4.1 | NA | 27.0±2.9 |
| | AD-D | NA | 20.4±3.7 | NA | 18.8±4.9 | NA | 21.0±4.4 | NA | 20.2±4.1 | NA | 20.9±3.8 |
| APOE4 carriers ($n_{APOE}$/ $n_{total}$) | CU | NA | 0/0 | NA | 0/0 | NA | 0/0 | NA | 0/0 | NA | 0/0 |
| | MCI | NA | 0/0 | NA | 0/0 | NA | 0/0 | NA | 0/0 | NA | 0/0 |
| | AD-D | NA | 0/0 | NA | 0/0 | NA | 0/0 | NA | 0/0 | NA | 0/0 |
| **Characteristic per diagnostic group** | | **Validation Cohort: NACC $n = 190$ (Aβ+); $n = 1{,}085$** | | | | | | | | | |
| | | Diffuse cortical atrophy | | Parieto-temporal atrophy | | Frontal atrophy | | Subcortical atrophy | | Outliers | |
| Age | CU | NA | NA | NA | NA | NA | NA | NA | NA | NA | NA |
| | MCI[#] | 76.1±11.9 | 73.4±10.0 | 64.0±15.6 | 67.3±13.5 | 76.4±5.4 | 74.9±9.0 | 76.4±6.0 | 74.2±8.4 | 73.0±14.7 | 71.4±11.1 |
| | AD-D*[#] | 70.2±10.9 | 73.0±9.6 | 62.1±9.0 | 64.5±10.6 | 74.4±11.0 | 73.2±10.3 | 74.8±9.8 | 74.6±9.0 | 69.2±10.3 | 69.2±12.7 |
| N (%) | CU | 0 (NA) | 0 (NA) | 0 (NA) | 0 (NA) | 0 (NA) | 0 (NA) | 0 (NA) | 0 (NA) | 0 (NA) | 0 (NA) |
| | MCI | 25 (39.1%) | 159 (28.1%) | 2 (3.1%) | 20 (3.5%) | 8 (12.5%) | 118 (20.8%) | 18 (28.1%) | 166 (29.3%) | 11 (17.2%) | 103 (18.2%) |
| | AD-D | 25 (19.8%) | 109 (21.0%) | 19 (15.1%) | 58 (11.2%) | 34 (27.0%) | 148 (28.5%) | 31 (24.6%) | 128 (24.7%) | 17 (13.5%) | 76 (14.6%) |
| Sex ($n_{male}$/ $n_{female}$) | CU | NA | NA | NA | NA | NA | NA | NA | NA | NA | NA |
| | MCI | 11/14 | 65/94 | 1/1 | 12/8 | 4/4 | 55/63 | 12/6 | 87/79 | 4/7 | 25/78 |
| | AD-D | 12/13 | 48/61 | 12/7 | 26/32 | 20/14 | 68/80 | 19/12 | 66/62 | 9/8 | 32/44 |
| MMSE | CU | NA | NA | NA | NA | NA | NA | NA | NA | NA | NA |
| | MCI[#] | 25.0±2.0 | 26.0±3.2 | 23.0 | 25.6±3.8 | 25.7±2.3 | 26.1±3.6 | 26.4±2.5 | 27.1±2.6 | 28.3±1.4 | 27.3±2.5 |
| | AD-D*[#] | 23.9±3.9 | 22.2±5.4 | 18.7±7.3 | 18.6±6.5 | 20.1±6.1 | 21.4±5.4 | 22.3±4.4 | 21.2±5.2 | 19.2±7.0 | 20.6±6.4 |
| APOE4 carriers ($n_{APOE}$/ $n_{total}$) | CU | NA | NA | NA | NA | NA | NA | NA | NA | NA | NA |
| | MCI | 13/24 | 61/140 | 2/2 | 6/16 | 5/8 | 43/103 | 14/18 | 69/141 | 6/11 | 32/87 |
| | AD-D | 15/24 | 54/90 | 13/17 | 31/49 | 14/31 | 56/124 | 19/30 | 66/112 | 12/17 | 26/58 |
| | | **Validation Cohort: OASIS $n = 0$ (Aβ+); $n = 302$** | | | | | | | | | |

| Characteristic per diagnostic group | | Diffuse cortical atrophy | | Parieto-temporal atrophy | | Frontal atrophy | | Subcortical atrophy | | Outliers | |
|---|---|---|---|---|---|---|---|---|---|---|---|
| Age | CU | NA | 66.7±13.7 | NA | 63.5±23.3 | NA | 68.3±12.7 | NA | 70.0±12.0 | NA | 70.7±10.5 |
| | MCI | NA | 78.2±8.3 | NA | 68.5±2.1 | NA | 73.4±7.6 | NA | 76.3±5.0 | NA | 75.1±8.3 |
| | AD-D# | NA | 79.7±7.5 | NA | NA | NA | 66.0±5.6 | NA | 75.1±4.5 | NA | 76.2±4.3 |
| N (%) | CU | 0 (NA) | 34 (18.4%) | 0 (NA) | 2 (1.1%) | 0 (NA) | 29 (15.7%) | 0 (NA) | 34 (18.4%) | 0 (NA) | 86 (46.5%) |
| | MCI | 0 (NA) | 22 (24.4%) | 0 (NA) | 2 (2.2%) | 0 (NA) | 14 (15.6%) | 0 (NA) | 29 (32.2%) | 0 (NA) | 23 (25.6%) |
| | AD-D | 0 (NA) | 11 (40.7%) | 0 (NA) | 0 (0.0%) | 0 (NA) | 3 (11.1%) | 0 (NA) | 8 (29.6%) | 0 (NA) | 5 (18.5%) |
| Sex ($n_{male}$/ $n_{female}$) | CU | NA | 13/21 | NA | 0/2 | NA | 10/19 | NA | 17/17 | NA | 13/73 |
| | MCI | NA | 10/12 | NA | 2/0 | NA | 10/4 | NA | 14/15 | NA | 9/14 |
| | AD-D# | NA | 0/11 | NA | NA | NA | 1/2 | NA | 4/4 | NA | 2/3 |
| MMSE | CU | NA | 29.1±1.2 | NA | 29.5±0.7 | NA | 29.6±0.6 | NA | 28.9±1.2 | NA | 29.2±0.9 |
| | MCI | NA | 25.4±3.7 | NA | 25.0±2.8 | NA | 26.1±2.7 | NA | 26.2±2.6 | NA | 27.0±3.2 |
| | AD-D | NA | 22.2±3.8 | NA | NA | NA | 22.3±7.1 | NA | 21.6±4.3 | NA | 22.4±1.9 |
| APOE4 carriers ($n_{APOE}$/ $n_{total}$) | CU | NA | 0/0 | NA | 0/0 | NA | 0/0 | NA | 0/0 | NA | 0/0 |
| | MCI | NA | 0/0 | NA | 0/0 | NA | 0/0 | NA | 0/0 | NA | 0/0 |
| | AD-D | NA | 0/0 | NA | 0/0 | NA | 0/0 | NA | 0/0 | NA | 0/0 |
| **Characteristic per diagnostic group** | | **Validation Cohort: PharmaCog $n = 48$ (Aβ+); $n = 100$** | | | | | | | | | |
| | | Diffuse cortical atrophy | | Parieto-temporal atrophy | | Frontal atrophy | | Subcortical atrophy | | Outliers | |
| Age | CU | NA | NA | NA | NA | NA | NA | NA | NA | NA | NA |
| | MCI | 71.3±7.7 | 70.0±8.1 | 75.0±1.4 | 77.0±3.6 | 70.7±6.6 | 67.9±6.5 | 68.3±6.1 | 67.5±7.5 | 70.2±5.2 | 68.0±7.1 |
| | AD-D | NA | NA | NA | NA | NA | NA | NA | NA | NA | NA |
| N (%) | CU | 0 (NA) | 0 (NA) | 0 (NA) | 0 (NA) | 0 (NA) | 0 (NA) | 0 (NA) | 0 (NA) | 0 (NA) | 0 (NA) |
| | MCI | 11 (22.9%) | 26 (26.0%) | 2 (4.2%) | 3 (3.0%) | 10 (20.8%) | 19 (19.0%) | 17 (35.4%) | 27 (27.0%) | 8 (16.7%) | 25 (25.0%) |
| | AD-D | 0 (NA) | 0 (NA) | 0 (NA) | 0 (NA) | 0 (NA) | 0 (NA) | 0 (NA) | 0 (NA) | 0 (NA) | 0 (NA) |
| Sex ($n_{male}$/ $n_{female}$) | CU | NA | NA | NA | NA | NA | NA | NA | NA | NA | NA |
| | MCI | 5/6 | 11/15 | 1/1 | 2/1 | 4/6 | 8/11 | 8/9 | 12/15 | 3/5 | 7/18 |
| | AD-D | NA | NA | NA | NA | NA | NA | NA | NA | NA | NA |
| MMSE | CU | NA | NA | NA | NA | NA | NA | NA | NA | NA | NA |
| | MCI | 26.4±1.9 | 27.2±1.7 | 24.5±2.1 | 26.3±3.5 | 25.6±1.8 | 25.9±1.5 | 26.1±1.4 | 26.2±1.5 | 26.9±2.0 | 27.1±2.1 |
| | AD-D | NA | NA | NA | NA | NA | NA | NA | NA | NA | NA |
| APOE4 carriers ($n_{APOE}$/ $n_{total}$) | CU | NA | NA | NA | NA | NA | NA | NA | NA | NA | NA |
| | MCI*# | 5/11 | 6/26 | 1/2 | 1/3 | 9/10 | 10/19 | 14/16 | 17/26 | 5/7 | 7/24 |
| | AD-D | NA | NA | NA | NA | NA | NA | NA | NA | NA | NA |
| **Characteristic per diagnostic group** | | **Validation Cohort: GMC (external)** | | | | | | | | | |
| | | Diffuse cortical atrophy | | Parieto-temporal atrophy | | Frontal atrophy | | Subcortical atrophy | | Outliers | |
| Age | CU | 73.5±3.5 | 67.8±11.1 | NA | 66.4±3.7 | 67.5±6.8 | 64.8±9.3 | 71.3±1.7 | 66.3±11.8 | 74.0±2.0 | 64.3±10.3 |
| | MCI | 72.6±7.1 | 73.8±9.9 | 69.5±8.1 | 73.4±9.1 | 73.7±2.8 | 70.0±9.5 | 73.9±5.1 | 72.8±9.9 | 73.2±8.1 | 71.0±11.4 |
| | AD-D | 69.9±7.5 | 76.5±8.6 | 71.2±9.2 | 72.3±9.2 | 66.8±6.5 | 75.4±6.9 | 68.4±14.4 | 77.5±10.0 | 77.0±2.0 | 80.1± 4.1 |
| N (%) | CU | 2 (12.5%) | 46 (27.5%) | NA | 5 (3.0%) | 4 (25.0%) | 28 (16.8%) | 8 (50.0%) | 38 (22.8%) | 2 (12.5%) | 50 (29.9%) |
| | MCI | 26 (24.1%) | 112 (26.7%) | 4 (3.7%) | 15 (3.6%) | 9 (8.3%) | 69 (16.5%) | 51 (47.2%) | 134 (32.0%) | 18 (16.7%) | 89 (21.2%) |
| | AD-D | 13 (37.1%) | 40 (29.2%) | 5 (14.3%) | 8 (5.8%) | 6 (17.1%) | 35 (25.6%) | 9 (25.7%) | 40 (29.2%) | 2 (5.7%) | 14 (10.2%) |
| | CU | 1/1 | 21/25 | NA | 3/2 | 2/2 | 11/17 | 3/5 | 14/24 | 0/2 | 8/42 |

| | | | | | | | | | | | |
|---|---|---|---|---|---|---|---|---|---|---|---|
| Sex ($n_{male}$/ $n_{female}$) | MCI | 11/15 | 53/59 | 3/1 | 8/7 | 7/2 | 36/33 | 19/32 | 61/73 | 6/12 | 25/64 |
| | AD-D | 10/3 | 21/19 | 2/3 | 2/6 | 4/2 | 12/23 | 3/6 | 16/24 | 2/0 | 6/8 |
| MMSE | CU | 28.5±0.5 | 28.4±1.5 | NA | 28.2±1.5 | 27.8±1.1 | 28.5±1.1 | 28.5±0.9 | 28.6±1.4 | 29.0±0.0 | 28.3±1.6 |
| | MCI | 25.3±2.9 | 25.4±3.3 | 20.3±9.0 | 24.6±5.9 | 26.3± 2.6 | 26.1±2.6 | 25.1±2.9 | 25.5±3.3 | 26.1±2.6 | 25.6±3.5 |
| | AD-D | 19.3±5.0 | 19.4±4.6 | 20.2±4.2 | 18.3±5.3 | 20.7±4.1 | 19.5±5.3 | 19.1±4.6 | 20.8±4.6 | 20.5±4.5 | 21.8± 3.6 |
| APOE4 carriers ($n_{APOE}$/ $n_{total}$) | CU | 1/2 | 5/6 | NA | 0/2 | 1/2 | 3/9 | 4/7 | 5/14 | 0/1 | 4/7 |
| | MCI | 9/13 | 13/25 | 1/1 | 2/2 | 4/7 | 8/15 | 9/15 | 13/25 | 2/5 | 4/12 |
| | AD-D | 0/1 | 2/3 | 1/1 | 1/1 | 0/1 | 1/2 | 0/0 | 0/1 | 0/0 | 1/1 |

**Supplementary Table 5**: Characteristics of atrophy-based subtypes assigned by the trained SuStaIn model in the internal and external validation cohorts. * indicates the corresponding measure is significantly different ($p < 0.05$) between the different subtypes (excluding the outliers group) in Aβ+ validation cohort, using ANOVA test for Age and MMSE characteristics, and $\chi^2$ contingency test for *APOE4* and Sex characteristics. $^{\#}$ indicates the significant difference ($p < 0.05$) using similar tests in the clinical validation cohort. Abbreviations: CU: Cognitively unimpaired (Cognitively normal or subjective cognitive decline); MCI: Mild cognitive impairment; AD-D: Alzheimer's disease dementia; NA: Not applicable.

| | | Typical | | Limbic-predominant | | Hippocampal-sparing | | Subcortical atrophy | | Outliers | |
|---|---|---|---|---|---|---|---|---|---|---|---|
| | | Aβ+ | All | Aβ+ | All | Aβ+ | All | Aβ+ | All | Aβ+ | All |
| **Characteristic per diagnostic group** | | **Validation Cohort: ADC** $n = 937$ (Aβ+); $n = 2{,}312$ | | | | | | | | | |
| Age | CU$^{\#}$ | 66.9±8.6 | 64.8±9.7 | 63.8±10.6 | 60.1±13.6 | 61.0±15.6 | 59.0±12.4 | NA | 68.5±2.1 | 63.0±9.3 | 59.5±9.9 |
| | MCI$^{\#}$ | 67.9±7.3 | 67.9±7.7 | 65.7±8.5 | 65.6±9.4 | 64.5±8.5 | 65.1±8.0 | 72.5±6.4 | 71.1±5.6 | 66.3±7.7 | 65.1±7.6 |
| | AD-D*$^{\#}$ | 63.8±7.8 | 65.9±8.5 | 67.0±7.1 | 69.7±7.8 | 60.0±6.7 | 60.9±7.0 | 70.3±4.0 | 71.1±4.7 | 64.7±9.0 | 66.8±8.8 |
| N (%) | CU | 38 (38.0%) | 186 (33.5%) | 9 (9.0%) | 62 (11.2%) | 2 (2.0%) | 14 (2.5%) | 0 (0.0%) | 2 (0.4%) | 51 (51.0%) | 292 (52.5%) |
| | MCI | 106 (52.0%) | 314 (47.6%) | 37 (18.1%) | 126 (19.1%) | 12 (5.9%) | 25 (3.8%) | 2 (1.0%) | 12 (1.8%) | 47 (23.0%) | 182 (27.6%) |
| | AD-D | 368 (58.1%) | 625 (57.0%) | 139 (22.0%) | 264 (24.1%) | 85 (13.4%) | 125 (11.4%) | 3 (0.5%) | 8 (0.7%) | 38 (6.0%) | 75 (6.8%) |
| Sex ($n_{male}$/ $n_{female}$) | CU | 19/19 | 112/74 | 7/2 | 46/16 | 0/2 | 7/7 | 0/0 | 2/0 | 22/29 | 123/169 |
| | MCI | 58/48 | 215/99 | 23/14 | 82/44 | 8/4 | 16/9 | 1/1 | 8/4 | 17/30 | 94/88 |
| | AD-D | 162/206 | 286/339 | 70/69 | 143/121 | 32/53 | 53/72 | 1/2 | 4/4 | 5/33 | 16/59 |
| MMSE | CU | 27.7±1.6 | 28.1±1.6 | 29.0±1.3 | 28.4±1.6 | 27.5±0.7 | 28.1±1.3 | NA | 29.0±0.0 | 28.2±1.2 | 28.3±1.8 |
| | MCI* | 26.7±2.1 | 26.6±2.4 | 25.7±2.2 | 26.0±2.3 | 24.9±3.4 | 26.0±2.9 | 24.5±0.7 | 27.3±2.2 | 26.8±2.2 | 26.6±2.3 |
| | AD-D*$^{\#}$ | 20.6±5.2 | 20.3±5.3 | 21.0±4.3 | 20.8±5.0 | 18.7±5.7 | 18.3±5.5 | 20.0±4.6 | 22.9±3.8 | 23.4±3.2 | 23.5±3.3 |
| APOE4 carriers ($n_{APOE}$/ $n_{total}$) | CU | 23/38 | 59/186 | 7/9 | 13/62 | 1/2 | 3/14 | 0/0 | 0/2 | 30/51 | 72/292 |
| | MCI | 72/106 | 132/314 | 27/37 | 54/126 | 9/12 | 14/25 | 1/2 | 3/12 | 31/47 | 65/182 |
| | AD-D*$^{\#}$ | 225/368 | 298/625 | 107/139 | 156/264 | 57/85 | 75/125 | 3/3 | 6/8 | 27/38 | 42/75 |
| **Characteristic per diagnostic group** | | **Validation Cohort: ADNI** $n = 465$ (Aβ+); $n = 1{,}098$ | | | | | | | | | |
| | | Typical | | Limbic-predominant | | Hippocampal-sparing | | Subcortical atrophy | | Outliers | |
| Age | CU | 75.9±6.4 | 76.0±5.4 | 70.2±4.0 | 71.9±4.3 | 85.0 | 78.3±4.5 | 76.0 | 77.0±1.4 | 72.5±5.4 | 73.1±5.5 |
| | MCI | 72.6±6.0 | 73.1±7.1 | 72.8±4.8 | 73.0±5.6 | 69.3±12.5 | 68.7±9.7 | 77.0±2.6 | 73.3±4.9 | 71.1±7.2 | 70.3±7.7 |
| | AD-D*$^{\#}$ | 73.2±9.2 | 74.3±8.1 | 74.2±6.7 | 74.9±7.1 | 66.5±7.1 | 68.0±6.8 | NA | NA | 76.9±8.2 | 75.6±7.8 |
| N (%) | CU | 25 (21.7%) | 62 (27.2%) | 6 (5.2%) | 10 (4.4%) | 1 (0.9%) | 6 (2.6%) | 1 (0.9%) | 2 (0.9%) | 82 (71.3%) | 148 (64.9%) |
| | MCI | 114 (47.5%) | 274 (40.8%) | 42 (17.5%) | 106 (15.8%) | 6 (2.5%) | 18 (2.7%) | 3 (1.2%) | 6 (0.9%) | 75 (31.2%) | 267 (39.8%) |
| | AD-D | 52 (47.3%) | 100 (50.3%) | 31 (28.2%) | 55 (27.6%) | 13 (11.8%) | 21 (10.6%) | 0 (0.0%) | 0 (0.0%) | 14 (12.7%) | 23 (11.6%) |
| Sex ($n_{male}$/ $n_{female}$) | CU | 19/6 | 40/22 | 5/1 | 8/2 | 1/0 | 4/2 | 1/0 | 2/0 | 26/56 | 58/90 |
| | MCI | 70/44 | 175/99 | 26/16 | 68/38 | 5/1 | 12/6 | 3/0 | 5/1 | 40/35 | 130/137 |
| | AD-D | 27/25 | 55/45 | 15/16 | 27/28 | 6/7 | 10/11 | 0/0 | 0/0 | 5/9 | 7/16 |
| MMSE | CU | 28.8±1.4 | 29.1±1.1 | 28.7±1.0 | 28.8±1.0 | 28.0 | 28.8±0.4 | 29.0 | 29.5±0.7 | 29.3±0.9 | 29.2±1.0 |
| | MCI | 27.3±2.1 | 27.3±1.9 | 27.4±1.7 | 27.6±1.8 | 26.2±1.5 | 27.6±1.5 | 27.7±0.6 | 28.7±1.2 | 27.9±2.0 | 28.1±1.7 |
| | AD-D | 23.4±2.1 | 23.3±2.1 | 23.3±2.4 | 23.2±2.2 | 22.4±2.2 | 22.4±2.2 | NA | NA | 23.2±3.0 | 23.2±2.8 |
| APOE4 carriers ($n_{APOE}$/ | CU | 10/25 | 19/62 | 3/6 | 3/10 | 0/1 | 3/6 | 0/1 | 0/2 | 45/82 | 61/143 |
| | MCI | 79/110 | 146/266 | 24/40 | 50/103 | 2/6 | 6/18 | 2/3 | 3/5 | 45/72 | 100/255 |
| | AD-D | 40/52 | 69/99 | 27/31 | 38/54 | 13/13 | 19/21 | 0/0 | 0/0 | 11/14 | 16/23 |

| | | | | | | | | | | | |
|---|---|---|---|---|---|---|---|---|---|---|---|
| $n_{total}$) | | | | | | | | | | | |
| **Characteristic per diagnostic group** | | **Validation Cohort: AIBL** $n = 111$ (Aβ+); $n = 182$ | | | | | | | | | |
| | | Typical | | Limbic-predominant | | Hippocampal-sparing | | Subcortical atrophy | | Outliers | |
| Age | CU | 78.0±6.3 | 77.1±5.6 | 75.5±6.4 | 72.7±6.7 | 82.7±3.5 | 82.7±3.5 | NA | NA | 74.3±6.8 | 73.7±6.7 |
| | MCI# | 74.5±6.2 | 73.9±6.4 | 79.7±7.2 | 78.9±5.4 | NA | NA | NA | NA | 74.8±5.4 | 73.7±7.8 |
| | AD-D*# | 67.4±11.2 | 71.5±10.2 | 75.5±6.2 | 76.6±6.1 | 61.0±4.2 | 61.0±4.2 | NA | NA | NA | 73.0±9.0 |
| N (%) | CU | 10 (16.4%) | 16 (18.0%) | 2 (3.3%) | 3 (3.4%) | 3 (4.9%) | 3 (3.4%) | 0 (0.0%) | 0 (0.0%) | 46 (75.4%) | 67 (75.3%) |
| | MCI | 15 (45.5%) | 22 (32.8%) | 6 (18.2%) | 11 (16.4%) | 0 (0.0%) | 0 (0.0%) | 0 (0.0%) | 0 (0.0%) | 12 (36.4%) | 34 (50.7%) |
| | AD-D | 5 (29.4%) | 8 (30.8%) | 10 (58.8%) | 13 (50.0%) | 2 (11.8%) | 2 (7.7%) | 0 (0.0%) | 0 (0.0%) | 0 (0.0%) | 3 (11.5%) |
| Sex ($n_{male}$/ $n_{female}$) | CU | 7/3 | 9/7 | 1/1 | 2/1 | 3/0 | 3/0 | NA | NA | 18/28 | 23/44 |
| | MCI | 9/6 | 13/9 | 3/3 | 6/5 | NA | NA | NA | NA | 2/10 | 15/19 |
| | AD-D | 1/4 | 2/6 | 5/5 | 6/7 | 1/1 | 1/1 | NA | NA | 0/0 | 2/1 |
| MMSE | CU | 28.3±1.7 | 28.4±1.5 | 26.0±0.0 | 26.3±0.6 | 27.3±2.1 | 27.3±2.1 | NA | NA | 28.5±1.4 | 28.6±1.3 |
| | MCI | 25.0±2.0 | 25.9±2.2 | 26.0±2.3 | 26.8±2.0 | NA | NA | NA | NA | 27.6±1.7 | 27.9±1.5 |
| | AD-D | 20.4±2.1 | 18.4±5.4 | 22.4±3.1 | 21.9±3.0 | 19.0±4.2 | 19.0±4.2 | NA | NA | NA | 24.3±3.5 |
| APOE4 carriers ($n_{APOE}$/ $n_{total}$) | CU | 5/10 | 5/16 | 1/2 | 2/3 | 2/3 | 2/4 | NA | NA | 21/46 | 24/67 |
| | MCI | 10/14 | 13/21 | 6/6 | 7/11 | NA | NA | NA | NA | 7/12 | 10/33 |
| | AD-D# | 4/5 | 7/8 | 6/8 | 8/11 | 0/2 | 0/2 | NA | NA | NA | 1/3 |
| **Characteristic per diagnostic group** | | **Validation Cohort: ARWiBo** $n = 17$ (Aβ+); $n = 767$ | | | | | | | | | |
| | | Typical | | Limbic-predominant | | Hippocampal-sparing | | Subcortical atrophy | | Outliers | |
| Age | CU | NA | 49.7±15.6 | NA | 43.6±18.8 | NA | 48.8±14.4 | NA | 50.0 | NA | 51.1±14.6 |
| | MCI# | 68.8±8.2 | 71.7±9.5 | NA | 76.1±7.2 | NA | 68.0±2.6 | NA | NA | 75.0±4.2 | 68.2±8.4 |
| | AD-D# | 64.5±3.5 | 71.3±9.4 | 63.0 | 70.8±8.0 | 50.0 | 59.6±7.5 | NA | NA | 67.0 | 71.2±5.4 |
| N (%) | CU | NA | 71 (12.0%) | NA | 25 (4.2%) | NA | 13 (2.2%) | NA | 1 (0.2%) | NA | 483 (81.5%) |
| | MCI | 5 (50.0%) | 23 (23.2%) | 0 (0.0%) | 9 (9.1%) | 0 (0.0%) | 3 (3.0%) | 0 (0.0%) | 0 (0.0%) | 5 (50.0%) | 64 (64.6%) |
| | AD-D | 4 (57.1%) | 39 (52.0%) | 1 (14.3%) | 14 (18.7%) | 1 (14.3%) | 5 (6.7%) | 0 (0.0%) | 0 (0.0%) | 1 (14.3%) | 17 (22.7%) |
| Sex ($n_{male}$/ $n_{female}$) | CU | NA | 22/49 | NA | 13/12 | NA | 7/6 | NA | 0/1 | NA | 188/295 |
| | MCI | 3/2 | 9/14 | NA | 4/5 | NA | 1/2 | NA | NA | 1/4 | 22/42 |
| | AD-D | 1/3 | 10/29 | 1/0 | 7/7 | 0/1 | 2/3 | NA | NA | 0/1 | 6/11 |
| MMSE | CU | NA | 28.6±2.2 | NA | 28.6±1.4 | NA | 28.9±1.0 | NA | 28.0 | NA | 28.7±1.6 |
| | MCI | 27.4±2.9 | 25.8±3.0 | NA | 25.2±2.7 | NA | 27.7±3.2 | NA | NA | 22.6±6.3 | 25.5±3.7 |
| | AD-D | 22.8±7.5 | 22.7±5.0 | 27.0 | 25.0±4.4 | 29.0 | 20.0±5.9 | NA | NA | 27.0 | 23.1±6.4 |
| APOE4 carriers ($n_{APOE}$/ $n_{total}$) | CU | NA | 9/33 | NA | 3/11 | NA | 1/7 | NA | 0/1 | NA | 42/206 |
| | MCI | 1/3 | 5/14 | NA | 2/4 | NA | 0/2 | NA | NA | 1/3 | 15/35 |
| | AD-D | 2/4 | 10/24 | 1/1 | 4/8 | 0/0 | 1/3 | NA | NA | 1/1 | 2/4 |
| **Characteristic per diagnostic group** | | **Validation Cohort: EDSD** $n = 28$ (Aβ+); $n = 347$ | | | | | | | | | |
| | | Typical | | Limbic-predominant | | Hippocampal-sparing | | Subcortical atrophy | | Outliers | |
| Age | CU | NA | 70.3±6.9 | NA | 68.7±6.5 | NA | 66.0 | NA | NA | NA | 68.1±5.6 |
| | MCI | 67.2±7.2 | 70.0±7.9 | 75.2±3.6 | 74.6±2.6 | NA | 59.0 | NA | 73.0 | 72.0±6.5 | 71.0±7.5 |
| | AD-D | NA | 72.0±8.4 | 73.0 | 71.4±8.1 | NA | 71.2±7.5 | NA | 70.0 | NA | 78.2±6.0 |
| N (%) | CU | NA | 25 (18.4%) | NA | 9 (6.6%) | NA | 1 (0.7%) | NA | 0 (0.0%) | NA | 101 (74.3%) |
| | MCI | 18 (66.7%) | 43 (45.7%) | 4 (14.8%) | 9 (9.6%) | 0 (0.0%) | 1 (1.1%) | 0 (0.0%) | 1 (1.1%) | 5 (18.5%) | 40 (42.6%) |
| | AD-D | 0 (0.0%) | 66 (56.4%) | 1 (100.0%) | 29 (24.8%) | 0 (0.0%) | 5 (4.3%) | 0 (0.0%) | 1 (0.9%) | 0 (0.0%) | 16 (13.7%) |

| | | | | | | | | | | | |
|---|---|---|---|---|---|---|---|---|---|---|---|
| Sex ($n_{male}$/ $n_{female}$) | CU | NA | 17/8 | NA | 7/2 | NA | 1/0 | NA | NA | NA | 40/61 |
| | MCI | 12/6 | 23/20 | 3/1 | 7/2 | NA | 0/1 | NA | 0/1 | 3/2 | 22/18 |
| | AD-D | NA | 27/39 | 1/0 | 12/17 | NA | 4/1 | NA | 1/0 | NA | 5/11 |
| MMSE | CU | NA | 28.6±1.3 | NA | 28.2±0.8 | NA | 30.0 | NA | NA | NA | 28.8±1.1 |
| | MCI* | 25.7±2.5 | 26.3±2.2 | 27.5±1.7 | 26.2±2.1 | NA | 27.0 | NA | 28.0 | 25.8±1.9 | 26.5±1.9 |
| | AD-D | NA | 19.5±5.5 | 29.0 | 21.5±4.2 | NA | 18.4±9.0 | NA | 20.0 | NA | 23.1±2.8 |
| APOE4 carriers ($n_{APOE}$ /$n_{total}$) | CU | NA | 2/6 | NA | 2/7 | NA | 0/0 | NA | 0/0 | 0/0 | 14/43 |
| | MCI | 8/14 | 10/22 | 4/4 | 4/6 | NA | 0/0 | NA | 0/0 | 4/4 | 12/20 |
| | AD-D | NA | 11/16 | 0/0 | 6/12 | NA | 3/4 | NA | 0/1 | 0/0 | 2/7 |
| **Characteristic per diagnostic group** | | **Validation Cohort: I-ADNI** $n = 0$ (Aβ+); $n = 169$ | | | | | | | | | |
| | | Typical | | Limbic-predominant | | Hippocampal-sparing | | Subcortical atrophy | | Outliers | |
| Age | CU | NA | 65.7±12.2 | NA | 70.0 | NA | NA | NA | NA | NA | 74.3±11.1 |
| | MCI | NA | 70.9±7.6 | NA | 74.0±2.9 | NA | 67.0±8.7 | NA | NA | NA | 70.4±6.7 |
| | AD-D[#] | NA | 73.1±8.0 | NA | 75.7±7.9 | NA | 62.2±7.1 | NA | 75.0 | NA | 73.7±4.2 |
| N (%) | CU | NA | 3 (42.9%) | NA | 1 (14.3%) | NA | 0 (0.0%) | NA | 0 (0.0%) | NA | 3 (42.9%) |
| | MCI | NA | 16 (45.7%) | NA | 5 (14.3%) | NA | 3 (8.6%) | NA | 0 (0.0%) | NA | 11 (31.4%) |
| | AD-D | NA | 77 (60.6%) | NA | 18 (14.2%) | NA | 12 (9.4%) | NA | 1 (0.8%) | NA | 19 (15.0%) |
| Sex ($n_{male}$/ $n_{female}$) | CU | NA | 3/0 | NA | 1/0 | NA | NA | NA | NA | NA | 0/3 |
| | MCI | NA | 12/4 | NA | 2/3 | NA | 3/0 | NA | NA | NA | 1/10 |
| | AD-D | NA | 25/52 | NA | 9/9 | NA | 4/8 | NA | 0/1 | NA | 4/15 |
| MMSE | CU | NA | 29.7±0.6 | NA | 29.0 | NA | NA | NA | NA | NA | 28.0±1.7 |
| | MCI | NA | 26.5±2.4 | NA | 25.0±3.4 | NA | 28.0±0.0 | NA | NA | NA | 27.0±2.5 |
| | AD-D | NA | 19.6±4.3 | NA | 20.9±4.0 | NA | 19.4±3.8 | NA | 15.0 | NA | 23.0±3.1 |
| APOE4 carriers ($n_{APOE}$/ $n_{total}$) | CU | NA | NA | NA | NA | NA | NA | NA | NA | NA | NA |
| | MCI | NA | NA | NA | NA | NA | NA | NA | NA | NA | NA |
| | AD-D | NA | NA | NA | NA | NA | NA | NA | NA | NA | NA |
| **Characteristic per diagnostic group** | | **Validation Cohort: NACC** $n = 190$ (Aβ+); $n = 1,085$ | | | | | | | | | |
| | | Typical | | Limbic-predominant | | Hippocampal-sparing | | Subcortical atrophy | | Outliers | |
| Age | CU | NA | NA | NA | NA | NA | NA | NA | NA | NA | NA |
| | MCI[#] | 77.7±9.3 | 75.3±8.8 | 73.3±12.9 | 72.6±8.9 | 73.8±10.4 | 68.5±13.0 | NA | 82.0 | 70.8±9.4 | 71.8±10.7 |
| | AD-D*[#] | 71.1±11.6 | 72.3±10.8 | 75.8±9.2 | 73.7±8.8 | 61.9±6.3 | 63.8±9.2 | 70.5±17.7 | 72.2±14.8 | 66.2±5.8 | 72.9±11.3 |
| N (%) | CU | NA | NA | NA | NA | NA | NA | NA | NA | NA | NA |
| | MCI | 34 (53.1%) | 248 (43.8%) | 17 (26.6%) | 101 (17.8%) | 4 (6.2%) | 19 (3.4%) | 0 (0.0%) | 1 (0.2%) | 9 (14.1%) | 197 (34.8%) |
| | AD-D | 72 (57.1%) | 273 (52.6%) | 33 (26.2%) | 111 (21.4%) | 14 (11.1%) | 46 (8.9%) | 2 (1.6%) | 8 (1.5%) | 5 (4.0%) | 81 (15.6%) |
| Sex ($n_{male}$/ $n_{female}$) | CU | NA | NA | NA | NA | NA | NA | NA | NA | NA | NA |
| | MCI | 17/17 | 123/125 | 10/7 | 53/48 | 1/3 | 9/10 | 0/0 | 0/1 | 4/5 | 59/138 |
| | AD-D | 39/33 | 128/145 | 21/12 | 58/53 | 8/6 | 20/26 | 2/0 | 6/2 | 2/3 | 28/53 |
| MMSE | CU | NA | NA | NA | NA | NA | NA | NA | NA | NA | NA |
| | MCI | 25.7±2.6 | 26.3±3.2 | 25.7±1.6 | 25.9±3.3 | 24.0 | 26.2±3.1 | NA | 27.0 | 28.8±1.5 | 27.2±2.6 |
| | AD-D | 20.6±6.5 | 20.8±5.8 | 20.9±4.5 | 21.4±5.3 | 20.9±6.1 | 20.2±6.0 | 28.0 | 21.4±4.9 | 25.2±2.5 | 22.0±5.6 |
| APOE4 carriers ($n_{APOE}$/ $n_{total}$) | CU | NA | NA | NA | NA | NA | NA | NA | NA | NA | NA |
| | MCI | 22/33 | 97/223 | 11/17 | 47/91 | 2/4 | 7/18 | 0/0 | 1/1 | 5/9 | 59/154 |
| | AD-D | 41/68 | 123/232 | 18/31 | 52/96 | 10/13 | 27/40 | 0/2 | 2/5 | 4/5 | 29/60 |
| | | **Validation Cohort: OASIS** $n = 0$ (Aβ+); $n = 302$ | | | | | | | | | |

| Characteristic per diagnostic group | | Typical | | Limbic-predominant | | Hippocampal-sparing | | Subcortical atrophy | | Outliers | |
|---|---|---|---|---|---|---|---|---|---|---|---|
| Age | CU | NA | 73.7±12.5 | NA | 67.9±20.2 | NA | 70.2±14.4 | NA | NA | NA | 68.8±11.2 |
| | MCI | NA | 76.0±7.9 | NA | 76.5±6.8 | NA | 76.0±7.1 | NA | NA | NA | 75.5±7.3 |
| | AD-D | NA | 74.5±7.6 | NA | 74.2±2.8 | NA | 79.0±9.9 | NA | NA | NA | 80.0±7.1 |
| N (%) | CU | NA | 20 (10.8%) | NA | 7 (3.8%) | NA | 5 (2.7%) | NA | 0 (0.0%) | NA | 153 (82.7%) |
| | MCI | NA | 30 (33.3%) | NA | 14 (15.6%) | NA | 2 (2.2%) | NA | 0 (0.0%) | NA | 44 (48.9%) |
| | AD-D | NA | 13 (48.1%) | NA | 5 (18.5%) | NA | 2 (7.4%) | NA | 0 (0.0%) | NA | 7 (25.9%) |
| Sex ($n_{male}$/ $n_{female}$) | CU | NA | 9/11 | NA | 5/2 | NA | 2/3 | NA | NA | NA | 37/116 |
| | MCI | NA | 12/18 | NA | 10/4 | NA | 2/0 | NA | NA | NA | 21/23 |
| | AD-D | NA | 3/10 | NA | 1/4 | NA | 0/2 | NA | NA | NA | 3/4 |
| MMSE | CU | NA | 29.4±0.8 | NA | 29.0±1.5 | NA | 28.8±1.8 | NA | NA | NA | 29.2±1.0 |
| | MCI | NA | 25.9±3.0 | NA | 25.1±3.7 | NA | 24.5±6.4 | NA | NA | NA | 26.8±2.7 |
| | AD-D | NA | 22.8±4.0 | NA | 19.0±3.7 | NA | 21.0±8.5 | NA | NA | NA | 23.1±1.3 |
| APOE4 carriers ($n_{APOE}$/ $n_{total}$) | CU | NA | NA | NA | NA | NA | NA | NA | NA | NA | NA |
| | MCI | NA | NA | NA | NA | NA | NA | NA | NA | NA | NA |
| | AD-D | NA | NA | NA | NA | NA | NA | NA | NA | NA | NA |
| **Characteristic per diagnostic group** | | **Validation Cohort: PharmaCog $n = 48$ (Aβ+); $n = 100$** | | | | | | | | | |
| | | Typical | | Limbic-predominant | | Hippocampal-sparing | | Subcortical atrophy | | Outliers | |
| Age | CU | NA | NA | NA | NA | NA | NA | NA | NA | NA | NA |
| | MCI | 71.3±7.1 | 71.3±7.7 | 66.3±7.2 | 67.1±7.8 | 67.0 | 72.0±4.4 | NA | NA | 70.1±5.2 | 67.5±7.0 |
| | AD-D | NA | NA | NA | NA | NA | NA | NA | NA | NA | NA |
| N (%) | CU | NA | NA | NA | NA | NA | NA | NA | NA | NA | NA |
| | MCI | 20 (41.7%) | 28 (28.0%) | 6 (12.5%) | 14 (14.0%) | 1 (2.1%) | 3 (3.0%) | 0 (0.0%) | 0 (0.0%) | 21 (43.8%) | 55 (55.0%) |
| | AD-D | NA | NA | NA | NA | NA | NA | NA | NA | NA | NA |
| Sex ($n_{male}$/ $n_{female}$) | CU | NA | NA | NA | NA | NA | NA | NA | NA | NA | NA |
| | MCI | 8/12 | 14/14 | 4/2 | 8/6 | 1/0 | 1/2 | NA | NA | 8/13 | 17/38 |
| | AD-D | NA | NA | NA | NA | NA | NA | NA | NA | NA | NA |
| MMSE | CU | NA | NA | NA | NA | NA | NA | NA | NA | NA | NA |
| | MCI | 25.9±1.7 | 26.5±1.8 | 26.3±1.8 | 26.1±1.7 | 26.0 | 27.7±2.1 | NA | NA | 26.2±1.9 | 26.8±1.9 |
| | AD-D | NA | NA | NA | NA | NA | NA | NA | NA | NA | NA |
| APOE4 carriers ($n_{APOE}$/ $n_{total}$) | CU | NA | NA | NA | NA | NA | NA | NA | NA | NA | NA |
| | MCI | 15/20 | 15/28 | 5/6 | 7/14 | 0/1 | 0/3 | NA | NA | 14/19 | 19/53 |
| | AD-D | NA | NA | NA | NA | NA | NA | NA | NA | NA | NA |
| **Characteristic per diagnostic group** | | **Validation Cohort: GMC (external)** | | | | | | | | | |
| | | Typical | | Limbic-predominant | | Hippocampal-sparing | | Subcortical atrophy | | Outliers | |
| Age | CU | 71.1± 4.8 | 65.9±10.6 | 70.0 | 57.9±10.7 | NA | 65.5±7.5 | NA | NA | 70.0 | 67.8±11.1 |
| | MCI | 73.9±5.8 | 71.8±10.3 | 74.0±5.9 | 74.4±9.3 | 68.8± 7.1 | 70.4±9.5 | NA | 76.5±14.4 | 68.9±6.6 | 70.5±10.4 |
| | AD-D[#] | 68.4±10.9 | 75.9±9.0 | 74.7±2.3 | 79.9±5.3 | 67.0±9.9 | 75.2±11.5 | NA | NA | 72.0±5.0 | 80.0±6.4 |
| N (%) | CU | 14 (87.5%) | 121 (72.5%) | 1 (6.3%) | 7 (4.2%) | NA | 10 (6.0%) | NA | NA | 1 (6.3%) | 29 (17.4%) |
| | MCI | 56 (51.9%) | 257 (61.3%) | 38 (35.2%) | 89 (21.2%) | 4 (3.7%) | 17 (4.1%) | NA | 6 (1.4%) | 10 (9.3%) | 50 (11.9%) |
| | AD-D | 24 (68.5%) | 91 (66.4%) | 6 (17.1%) | 33 (24.1%) | 3 (8.6%) | 6 (4.4%) | NA | NA | 2 (5.7%) | 7 (5.1%) |
| Sex ($n_{male}$/ | CU | 5/9 | 40/81 | 0/1 | 2/5 | NA | 3/7 | NA | NA | 1/0 | 12/27 |
| | MCI[#] | 30/26 | 110/147 | 11/27 | 34/55 | 2/2 | 11/6 | NA | 5/1 | 3/7 | 23/27 |

| | | | | | | | | | | | |
|---|---|---|---|---|---|---|---|---|---|---|---|
| $n_{female}$) | AD-D | 12/12 | 31/60 | 5/1 | 18/15 | 2/1 | 4/2 | NA | NA | 2/0 | 4/3 |
| MMSE | CU | 28.4±1.0 | 28.4±1.4 | 28 | 28.9±1.0 | NA | 28.0±1.8 | NA | NA | 28 | 28.3±1.8 |
| | MCI*# | 25.9± 2.6 | 25.8±3.1 | 24.9±3.0 | 25.1±3.3 | 19.5±8.4 | 23.1±5.2 | NA | 24.3±3.9 | 25.5±3.4 | 26.2±3.1 |
| | AD-D | 19.5±4.6 | 19.5±4.6 | 21.8±2.9 | 21.0±5.5 | 14.7±4.2 | 18.2±5.4 | NA | NA | 23.5±2.5 | 22.7±3.4 |
| APOE4 carriers ($n_{APOE}$/ $n_{total}$) | CU | 4/10 | 12/28 | 1/1 | 1/2 | NA | 0/4 | NA | NA | 1/1 | 4/4 |
| | MCI | 15/24 | 26/49 | 7/13 | 8/16 | 1/2 | 1/2 | NA | NA | 2/4 | 5/12 |
| | AD-D | 1/3 | 5/7 | NA | 0/1 | NA | NA | NA | NA | NA | NA |

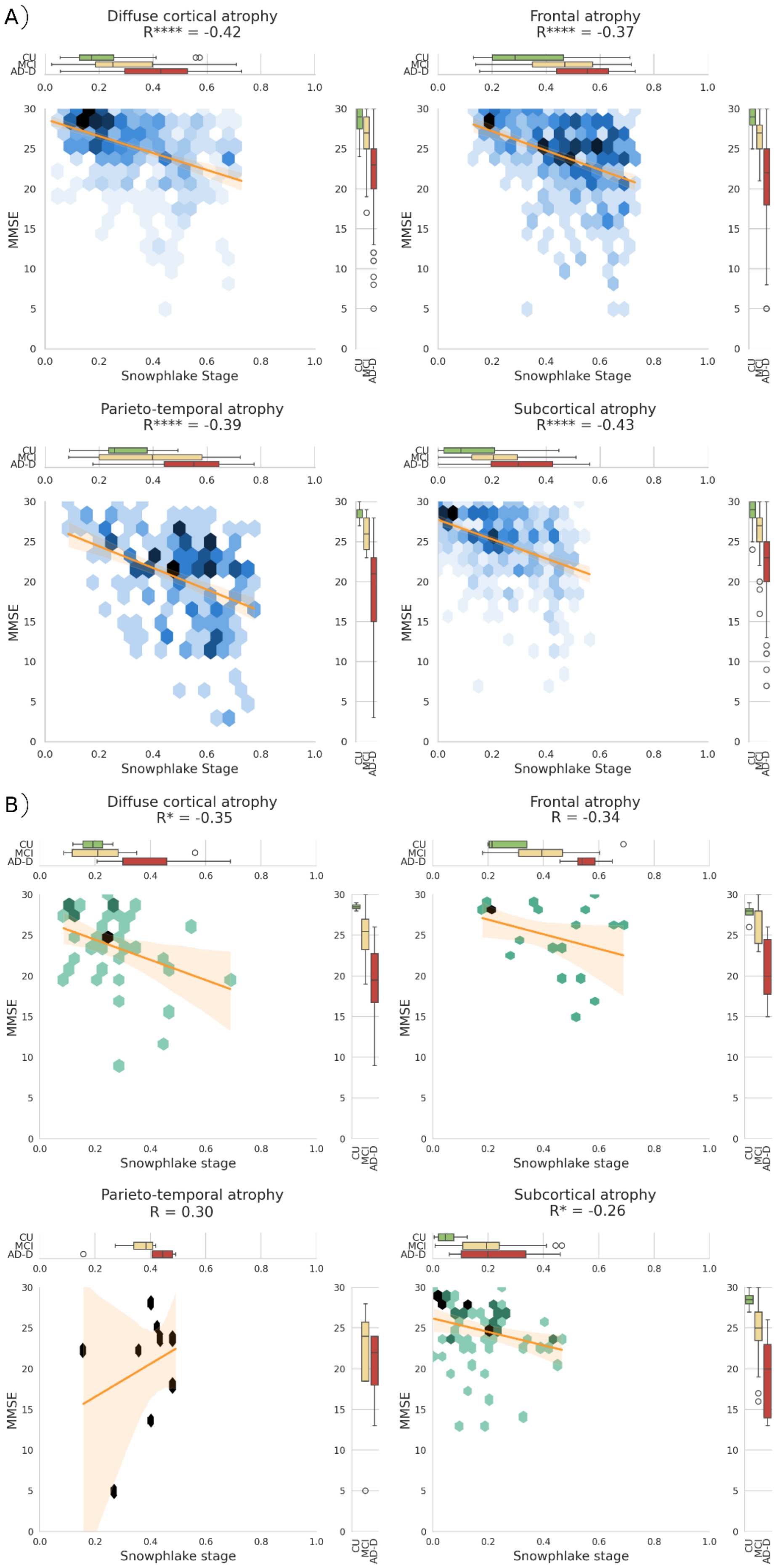

A)
Diffuse cortical atrophy
R**** = -0.42
Frontal atrophy
R**** = -0.37
Parieto-temporal atrophy
R**** = -0.39
Subcortical atrophy
R**** = -0.43
Snowphlake Stage
MMSE
CU
MCI
AD-D
B)
Diffuse cortical atrophy
R* = -0.35
Frontal atrophy
R = -0.34
Parieto-temporal atrophy
R = 0.30
Subcortical atrophy
R* = -0.26
Snowphlake stage

**Supplementary Figure 6: Experiment 1:** Correlation of the estimated stage (measuring atrophy severity) using Snowphlake with MMSE in A) Aβ+ validation-dataset B) Aβ+ subset of the external-validation cohort. Figures A) and B) both consists of 4 hex-plots, one for each subtype assigned using the trained Snowphlake model. The colour of a bin in the hex-plot denotes the relative proportion of the participants. The boxplot on top of each hex-plot shows the distribution of estimated Snowphlake stage for the participants in the different clinical groups. The boxplot at the right of each hex-plot shows the distribution of MMSE in the different clinical groups. The line overlaying on each hex-plot shows the regression line between MMSE and Snowphlake's stage. The text on top of each hex-plot shows the correlation coefficient (R) between estimated stage and MMSE. The asterisk (*) next to R denotes the significance level. * corresponds to $p < 0.05$; ** corresponds to $p < 0.01$; *** corresponds to $p < 0.001$; **** corresponds to $p < 0.0001$. Abbreviations: CU: Cognitively unimpaired (Cognitively normal or subjective cognitive decline); MCI: Mild cognitive impairment; AD-D: Alzheimer's disease dementia;

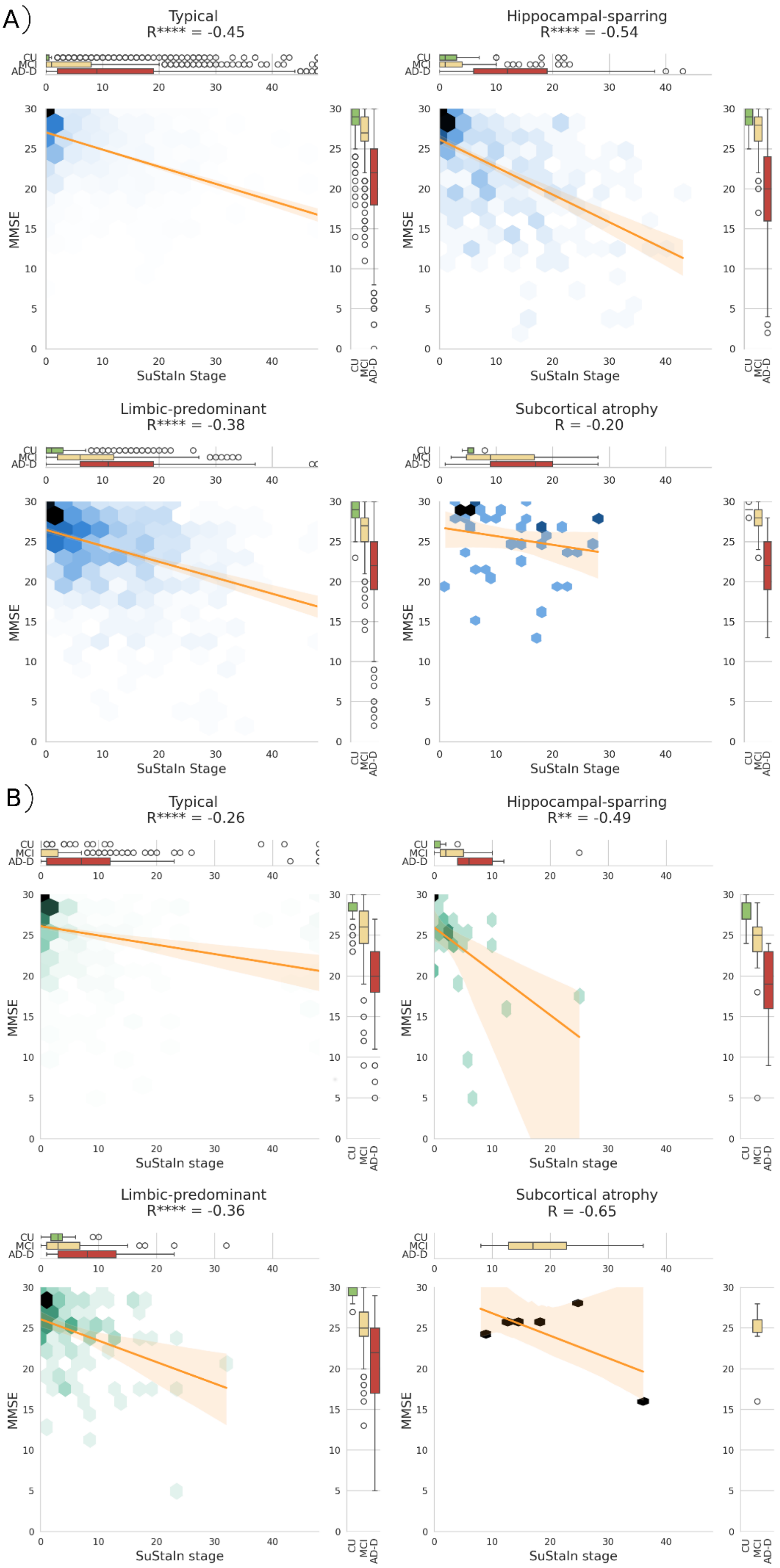

A)
Typical
R**** = -0.45
Hippocampal-sparring
R**** = -0.54
Limbic-predominant
R**** = -0.38
Subcortical atrophy
R = -0.20
B)
Typical
R**** = -0.26
Hippocampal-sparring
R** = -0.49
Limbic-predominant
R**** = -0.36
Subcortical atrophy
R = -0.65
CU
MCI
AD-D
MMSE
SuStaIn Stage
SuStaIn stage

**Supplementary Figure 7: Experiment 1:** Correlation of the estimated stage (measuring atrophy severity) using SuStaIn with MMSE in A) clinical-validation dataset B) external-validation cohort. Figures A) and B) both consists of 4 hex-plots, one for each subtype assigned using the trained SuStaIn model. The colour of a bin in the hex-plot denotes the relative proportion of the participants. The boxplot on top of each hex-plot shows the distribution of estimated SuStaIn stage for the participants in the different clinical groups. The boxplot at the right of each hex-plot shows the distribution of MMSE in the different clinical groups. The line overlaying on each hex-plot shows the regression line between MMSE and SuStaIn's stage. The text on top of each hex-plot shows the correlation coefficient (R) between estimated stage and MMSE. The asterisk (*) next to R denotes the significance level. * corresponds to $p < 0.05$; ** corresponds to $p < 0.01$; *** corresponds to $p < 0.001$; **** corresponds to $p < 0.0001$. Abbreviations: CU: Cognitively unimpaired (Cognitively normal or subjective cognitive decline); MCI: Mild cognitive impairment; AD-D: Alzheimer's disease dementia.

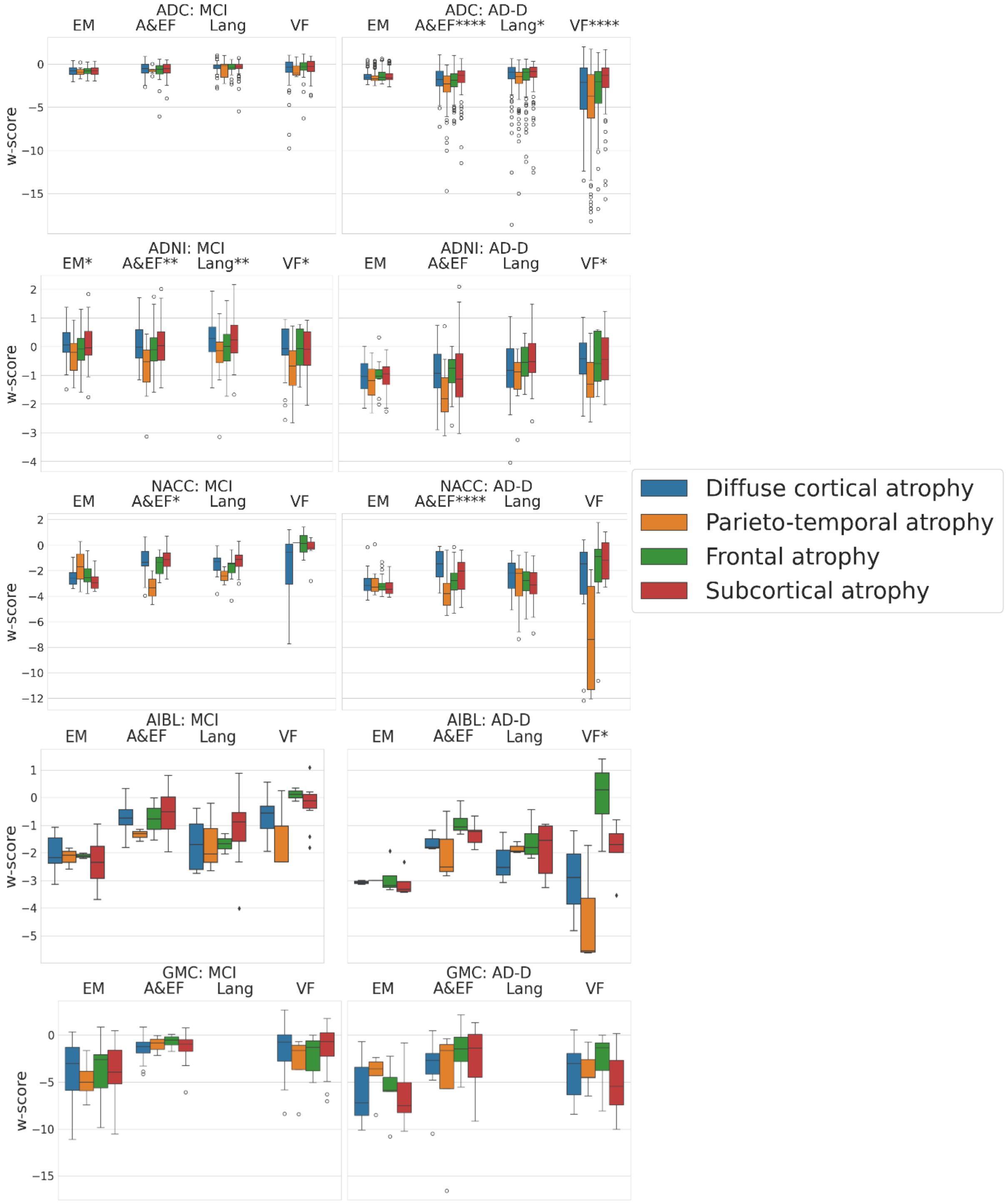


**Supplementary Figure 8: Experiment 2:** Cognitive domain scores of Aβ+ participants with MCI (left) or AD-D (right) in the validation cohorts, for the different subtypes assigned using the trained Snowphlake model. The plotted w-scores depict the raw cognitive domain scores, corrected for participant's age, sex, and education level. The asterisk (*) next to domain name denotes the significance level of the differences between the 4 subtypes, after correcting for the aforementioned confounders, in the corresponding cohort. * corresponds to $p < 0.05$; ** corresponds to $p < 0.01$; *** corresponds to $p < 0.001$; **** corresponds to $p < 0.0001$. Abberivations: EM: Episodic memory; A&EF: Attention and executive function; Lang: Language; VF: Visuospatial function.

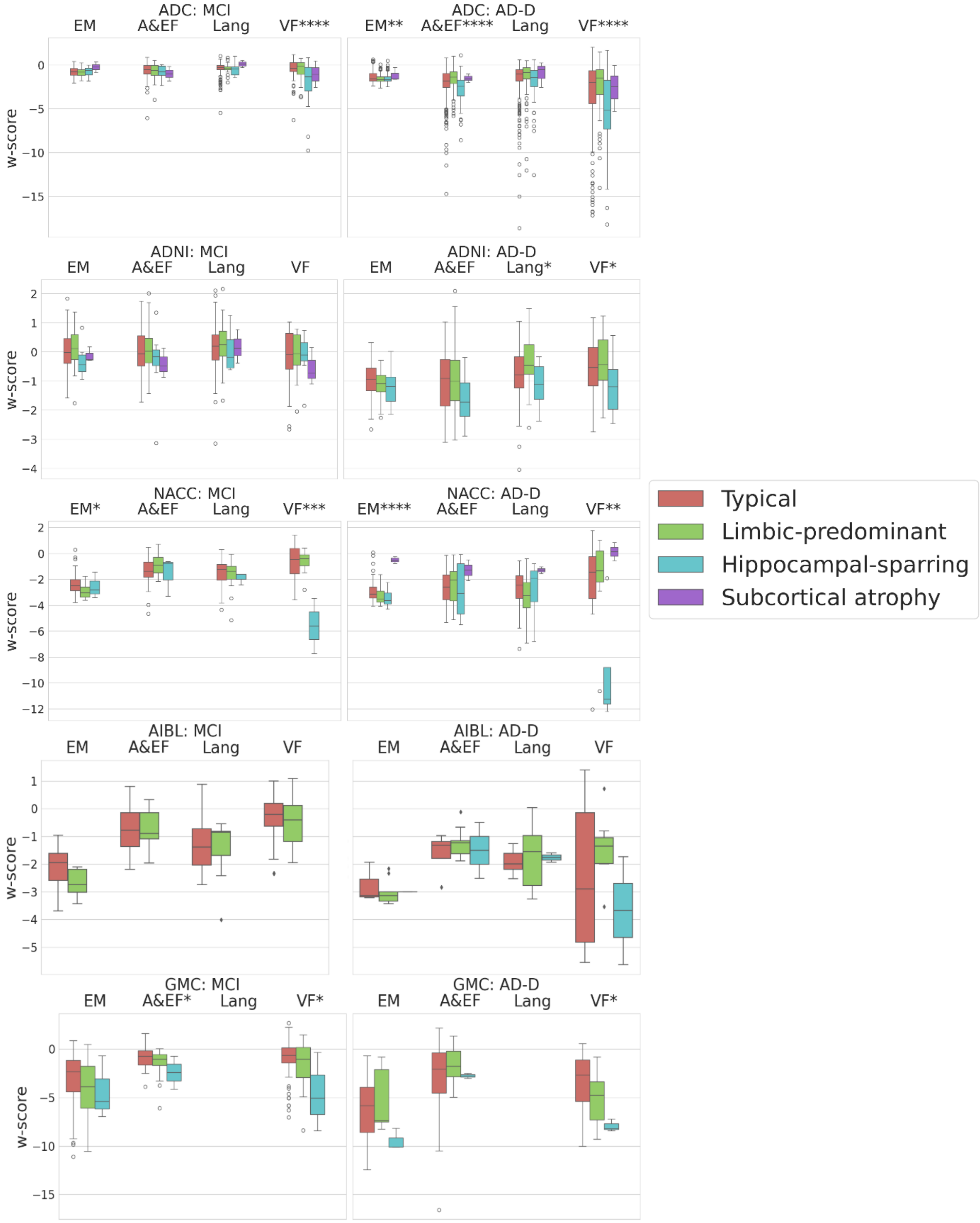


**Supplementary Figure 9: Experiment 2:** Cognitive domain scores of Aβ+ participants with MCI (left) or AD-D (right) in the validation cohorts, for the different subtypes assigned using the trained SuStaIn model. The plotted w-scores depict the raw cognitive domain scores corrected for participant's age, sex, and education level. The asterisk (*) next to domain name denotes the significance level of the differences between the 4 subtypes, after correcting for the aforementioned confounders, in the corresponding cohort. * corresponds to $p < 0.05$; ** corresponds to $p < 0.01$; *** corresponds to $p < 0.001$; **** corresponds to $p < 0.0001$. Abberivations: EM: Episodic memory; A&EF: Attention and executive function; Lang: Language; VF: Visuospatial function.

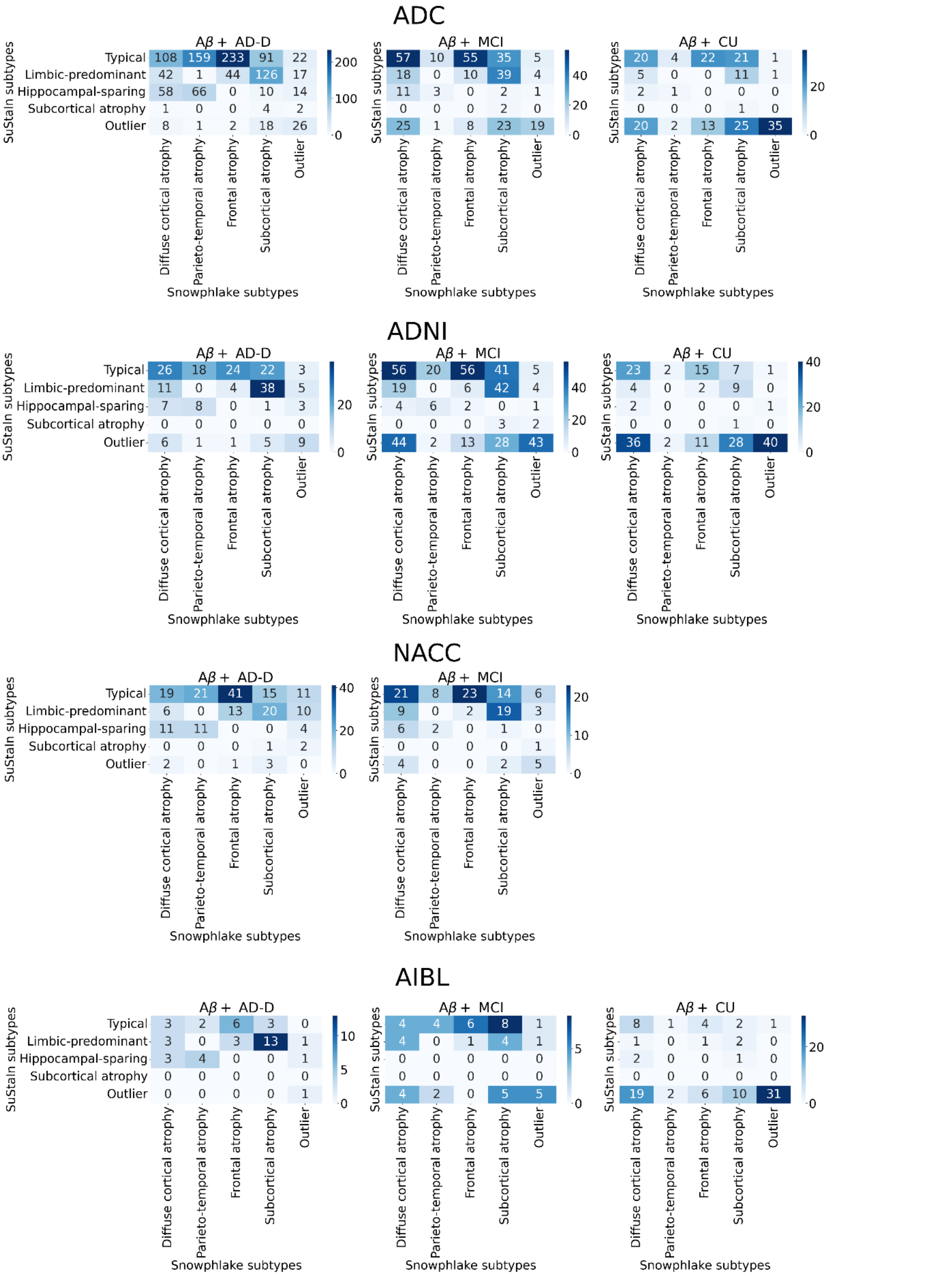


**Supplementary Figure 10: Experiment 3:** Cohort-wise concordance of Snowphlake and SuStaIn subtypes. The Figure shows the contingency matrix of estimated atrophy-based subtypes using Snowphlake and SuStaIn among Aβ+ participants in ADC, ADNI, NACC, and AIBL cohorts, in different clinical stages of the disease. Abbreviations: CU: Cognitively unimpaired (Cognitively normal or subjective cognitive decline); MCI: Mild cognitive impairment; AD-D: Alzheimer's disease dementia;

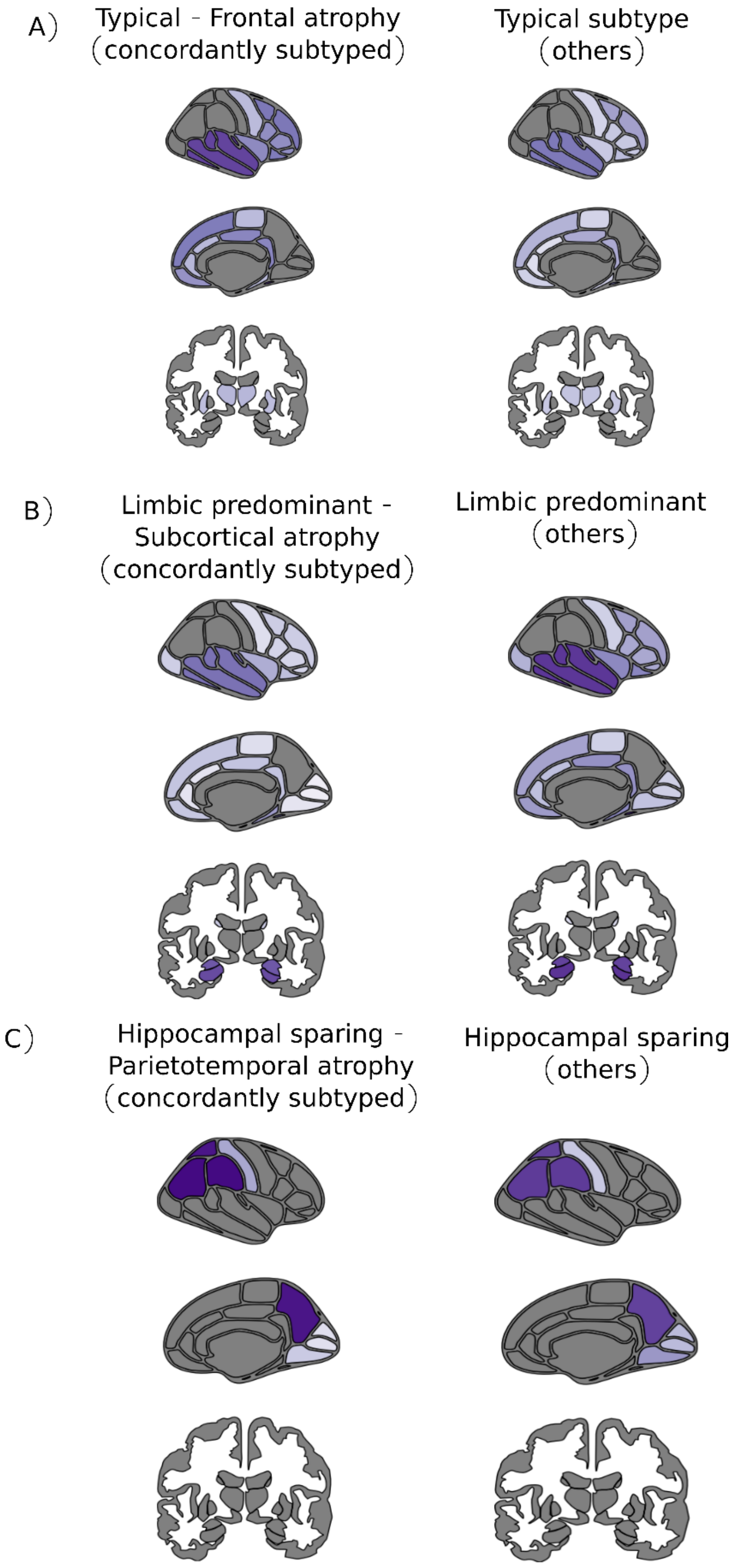


**Supplementary Figure 11: Experiment 3:** Regional differences (average w-score maps) between concordantly subtyped Aβ+ AD-D individuals and others assigned the same SuStaIn subtype. Only the regions with significant differences (FDR corrected $p < 0.05$) in average w-score values between the two groups are shown here. The average w-score were created using all the Aβ+ AD-D participants in the group and is thus not dependent on the disease severity. The colormap used shows the average w-score in that region, with light shades of violet indicating values closer to 0. Darker shades of violet indicates a more negative w-score value indicating more atrophy.
